# Snip Happens: A Retrospective Study of Vasectomy and Birth rates in Australia

**DOI:** 10.64898/2026.06.03.26354864

**Authors:** Jack Janetzki, Natansh Modi, Bianca Varney, Nicole Pratt, Michael Ward, Michael Wiese, Renly Lim, Lisa Kalisch Ellett

## Abstract

**Background:** Fertility rates in Australia have been declining over recent decades, reaching a record low total fertility rate of 1.48 births per woman in 2024. Concurrently, vasectomy remains widely accessible and increasingly normalised as a permanent contraceptive option. Despite extensive commentary on falling birth rates, no contemporary Australian study has examined vasectomy rates relative to birth rates over time. We aimed to compare population level vasectomy and birth rates across Australian jurisdictions and age groups.

**Study design:** Nationwide retrospective time-series study. Retrospective population-based study using Medicare Benefits Schedule item 37623 to identify vasectomy procedures performed between July 2015 and December 2024. Rates were calculated per 100,000 male population using quarterly Australian Bureau of Statistics (ABS) population estimates and summarised as rolling 12-month averages. Birth rates were derived using matched ABS data for women across equivalent age strata (18–24, 25–34, 35–44 years).

**Results:** Vasectomy rates increased nationally from 32 per 100,000 in 2016 to 55 per 100,000 in 2023 before declining modestly in 2024. Birth rates declined from 5,200 to 3,800 per 100,000 over the same period. Trends were consistent across states and age groups, with the greatest vasectomy uptake in men aged 35–44 years.

**Conclusion:** Australia is undergoing a demographic shift characterised by rising vasectomy uptake and declining fertility. We have observed changing secular trends in two related but distinct demographic indicators of reproductive intention. Ongoing monitoring of permanent and long-acting contraception is essential to understand evolving population dynamics and inform reproductive health policy.

**Short summary for non-experts:** This nationwide study demonstrates that rising vasectomy uptake in Australia coincides with declining birth rates, particularly among men aged 35–44 years. Monitoring permanent contraceptive use alongside fertility trends may help clinicians and policymakers better understand changing reproductive intentions and inform future reproductive health planning.

**Short summary for non-experts:** Australia’s birth rate has fallen to record lows, raising growing concerns about how changing reproductive choices may shape future society and healthcare needs. In this study, we found that vasectomy rates increased steadily across Australia between 2016 and 2024 while birth rates declined, particularly among adults aged 35-44 years. These findings suggest Australian men are increasingly choosing permanent contraception and highlight the need for ongoing monitoring of reproductive health trends to inform future policy and service planning.

## 1. Introduction

Human reproduction is commonly assumed to outpace most other biological processes, assisted by enthusiasm, optimism, and a general disregard for sleep. In Australia, births have historically outnumbered other contributors impacting population changes, including deaths and migration.(1) This trend is essential to sustain natural population growth, maintain a stable age structure, and support ongoing economic and social development. Birth rates often rise after holiday periods, particularly during festive seasons, and are also influenced by seasonal climate patterns across Australia’s diverse regions.(2) By contrast, vasectomy is an intentional, clinic-based intervention that requires planning, consent, and an awareness of anatomy.

The fertility rate in Australia has shifted markedly. Like many high-income countries, Australia has experienced sustained declines in fertility, recently reaching a record low rate of 1.48 births per woman, continuing a downward trend that began in the 1960s.(2-4) Delayed parenthood, a rise in the proportion of women who remain childless, and a trend toward smaller families among those who do become parents are the three major contributors to the steady fall in births over recent decades.(5-7) These changes are driven by economic uncertainty, the high cost of raising children, career goals, access to affordable childcare, and shifting attitudes toward family size.(8-10) At the same time, vasectomy has remained a widely available, highly effective, relatively inexpensive and permanent form of contraception that is increasingly framed as a shared responsibility within long-term partnerships.

Despite extensive commentary on falling birth rates, little attention has been paid to how these trends compare with population-level rates of permanent contraception. Vasectomy is rarely framed as a demographic counterweight to birth itself and no study has directly examined vasectomy rates alongside national birth rates over time to determine whether the former is beginning to eclipse the latter. Moreover, some may also consider vasectomy to be an alternative permanent form of contraception compared to female sterilisation such as tubal ligation and be considered to be of lower risk of complications.(11) Limited studies have investigated historical trends of vasectomy procedures, however this has not been reviewed recently. In an Australian study from 1993-94 to 2002-03 there was a peak in vasectomies in 1996-97 followed by declining procedure rates in all Australian states or territories except the Australian Capital Territory.(12) Moreover, an increase in the mean age at which a person received the procedure was observed throughout the study period.(12)

We therefore conducted a retrospective, population-based study examining vasectomy procedures in Australia between July 2015 and December 2024. Our primary aim was to describe temporal trends in vasectomy rates and compare them with contemporaneous birth rates, to assess whether vasectomy activity has approached or exceeded the rate of births in Australia.

## 2. Materials and Methods

In Australia, the Medicare Benefits Schedule (MBS) provides a list of billable codes that relate to subsidised medical services provided by health practitioners, primarily medical practitioners.(13) Claims for vasectomy procedures were identified using the Medicare item number 37623 (“vasotomy or vasectomy, unilateral or bilateral”), extracted from the Australian Government Services Australia Medicare Item Reports database.(14) Vasectomies performed between July 2015 and December 2024 were obtained from the patient demographic reports by Australian state or territory and month.(14) The number of procedures in this report is stratified by gender and age range. Records of procedures for males aged 15-24, 25-34 and 35-44 years were retrieved. Though the database lists males aged 15-24 years, a person must be at least 18 years of age to have a vasectomy in Australia(15); therefore this age group relates to adult males aged 18-24.

We examined the rate of vasectomy across all Australian states and territories, stratified by age and male sex. Monthly vasectomy rates per 100,000 male population were calculated by dividing the number of vasectomy procedures by the corresponding quarterly male population estimates from the Australian Bureau of Statistics (ABS) ‘Quarterly Population Estimates (ERP), by State/Territory, Sex and Age,’ and multiplying by 100,000. Rates were standardised per 100,000 population to enable comparisons across jurisdictions. The same population estimate was used for each of the three months within a corresponding quarter. Standardising the number of vasectomies performed per 100,000 population allows for comparison of rates between jurisdictions. The rolling 12-month average of vasectomies performed per 100,000 population was then calculated using a look back period of 12 months beginning at the start of the study period(July 2015).(16). This provided a clearer representation of underlying trends in rate change over time, by smoothing short-term fluctuations and seasonal effects.

The number of births by age of mother and in each state or territory was also obtained from the Australian Bureau of Statistics using the same age brackets of 18-24, 25-34 and 35-44 years.(17) Quarterly female population estimates were obtained from the ABS to determine the number of births by age of mother per 100,000 population in each state or territory.(16) Standardisation to the state or territory population was performed as described above for vasectomies and the rolling 12-month averages were calculated in the same way.

All statistical analyses were conducted using R (version 4.5.2) and RStudio (version 2026.01.0); R Foundation for Statistical Computing, Vienna, Austria). Data was subsequently visualised in Excel (Version 2512).

### 2.1 Ethics statement and data sources

The study used de-identified, publicly available administrative data from the MBS and ABS. As all data were aggregated and contained no personal identifiable information, the study did not require ethics approval. This study was reported in accordance with the RECORD-PE statement.

## 3. Results

### 3.1 Australia-wide

Between June 2016 and December 2024, overall vasectomy and birth rates in Australia demonstrated opposing temporal trends across all age groups examined. Vasectomy rates increased steadily over the study period in all age groups and overall (Table 1). Birth rates declined in all age groups and overall.

**Table 1:** Rolling average vasectomies per 100,000 male populationa and rolling average of births per 100,000 female population in June each year of study period.

| Month and year | Rolling average vasectomies per 100,000 male population |  |  |  | Rolling average births per 100,000 female population |  |  |  |
| --- | --- | --- | --- | --- | --- | --- | --- | --- |
|  | Vasectomies |  |  |  | Births |  |  |  |
|  | 18-24 | 25-34 | 35-44 | Total | 18-24 | 25-34 | 35-44 | Total |
| Jun-16 | 0.33 | 21 | 71 | 33 | 2968 | 7269 | 2208 | 4393 |
| Jun-17 | 0.47 | 21 | 74 | 34 | 2743 | 6967 | 2249 | 4247 |
| Jun-18 | 0.35 | 22 | 77 | 36 | 2557 | 6677 | 2169 | 4063 |
| Jun-19 | 0.54 | 23 | 82 | 38 | 2455 | 6504 | 2196 | 3979 |
| Jun-20 | 0.56 | 23 | 79 | 37 | 2281 | 6411 | 2210 | 3909 |
| Jun-21 | 0.74 | 29 | 95 | 46 | 2274 | 6622 | 2226 | 3995 |
| Jun-22 | 0.87 | 29 | 92 | 46 | 2163 | 6524 | 2265 | 3936 |
| Jun-23 | 1.08 | 35 | 106 | 53 | 1946 | 5944 | 2122 | 3599 |
| Jun-24 | 0.89 | 32 | 102 | 50 | 1816 | 5648 | 2084 | 3436 |

Vasectomy procedures were most prominent in the 35-44 year old age group and births were most prominent in the 25-34 year old age group. Full month by month changes in the rolling average vasectomies and births per 100,000 male or female population for Australia can be found in Appendix 1.

### 3.2 State-based analysis

Vasectomy and birth trends varied by Australian states (Figure 1 and Figure 2). New South Wales and Victoria in general showed steady rises in vasectomy rates alongside gradual declines in birth rates across the entire study period, with minor declines in vasectomies during early to mid-2020 (Figure 1 Panel A). Queensland consistently reported the highest vasectomy rates of all states across the study period. South Australia’s vasectomy rates were low at the start of the study period but eventually surpassed those of Victoria, while Western Australia, Tasmania, and the ACT showed moderate increases in the rate of vasectomies over the study period. The Northern Territory had lower vasectomy rates overall.

**Figure 1:**
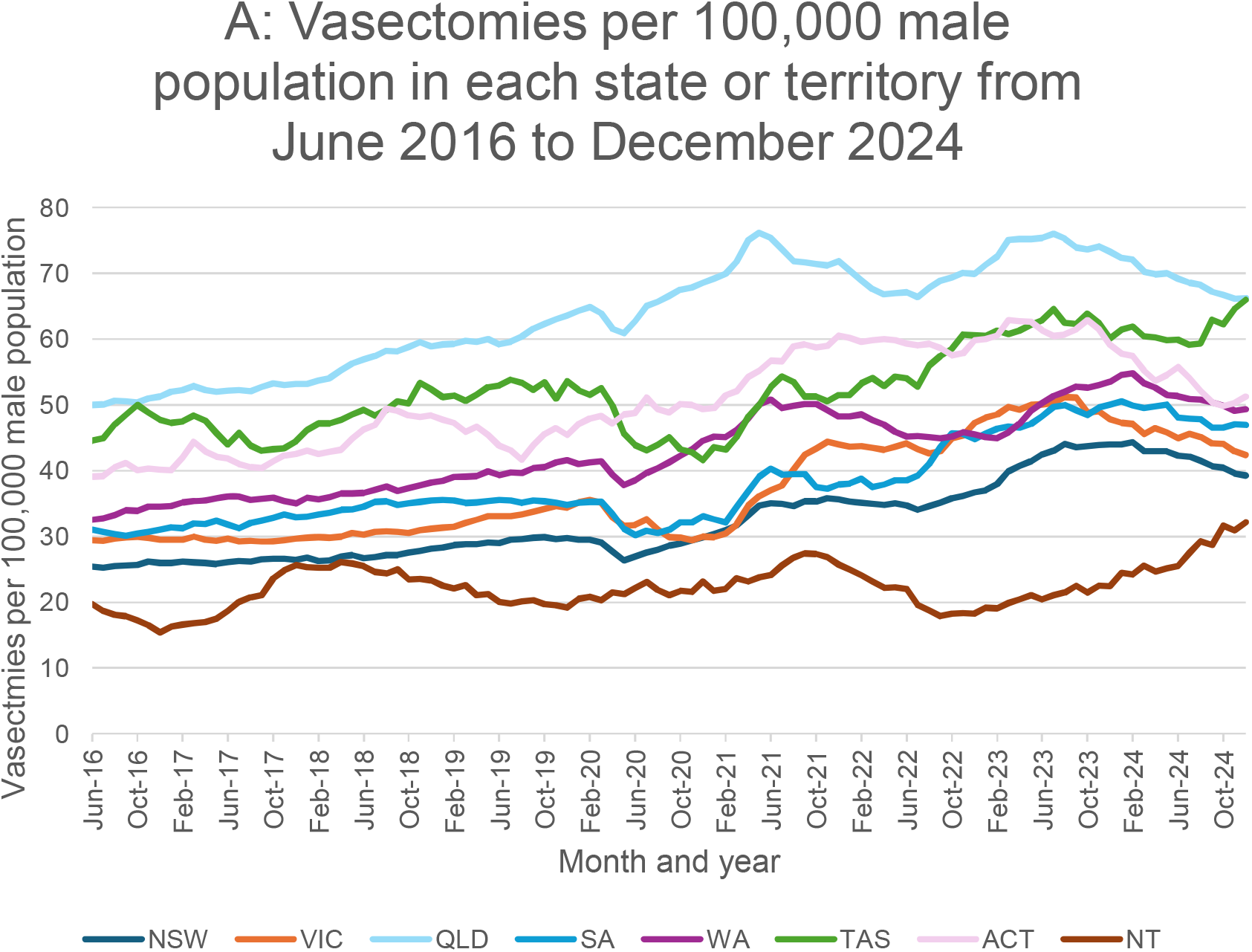
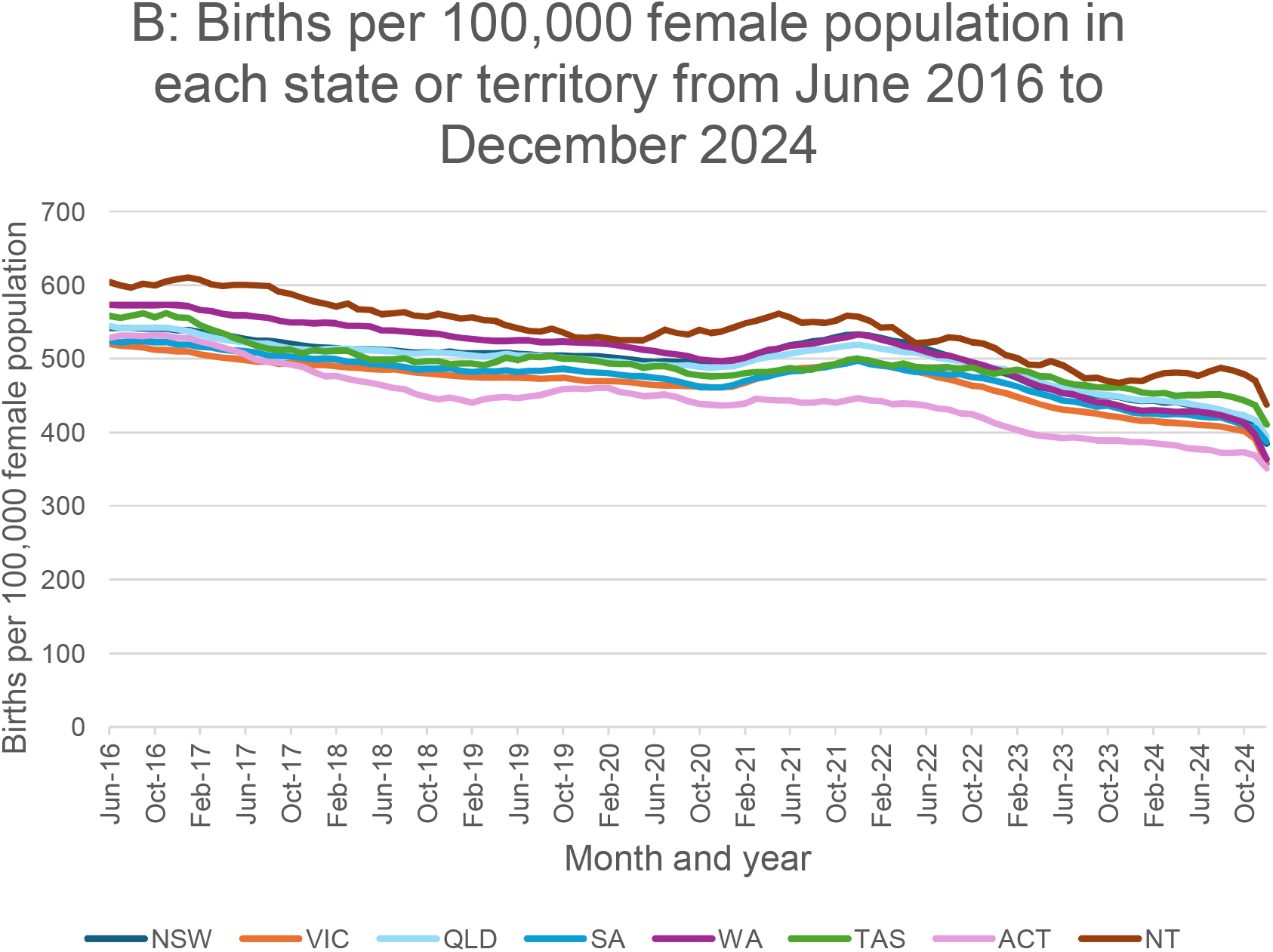
Panel A) Rolling average of vasectomies per 100,000 male population in each state and territory from June 2016 to December 2024.Panel B) Rolling average of births per 100,000 female population in each state or territory from June 2016 to December 2024.

**Figure 2:**
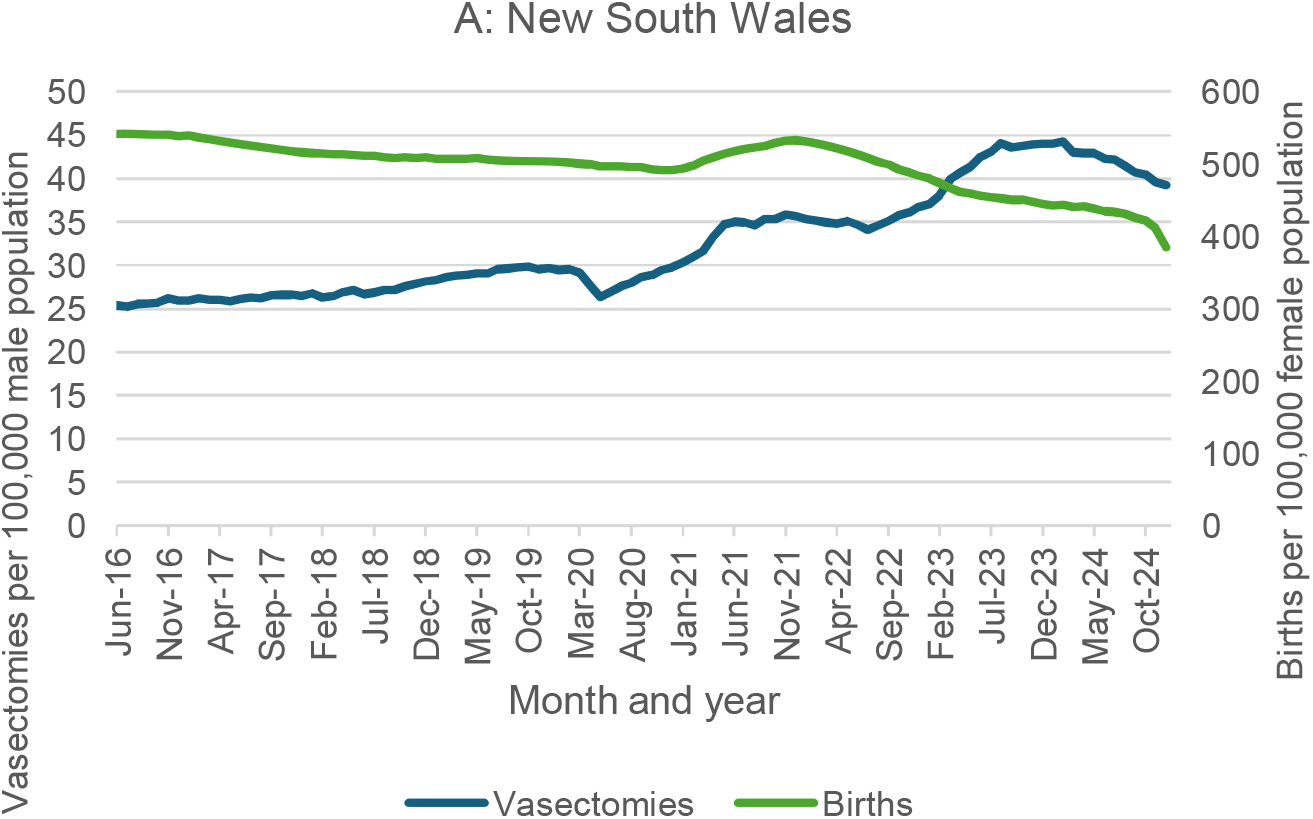
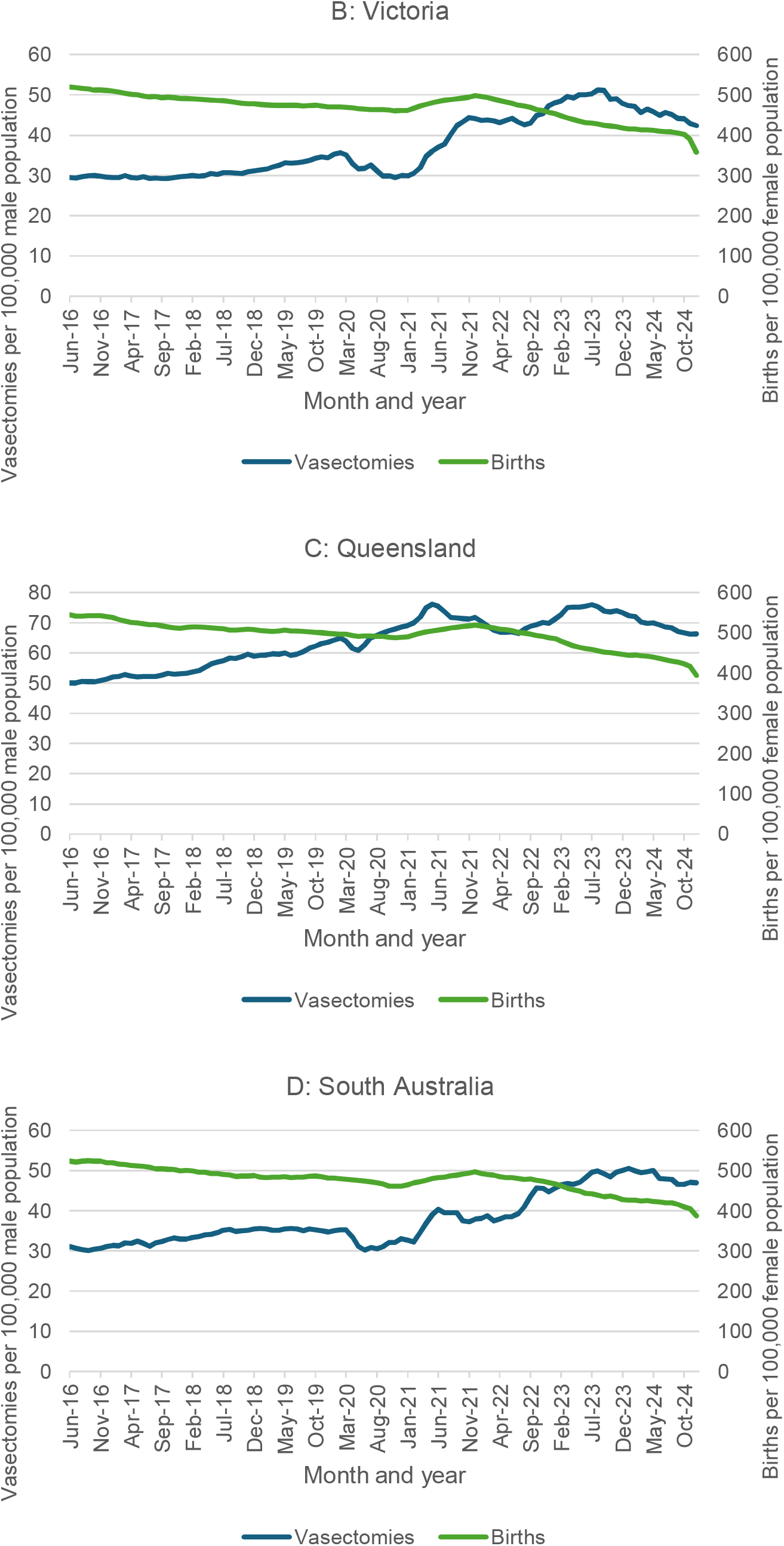
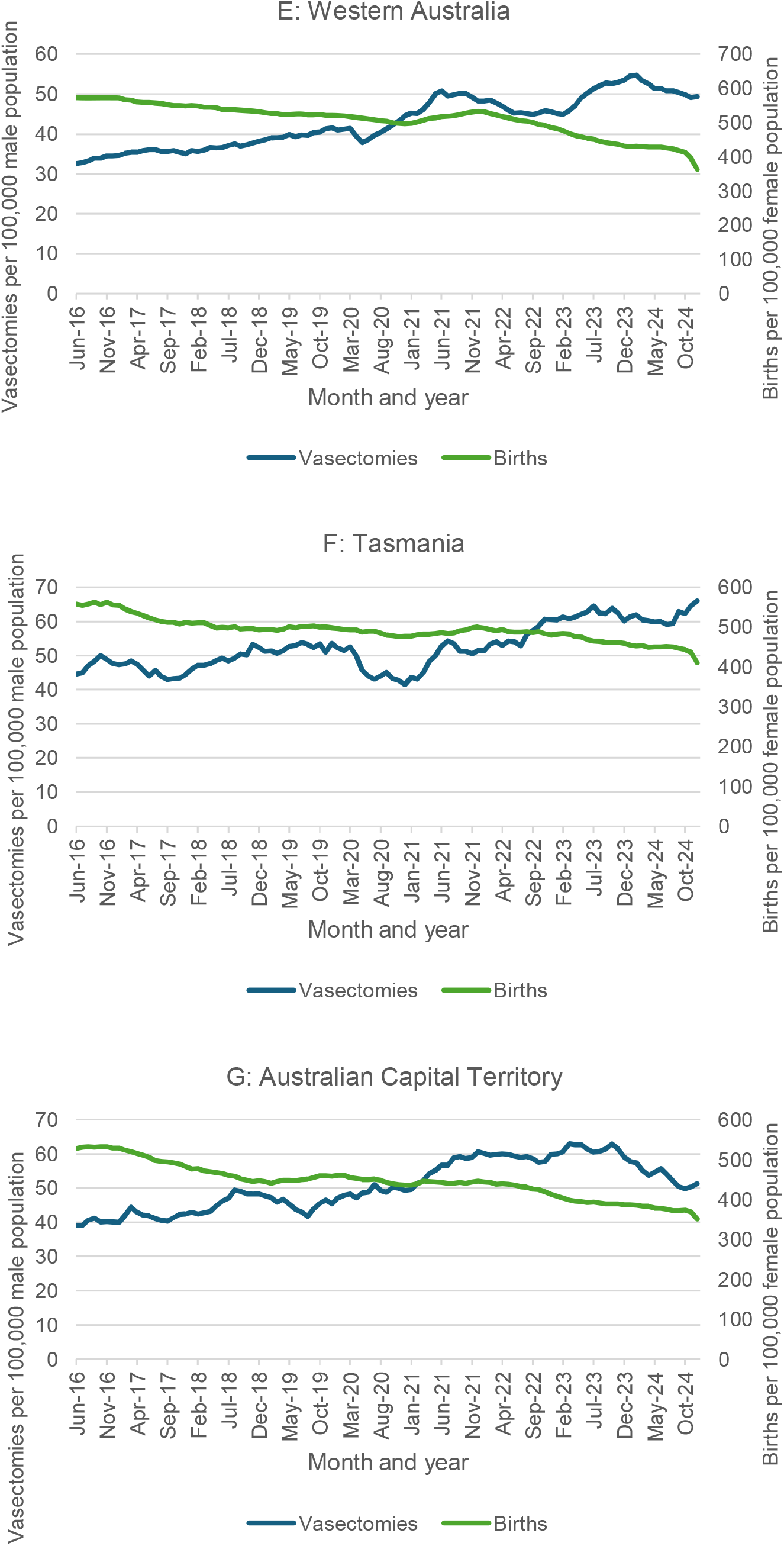
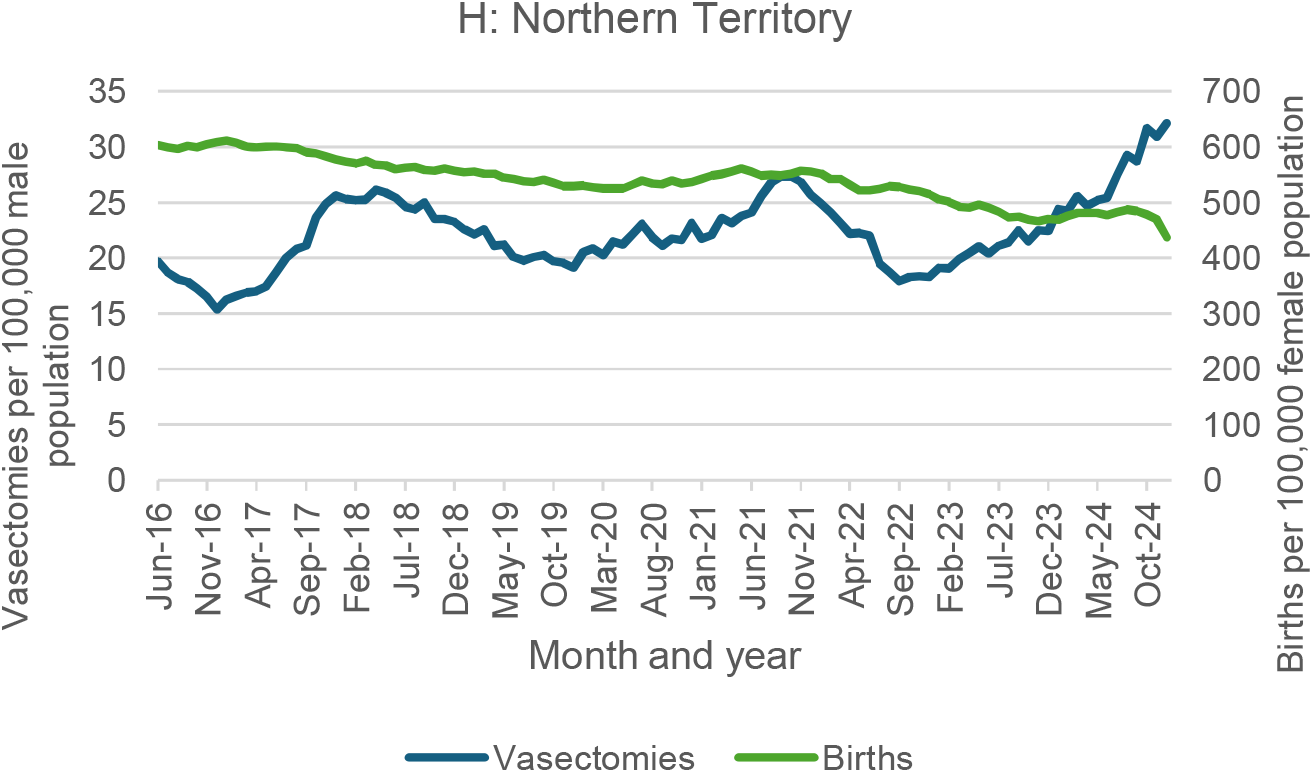
Rolling 12-month average vasectomies and births per 100,000 male and female population respectively for people aged 18-44 years from April 2016 to December 2024 in A) New South Wales, B) Victoria, C) Queensland, D) South Australia, E) Western Australia, F) Tasmania, G) Australian Capital Territory and H) Northern Territory.

Across all states, birth rates generally declined across the study period, with short-lived increases in some regions around 2021–2022, though these increases were not observed in all states (for example in Tasmania and ACT) (Figure 1 Panel B). See Appendix A for detailed state-level analysis.

Individual state or territory birth and vasectomy rates are overlayed in Figure 2 Panels A to H.

## 4. Discussion

This retrospective population-based study provides the first contemporary comparison of vasectomy and birth rates across Australia. We observed a sustained increase in vasectomy rates in Australia alongside a concurrent decline in birth rates. The decline in Australian birth rates is consistent with well-described social and economic shifts, including delayed childbearing, rising housing costs, and changing expectations around work and family life.(18, 19) Contemporary Australian economic analyses explicitly identify financial pressures as a major deterrent to family formation and family expansion, with marked regional variation in fertility outcomes.(20) The patterns observed in this study, which indicate declining birth rates alongside increasing vasectomy rates across jurisdictions, are consistent with broader national demographic trends. Australia’s total fertility rate declined to 1.50 in 2023 and further to 1.48 in 2024, representing the lowest recorded level and the largest year-on-year decline since the early 1970s.(3) The increasing normalisation of vasectomy as a shared contraceptive responsibility may also contribute to its rising uptake, particularly as alternative contraceptive options, including permanent methods such as tubal ligation, place a disproportionate physical and logistical burden on female partners.

A transient spike in vasectomy procedures was observed during mid-2020, coinciding with the COVID-19 pandemic. This is likely attributable to hospital restrictions that limited access to many elective surgical procedures,(21-23) which may have prompted private urologists to shift their caseloads toward interventions like vasectomies that could be performed in consultation-room settings. Vasectomies that might otherwise have occurred later may therefore have been brought forward during this period.

Importantly, our findings do not imply that vasectomy causes declining birth rates, nor that individuals undergoing vasectomy would otherwise have contributed to national fertility statistics. From a population perspective, the balance between births and vasectomies provides a revealing indicator of collective reproductive intent. Internationally, Australia’s experience sits within a broader context of declining fertility and heterogeneous uptake of male sterilisation across high-income countries. Vasectomy prevalence varies widely; in 2019, 11.3% of males aged 15-49 years in the United States of America had had a vasectomy.(24) In 2015 in New Zealand 10% of males aged 16-49 years had had a vasectomy compared to only 1.4% of men in China in 2017 aged 15-49 years. There is also substantially lower prevalence of vasectomy across most low- and middle-income countries, particularly in Africa and Southern Asia.(24) International comparisons highlight that the drivers of vasectomy uptake differ substantially across sociopolitical contexts. In the United States, increased vasectomy interest following the Dobbs decision (which affected access abortion services and allowed states to set their own abortion regulations) has been linked to heightened reproductive rights uncertainty rather than economic pressures alone.(25) This highlights the importance of situating Australian trends within their specific economic, social and political context and cautions against assuming uniform drivers across jurisdictions.

This study has several limitations. Using the data available for this study we could not capture vasectomies that were performed privately; vasectomies for which no Medicare claim was made. Although the numbers are likely small, this may have led to undercounting of vasectomies performed each month. Inclusion of intrauterine device (IUD) insertion in the analysis was also not possible, because Medicare Benefits Schedule data for IUD insertions includes females aged 15-24 years, and we were unable to restrict the dataset to those aged 18 years or older without compromising interpretability. Moreover, people may have been included in the analysis twice due to multiple IUD insertions over their lifetime and IUD insertion is not a permanent sterilisation method as fertility returns after removal. We were also unable to account for the use of other deliberate sterilisation methods in this study, particularly tubal ligation, because the MBS item codes which are used to claim for this procedure (item 35637) also include other procedures which are not related to permanent contraception. These limitations precluded meaningful inclusion of these procedures. One study conducted in New South Wales which used hospital data admission data between 2010 and 2019 found, however, a declining number of cases of female sterilisation and a sterilisation rate decline from 22.6 to 5.4 per 10,000 females between 2010 and 2019. Sterilisation incidence was most highest in the 35-39 age group over the study.(26)

Internationally, vasectomy prevalence is typically reported within defined reproductive-age populations, most commonly 15-49 or 18-49 years, to facilitate demographic comparability. This is particularly important when interpreting temporal change, as vasectomy typically occurs only once per male, so the outcome is best reported as a population prevalence. Inclusion of older age groups (≥50 years) may inflate estimates of “ever having had a vasectomy” through cohort ageing rather than reflecting contemporary behavioural change. Prior Australian analyses restricted to reproductive-age cohorts have reported largely stable vasectomy prevalence between 2006 and 2016, suggesting that any recent increases warrant careful contextual interpretation rather than being assumed to represent long-term growth.(24) From a comparability and interpretive standpoint, limiting analyses to ages 18-44 years has been shown to provide more meaningful insights than analyses inclusive of all adult ages.(24) It should also be noted that males may not father children with women of the same age and therefore comparison between the rates in the figures above with respect to age should be carefully considered.

## 5. Conclusion

In this nationwide retrospective analysis, vasectomy rates in Australia rose steadily over the past decade while birth rates declined across all states and age groups examined. Although vasectomy rates remain substantially lower than birth rates overall, the increasing rate of vasectomies signals a shift in reproductive intentions. Increased uptake of vasectomy, particularly among men aged 25–44 years, coincides with decreasing fertility rates and suggests growing male engagement in permanent contraception.

Whether this divergence in permanent sterilisation and birth rates represents a temporary response to uncertainty or a sustained demographic shift remains unclear. Ongoing monitoring of permanent and long-acting contraception across genders will be important for understanding future population dynamics and informing reproductive health policy.

## Author statements

### Conflicts of interest

All authors have no conflicts of interest to declare.

### Data availability

The data that support this study are available in https://medicarestatistics.humanservices.gov.au/statistics/mbs_item.html and https://www.abs.gov.au/statistics/people/population

### Author contributions

Jack Janetzki: conceptualisation, data curation, formal analysis, investigation, methodology, project administration, resources, software, visualisation, writing -original draft

Natansh Modi: investigation, methodology, supervision, visualisation, writing – review and editing

Bianca Varney: methodology, supervision, visualisation, writing – review and editing Nicole Pratt: supervision, writing – review and editing

Michael Ward: conceptualisation, supervision, visualisation, writing – review and editing Michael Wiese: supervision, writing – review and editing

Renly Lim: supervision, writing – review and editing

Lisa Kalisch Ellett: conceptualisation, investigation, methodology, supervision, visualisation, writing – review and editing

### Declaration of funding

This research did not receive any specific grant from funding agencies in the public, commercial, or not-for-profit sectors.

### Ethics statement

The study used de-identified, publicly available administrative data from the Medicare Benefits Schedule and Australian Bureau of Statistics. As all data were aggregated and contained no personal identifiable information, the study did not require ethics approval.

