## Appendices for "Snip Happens: A Retrospective Study of Vasectomy and Birth rates in Australia"

In New South Wales (Figure 2A), vasectomy rates increased from 25 per 100,000 male population per month in April 2016 to approximately 40 per 100,000 male population in December 2024. A small reduction in vasectomies was observed in early 2020 before peaking at 44 per 100,000 male population in February 2024. In April 2016 there were an average of 540 births per 100,000 female population which reduced to 410 per 100,000 female population by November 2024. There was a slight observable increase in average births per 100,000 female population between February 2021 and October 2022 before the declining trajectory continued.

In Victoria (Figure 2B), there was a slightly greater number of vasectomies performed of 29 per 100,000 male population per month in April 2016 compared to NSW, and a slightly greater number performed of 42 per 100,000 male population per month at the end of the study period. Despite this, there was also a considerable decline in vasectomies in early 2020 to early 2021 as observed in NSW. The birth rate per 100,000 female population in VIC was similar to NSW at 521 in April 2016 declining to 390 per 100,000 female population per month in November 2024. The same slight observable increase in average births per 100,000 female population was observed in Victoria as was seen in NSW during the study period.

Queensland average monthly vasectomies were 50 per 100,000 male population per month in April 2016 increasing to 66 per 100,000 male population per month in December 2024 (Figure 2C); far beyond that observed in NSW and Victoria. In contrast to the observed decline in vasectomies seen throughout the study period in other states, vasectomies performed increased throughout 2020 and early 2021 to a peak of 76/100,000 population/month in May 2021. There was a subsequent decline thereafter followed by a subsequent peak in July 2023 of 76/100,000 population/month before declining thereafter. In comparison, the number of births declined throughout the study period from 541 per 100,000 female population per month in April 2016 to 394/100,000 female population/month in December 2024. There was a brief yet marginal increase in average births per 100,00 female population towards the end of 2021 and early 2022.

South Australian men had lower rates of vasectomies performed per 100,000 male population per month compared to Victoria and NSW (30 per 100,000 male population/month in April 2016 (Figure 2D)) but not QLD at the start of the study period. The number of vasectomies performed per 100,000 male population per month increased to 47 per 100,000 male population per month by the end of the study period, eclipsing that observed in NSW and Victoria. There was brief decline in vasectomies performed each month between June 2020 and February 2021 and again in October 2021 to June 2022. Births per 100,000 female population per month were similar to that observed in the aforementioned states at 526 in April 2016 and reducing to 405 by November 2024.

In Western Australia, there were an average of 32 vasectomies performed per 100,000 male population in April 2016 increasing to 49 per 100,000 male population in December 2024 (Figure 2E). During the study period there were two notable peaks in average vasectomies performed: 50 in July-October 2021 and 55 in January and February 2024. Both peaks were preceded by brief declines in the number of vasectomies performed in the preceding months. The number of births in WA declined from 572 per 100,000 female population/month in April 2016 to 396 by November 2024. Similar to that observed in other states, there was a brief increase in average births per 100,000 female population in late 2021 to early 2022 before declining again thereafter.

Tasmanian’s had similar rates of vasectomies performed per 100,000 male population per month compared to other states at 41 in April 2016 and increasing to 66 in December 2024 (Figure 2F). A decline in vasectomies performed per month was observed between May 2020 and March 2021 before rising again for the rest of the study period. The number of births per 100,000 female population per month was also comparable to other states of 554 in April 2016 declining to 437 in November 2024. The increase in births that were seen in 2021 and 2022 in the aforementioned states was not observed in Tasmania.

In the ACT, the number of vasectomies performed in April 2016 was an average of 39 per 100,000 male population increasing to 51 per 100,000 male population by the end of the study period (Figure 2G). The average number of vasectomies performed increased throughout the study period peaking at 63 per 100,000 male population in October 2023 before declining thereafter. Births per 100,00 female population were similar to that observed in other states at 525 per 100,000 female population in April 2016, decreasing to 351 in December 2024. Similarly to Tasmania, the increase in births observed in other states in 2021 and 2022 was not observed in the ACT.

Vasectomies performed per 100,000 male population per month varied throughout the study period in the NT (Figure 2H). In April 2016, the number of vasectomies performed was lower than other states at 21 per 100,000 male population and increased to 32 per 100,000 male population by December 2024. Throughout the study period there were two major peaks in number of vasectomies performed in early 2018 and late 2021. In both cases, the number of vasectomies performed per month decreased for several months thereafter.

New South Wales

Rolling average vasectomies and births per 100,000 male and female population respectively for 18-24 year olds from April 2016 to December 2024

Vasectomies per 100,000 male population

Births per 100,000 female population

| Month and year | Vasectomies |
| --- | --- |
| 2016-04-01 | 0.163351 |
| 2016-05-01 | 0.173139 |
| 2016-06-01 | 0.181296 |
| 2016-07-01 | 0.226206 |
| 2016-08-01 | 0.22592 |
| 2016-09-01 | 0.270544 |
| 2016-10-01 | 0.293004 |
| 2016-11-01 | 0.292719 |
| 2016-12-01 | 0.314894 |
| 2017-01-01 | 0.359319 |
| 2017-02-01 | 0.336753 |
| 2017-03-01 | 0.336753 |
| 2017-04-01 | 0.336405 |
| 2017-05-01 | 0.358295 |
| 2017-06-01 | 0.33571 |
| 2017-07-01 | 0.312879 |
| 2017-08-01 | 0.290423 |
| 2017-09-01 | 0.245137 |
| 2017-10-01 | 0.222676 |
| 2017-11-01 | 0.200216 |
| 2017-12-01 | 0.155295 |
| 2018-01-01 | 0.132741 |
| 2018-02-01 | 0.132741 |
| 2018-03-01 | 0.154612 |
| 2018-04-01 | 0.154199 |
| 2018-05-01 | 0.175199 |
| 2018-06-01 | 0.218848 |
| 2018-07-01 | 0.218403 |
| 2018-08-01 | 0.218403 |
| 2018-09-01 | 0.217959 |
| 2018-10-01 | 0.239625 |
| 2018-11-01 | 0.326289 |
| 2018-12-01 | 0.326289 |
| 2019-01-01 | 0.325911 |
| 2019-02-01 | 0.347404 |
| 2019-03-01 | 0.368519 |
| 2019-04-01 | 0.368263 |
| 2019-05-01 | 0.324358 |
| 2019-06-01 | 0.280709 |
| 2019-07-01 | 0.259074 |
| 2019-08-01 | 0.345625 |
| 2019-09-01 | 0.323991 |
| 2019-10-01 | 0.324253 |
| 2019-11-01 | 0.259517 |
| 2019-12-01 | 0.259517 |
| 2020-01-01 | 0.238024 |
| 2020-02-01 | 0.216531 |
| 2020-03-01 | 0.23945 |
| 2020-04-01 | 0.217881 |
| 2020-05-01 | 0.196312 |
| 2020-06-01 | 0.218538 |
| 2020-07-01 | 0.286222 |
| 2020-08-01 | 0.222233 |
| 2020-09-01 | 0.244795 |
| 2020-10-01 | 0.314523 |
| 2020-11-01 | 0.292595 |
| 2020-12-01 | 0.407166 |
| 2021-01-01 | 0.430338 |
| 2021-02-01 | 0.430338 |
| 2021-03-01 | 0.387605 |
| 2021-04-01 | 0.43434 |
| 2021-05-01 | 0.481075 |
| 2021-06-01 | 0.482218 |
| 2021-07-01 | 0.461711 |
| 2021-08-01 | 0.462738 |
| 2021-09-01 | 0.463766 |
| 2021-10-01 | 0.419094 |
| 2021-11-01 | 0.536557 |
| 2021-12-01 | 0.46897 |
| 2022-01-01 | 0.468828 |
| 2022-02-01 | 0.491858 |
| 2022-03-01 | 0.468686 |
| 2022-04-01 | 0.444677 |
| 2022-05-01 | 0.443395 |
| 2022-06-01 | 0.442754 |
| 2022-07-01 | 0.440079 |
| 2022-08-01 | 0.438742 |
| 2022-09-01 | 0.459656 |
| 2022-10-01 | 0.522736 |
| 2022-11-01 | 0.493326 |
| 2022-12-01 | 0.490367 |
| 2023-01-01 | 0.4888 |
| 2023-02-01 | 0.508695 |
| 2023-03-01 | 0.573081 |
| 2023-04-01 | 0.571577 |
| 2023-05-01 | 0.58979 |
| 2023-06-01 | 0.609507 |
| 2023-07-01 | 0.58582 |
| 2023-08-01 | 0.646831 |
| 2023-09-01 | 0.664775 |
| 2023-10-01 | 0.658709 |
| 2023-11-01 | 0.653856 |
| 2023-12-01 | 0.651429 |
| 2024-01-01 | 0.691554 |
| 2024-02-01 | 0.689687 |
| 2024-03-01 | 0.686887 |
| 2024-04-01 | 0.665665 |
| 2024-05-01 | 0.622571 |
| 2024-06-01 | 0.62127 |
| 2024-07-01 | 0.682049 |
| 2024-08-01 | 0.659983 |
| 2024-09-01 | 0.638333 |
| 2024-10-01 | 0.595736 |
| 2024-11-01 | 0.553472 |
| 2024-12-01 | 0.552807 |

|  | Births |
| --- | --- |
| 2015-07-01 | 305.907 |
| 2015-08-01 | 307.252 |
| 2015-09-01 | 306.931 |
| 2015-10-01 | 305.902 |
| 2015-11-01 | 305.05 |
| 2015-12-01 | 302.85 |
| 2016-01-01 | 302.118 |
| 2016-02-01 | 299.073 |
| 2016-03-01 | 297.453 |
| 2016-04-01 | 296.008 |
| 2016-05-01 | 293.041 |
| 2016-06-01 | 291.203 |
| 2016-07-01 | 287.67 |
| 2016-08-01 | 285.466 |
| 2016-09-01 | 283.211 |
| 2016-10-01 | 280.546 |
| 2016-11-01 | 280.648 |
| 2016-12-01 | 280.571 |
| 2017-01-01 | 277.775 |
| 2017-02-01 | 278.915 |
| 2017-03-01 | 278.083 |
| 2017-04-01 | 277.739 |
| 2017-05-01 | 275.402 |
| 2017-06-01 | 274.217 |
| 2017-07-01 | 273.025 |
| 2017-08-01 | 272.9 |
| 2017-09-01 | 271.342 |
| 2017-10-01 | 269.567 |
| 2017-11-01 | 265.275 |
| 2017-12-01 | 263.692 |
| 2018-01-01 | 263.774 |
| 2018-02-01 | 262.515 |
| 2018-03-01 | 263.531 |
| 2018-04-01 | 263.14 |
| 2018-05-01 | 263.981 |
| 2018-06-01 | 264.845 |
| 2018-07-01 | 264.498 |
| 2018-08-01 | 263.344 |
| 2018-09-01 | 262.098 |
| 2018-10-01 | 261.709 |
| 2018-11-01 | 262.307 |
| 2018-12-01 | 261.467 |
| 2019-01-01 | 259.712 |
| 2019-02-01 | 258.741 |
| 2019-03-01 | 256.927 |
| 2019-04-01 | 254.67 |
| 2019-05-01 | 254.364 |
| 2019-06-01 | 250.787 |
| 2019-07-01 | 249.894 |
| 2019-08-01 | 247.386 |
| 2019-09-01 | 246.06 |
| 2019-10-01 | 247.201 |
| 2019-11-01 | 246.642 |
| 2019-12-01 | 244.824 |
| 2020-01-01 | 244.187 |
| 2020-02-01 | 243.951 |
| 2020-03-01 | 244.056 |
| 2020-04-01 | 245.13 |
| 2020-05-01 | 244.275 |
| 2020-06-01 | 246.656 |
| 2020-07-01 | 246.722 |
| 2020-08-01 | 248.496 |
| 2020-09-01 | 251.148 |
| 2020-10-01 | 250.841 |
| 2020-11-01 | 251.206 |
| 2020-12-01 | 251.154 |
| 2021-01-01 | 252.951 |
| 2021-02-01 | 252.374 |
| 2021-03-01 | 250.254 |
| 2021-04-01 | 246.824 |
| 2021-05-01 | 246.139 |
| 2021-06-01 | 243.638 |
| 2021-07-01 | 243.644 |
| 2021-08-01 | 239.853 |
| 2021-09-01 | 236.398 |
| 2021-10-01 | 233.344 |
| 2021-11-01 | 230.511 |
| 2021-12-01 | 228.969 |
| 2022-01-01 | 224.739 |
| 2022-02-01 | 223.284 |
| 2022-03-01 | 220.536 |
| 2022-04-01 | 220.94 |
| 2022-05-01 | 217.245 |
| 2022-06-01 | 215.293 |
| 2022-07-01 | 210.694 |
| 2022-08-01 | 208.959 |
| 2022-09-01 | 206.811 |
| 2022-10-01 | 205.284 |
| 2022-11-01 | 203.18 |
| 2022-12-01 | 201.828 |
| 2023-01-01 | 200.725 |
| 2023-02-01 | 197.759 |
| 2023-03-01 | 196.753 |
| 2023-04-01 | 193.961 |
| 2023-05-01 | 193.193 |
| 2023-06-01 | 190.672 |
| 2023-07-01 | 190.146 |
| 2023-08-01 | 188.688 |
| 2023-09-01 | 186.53 |
| 2023-10-01 | 184.336 |
| 2023-11-01 | 182.568 |
| 2023-12-01 | 177.867 |
| 2024-01-01 | 174.137 |
| 2024-02-01 | 168.563 |
| 2024-03-01 | 155.905 |

Rolling average vasectomies and births per 100,000 male and female population respectively for 25-34 year olds from April 2016 to December 2024

Vasectomies per 100,000 male population

Births per 100,000 female population

| Month and year | Vasectomies |
| --- | --- |
| 2016-04-01 | 15.0437 |
| 2016-05-01 | 15.031 |
| 2016-06-01 | 15.0496 |
| 2016-07-01 | 14.8136 |
| 2016-08-01 | 14.7336 |
| 2016-09-01 | 14.8338 |
| 2016-10-01 | 14.8157 |
| 2016-11-01 | 15.1909 |
| 2016-12-01 | 15.1429 |
| 2017-01-01 | 15.0026 |
| 2017-02-01 | 15.0637 |
| 2017-03-01 | 14.7689 |
| 2017-04-01 | 14.5727 |
| 2017-05-01 | 14.5284 |
| 2017-06-01 | 14.7116 |
| 2017-07-01 | 14.7434 |
| 2017-08-01 | 14.9244 |
| 2017-09-01 | 15.05 |
| 2017-10-01 | 14.7969 |
| 2017-11-01 | 14.6235 |
| 2017-12-01 | 14.5825 |
| 2018-01-01 | 14.665 |
| 2018-02-01 | 14.6178 |
| 2018-03-01 | 14.5402 |
| 2018-04-01 | 14.8218 |
| 2018-05-01 | 14.9154 |
| 2018-06-01 | 14.5558 |
| 2018-07-01 | 14.6928 |
| 2018-08-01 | 14.9628 |
| 2018-09-01 | 14.9271 |
| 2018-10-01 | 15.331 |
| 2018-11-01 | 15.7861 |
| 2018-12-01 | 16.0194 |
| 2019-01-01 | 15.9235 |
| 2019-02-01 | 15.8826 |
| 2019-03-01 | 16.1602 |
| 2019-04-01 | 16.3283 |
| 2019-05-01 | 16.3696 |
| 2019-06-01 | 16.2882 |
| 2019-07-01 | 16.638 |
| 2019-08-01 | 16.4423 |
| 2019-09-01 | 16.6244 |
| 2019-10-01 | 16.8086 |
| 2019-11-01 | 16.3247 |
| 2019-12-01 | 16.0649 |
| 2020-01-01 | 16.0628 |
| 2020-02-01 | 16.1852 |
| 2020-03-01 | 15.8491 |
| 2020-04-01 | 15.0735 |
| 2020-05-01 | 14.5209 |
| 2020-06-01 | 15.4575 |
| 2020-07-01 | 15.9147 |
| 2020-08-01 | 16.4147 |
| 2020-09-01 | 16.6752 |
| 2020-10-01 | 16.9917 |
| 2020-11-01 | 17.3308 |
| 2020-12-01 | 17.8657 |
| 2021-01-01 | 18.1031 |
| 2021-02-01 | 18.4889 |
| 2021-03-01 | 18.5461 |
| 2021-04-01 | 19.1746 |
| 2021-05-01 | 19.7529 |
| 2021-06-01 | 19.39 |
| 2021-07-01 | 19.0366 |
| 2021-08-01 | 18.8145 |
| 2021-09-01 | 19.2618 |
| 2021-10-01 | 19.3806 |
| 2021-11-01 | 19.9984 |
| 2021-12-01 | 19.9849 |
| 2022-01-01 | 20.0764 |
| 2022-02-01 | 19.9265 |
| 2022-03-01 | 20.204 |
| 2022-04-01 | 20.4265 |
| 2022-05-01 | 20.7193 |
| 2022-06-01 | 20.7121 |
| 2022-07-01 | 20.6783 |
| 2022-08-01 | 21.1618 |
| 2022-09-01 | 22.1734 |
| 2022-10-01 | 22.6965 |
| 2022-11-01 | 22.7939 |
| 2022-12-01 | 23.34 |
| 2023-01-01 | 23.7212 |
| 2023-02-01 | 24.4142 |
| 2023-03-01 | 25.656 |
| 2023-04-01 | 26.1148 |
| 2023-05-01 | 26.6839 |
| 2023-06-01 | 27.5421 |
| 2023-07-01 | 28.059 |
| 2023-08-01 | 28.4352 |
| 2023-09-01 | 27.4776 |
| 2023-10-01 | 27.2096 |
| 2023-11-01 | 27.0962 |
| 2023-12-01 | 26.9868 |
| 2024-01-01 | 26.9417 |
| 2024-02-01 | 26.8541 |
| 2024-03-01 | 26.041 |
| 2024-04-01 | 26.1235 |
| 2024-05-01 | 26.1421 |
| 2024-06-01 | 25.7442 |
| 2024-07-01 | 25.7143 |
| 2024-08-01 | 25.6597 |
| 2024-09-01 | 25.5208 |
| 2024-10-01 | 25.5775 |
| 2024-11-01 | 25.5688 |
| 2024-12-01 | 25.3772 |

|  | Births |
| --- | --- |
| 2015-07-01 | 903.417 |
| 2015-08-01 | 905.152 |
| 2015-09-01 | 906.396 |
| 2015-10-01 | 904.319 |
| 2015-11-01 | 905.252 |
| 2015-12-01 | 903.317 |
| 2016-01-01 | 903.141 |
| 2016-02-01 | 903.662 |
| 2016-03-01 | 900.887 |
| 2016-04-01 | 902.371 |
| 2016-05-01 | 896.263 |
| 2016-06-01 | 892.056 |
| 2016-07-01 | 887.205 |
| 2016-08-01 | 881.806 |
| 2016-09-01 | 875.135 |
| 2016-10-01 | 872.039 |
| 2016-11-01 | 868.826 |
| 2016-12-01 | 864.686 |
| 2017-01-01 | 859.622 |
| 2017-02-01 | 855.131 |
| 2017-03-01 | 850.665 |
| 2017-04-01 | 849.442 |
| 2017-05-01 | 848.374 |
| 2017-06-01 | 846.209 |
| 2017-07-01 | 846.776 |
| 2017-08-01 | 846.729 |
| 2017-09-01 | 847.357 |
| 2017-10-01 | 847.579 |
| 2017-11-01 | 846.379 |
| 2017-12-01 | 844.429 |
| 2018-01-01 | 844.101 |
| 2018-02-01 | 844.523 |
| 2018-03-01 | 845.409 |
| 2018-04-01 | 841.429 |
| 2018-05-01 | 840.215 |
| 2018-06-01 | 838.919 |
| 2018-07-01 | 838.043 |
| 2018-08-01 | 838.724 |
| 2018-09-01 | 836.683 |
| 2018-10-01 | 833.537 |
| 2018-11-01 | 831.479 |
| 2018-12-01 | 830.805 |
| 2019-01-01 | 832.985 |
| 2019-02-01 | 831.346 |
| 2019-03-01 | 830.422 |
| 2019-04-01 | 830.517 |
| 2019-05-01 | 828.923 |
| 2019-06-01 | 826.04 |
| 2019-07-01 | 823.725 |
| 2019-08-01 | 820.5 |
| 2019-09-01 | 819.749 |
| 2019-10-01 | 820.492 |
| 2019-11-01 | 819.847 |
| 2019-12-01 | 819.668 |
| 2020-01-01 | 815.37 |
| 2020-02-01 | 814.467 |
| 2020-03-01 | 816.486 |
| 2020-04-01 | 821.847 |
| 2020-05-01 | 830.343 |
| 2020-06-01 | 843.251 |
| 2020-07-01 | 852.98 |
| 2020-08-01 | 861.095 |
| 2020-09-01 | 868.311 |
| 2020-10-01 | 873.1 |
| 2020-11-01 | 877.349 |
| 2020-12-01 | 881.253 |
| 2021-01-01 | 886.821 |
| 2021-02-01 | 891.261 |
| 2021-03-01 | 893.008 |
| 2021-04-01 | 889.676 |
| 2021-05-01 | 885.591 |
| 2021-06-01 | 880.624 |
| 2021-07-01 | 875.12 |
| 2021-08-01 | 868.527 |
| 2021-09-01 | 861.146 |
| 2021-10-01 | 851.94 |
| 2021-11-01 | 845.056 |
| 2021-12-01 | 836.89 |
| 2022-01-01 | 826.887 |
| 2022-02-01 | 821.76 |
| 2022-03-01 | 814.263 |
| 2022-04-01 | 804.59 |
| 2022-05-01 | 794.608 |
| 2022-06-01 | 782.161 |
| 2022-07-01 | 772.199 |
| 2022-08-01 | 767.752 |
| 2022-09-01 | 763.604 |
| 2022-10-01 | 760.547 |
| 2022-11-01 | 757.768 |
| 2022-12-01 | 754.745 |
| 2023-01-01 | 752.477 |
| 2023-02-01 | 747.691 |
| 2023-03-01 | 740.962 |
| 2023-04-01 | 738.674 |
| 2023-05-01 | 739.121 |
| 2023-06-01 | 734.197 |
| 2023-07-01 | 733.032 |
| 2023-08-01 | 728.558 |
| 2023-09-01 | 722.358 |
| 2023-10-01 | 720.651 |
| 2023-11-01 | 715.645 |
| 2023-12-01 | 709.829 |
| 2024-01-01 | 704.649 |
| 2024-02-01 | 686.7 |
| 2024-03-01 | 644.595 |

Rolling average vasectomies and births per 100,000 male and female population respectively for 35-44 year olds from April 2016 to December 2024

Vasectomies per 100,000 male population

Births per 100,000 female population

| Month and year | Vasectomies |
| --- | --- |
| 2016-04-01 | 53.5407 |
| 2016-05-01 | 54.4421 |
| 2016-06-01 | 54.885 |
| 2016-07-01 | 54.6879 |
| 2016-08-01 | 55.622 |
| 2016-09-01 | 55.7344 |
| 2016-10-01 | 55.9368 |
| 2016-11-01 | 56.9793 |
| 2016-12-01 | 56.4519 |
| 2017-01-01 | 56.614 |
| 2017-02-01 | 57.2614 |
| 2017-03-01 | 57.1459 |
| 2017-04-01 | 57.248 |
| 2017-05-01 | 56.8232 |
| 2017-06-01 | 57.5413 |
| 2017-07-01 | 57.9053 |
| 2017-08-01 | 57.5988 |
| 2017-09-01 | 58.3302 |
| 2017-10-01 | 58.8475 |
| 2017-11-01 | 59.0532 |
| 2017-12-01 | 58.8092 |
| 2018-01-01 | 59.6858 |
| 2018-02-01 | 58.3778 |
| 2018-03-01 | 58.7541 |
| 2018-04-01 | 59.8692 |
| 2018-05-01 | 60.4045 |
| 2018-06-01 | 59.4877 |
| 2018-07-01 | 59.8976 |
| 2018-08-01 | 60.4098 |
| 2018-09-01 | 60.5307 |
| 2018-10-01 | 61.1982 |
| 2018-11-01 | 61.357 |
| 2018-12-01 | 61.9451 |
| 2019-01-01 | 62.545 |
| 2019-02-01 | 63.6501 |
| 2019-03-01 | 63.7934 |
| 2019-04-01 | 63.769 |
| 2019-05-01 | 64.2268 |
| 2019-06-01 | 64.2712 |
| 2019-07-01 | 65.2679 |
| 2019-08-01 | 65.5548 |
| 2019-09-01 | 65.9128 |
| 2019-10-01 | 65.8517 |
| 2019-11-01 | 65.5754 |
| 2019-12-01 | 66.1826 |
| 2020-01-01 | 65.5157 |
| 2020-02-01 | 65.338 |
| 2020-03-01 | 64.5489 |
| 2020-04-01 | 61.3444 |
| 2020-05-01 | 58.0989 |
| 2020-06-01 | 58.6024 |
| 2020-07-01 | 59.6178 |
| 2020-08-01 | 60.1582 |
| 2020-09-01 | 61.4879 |
| 2020-10-01 | 61.6849 |
| 2020-11-01 | 62.6952 |
| 2020-12-01 | 62.8989 |
| 2021-01-01 | 64.0053 |
| 2021-02-01 | 65.0729 |
| 2021-03-01 | 66.8096 |
| 2021-04-01 | 70.4805 |
| 2021-05-01 | 73.5677 |
| 2021-06-01 | 74.8004 |
| 2021-07-01 | 74.6412 |
| 2021-08-01 | 73.8386 |
| 2021-09-01 | 75.2693 |
| 2021-10-01 | 75.0654 |
| 2021-11-01 | 75.4868 |
| 2021-12-01 | 74.9585 |
| 2022-01-01 | 73.9647 |
| 2022-02-01 | 73.5155 |
| 2022-03-01 | 72.696 |
| 2022-04-01 | 71.8904 |
| 2022-05-01 | 72.4421 |
| 2022-06-01 | 71.4588 |
| 2022-07-01 | 69.9634 |
| 2022-08-01 | 70.9656 |
| 2022-09-01 | 71.2396 |
| 2022-10-01 | 72.6755 |
| 2022-11-01 | 73.5863 |
| 2022-12-01 | 74.6716 |
| 2023-01-01 | 75.3124 |
| 2023-02-01 | 77.2981 |
| 2023-03-01 | 81.1475 |
| 2023-04-01 | 82.9459 |
| 2023-05-01 | 84.4028 |
| 2023-06-01 | 86.6181 |
| 2023-07-01 | 87.71 |
| 2023-08-01 | 90.2308 |
| 2023-09-01 | 90.0365 |
| 2023-10-01 | 90.7191 |
| 2023-11-01 | 91.4849 |
| 2023-12-01 | 91.7855 |
| 2024-01-01 | 91.8649 |
| 2024-02-01 | 92.8709 |
| 2024-03-01 | 90.2181 |
| 2024-04-01 | 90.0954 |
| 2024-05-01 | 90.0893 |
| 2024-06-01 | 88.8211 |
| 2024-07-01 | 88.4253 |
| 2024-08-01 | 86.5743 |
| 2024-09-01 | 84.5465 |
| 2024-10-01 | 83.9388 |
| 2024-11-01 | 81.3958 |
| 2024-12-01 | 80.6648 |

|  | Births |
| --- | --- |
| 2015-07-01 | 300.271 |
| 2015-08-01 | 300.27 |
| 2015-09-01 | 301.137 |
| 2015-10-01 | 302.262 |
| 2015-11-01 | 302.373 |
| 2015-12-01 | 303.036 |
| 2016-01-01 | 302.751 |
| 2016-02-01 | 303.157 |
| 2016-03-01 | 301.799 |
| 2016-04-01 | 302.978 |
| 2016-05-01 | 302.921 |
| 2016-06-01 | 303.69 |
| 2016-07-01 | 302.616 |
| 2016-08-01 | 303.247 |
| 2016-09-01 | 304.03 |
| 2016-10-01 | 304.203 |
| 2016-11-01 | 303.83 |
| 2016-12-01 | 304.251 |
| 2017-01-01 | 304.776 |
| 2017-02-01 | 304.666 |
| 2017-03-01 | 303.238 |
| 2017-04-01 | 303.256 |
| 2017-05-01 | 303.16 |
| 2017-06-01 | 301.555 |
| 2017-07-01 | 301.569 |
| 2017-08-01 | 301.231 |
| 2017-09-01 | 299.322 |
| 2017-10-01 | 299.319 |
| 2017-11-01 | 299.315 |
| 2017-12-01 | 299.04 |
| 2018-01-01 | 300.493 |
| 2018-02-01 | 300.237 |
| 2018-03-01 | 301.482 |
| 2018-04-01 | 300.555 |
| 2018-05-01 | 301.53 |
| 2018-06-01 | 301.293 |
| 2018-07-01 | 303.195 |
| 2018-08-01 | 304.89 |
| 2018-09-01 | 304.549 |
| 2018-10-01 | 305.562 |
| 2018-11-01 | 304.647 |
| 2018-12-01 | 304.244 |
| 2019-01-01 | 304.437 |
| 2019-02-01 | 304.27 |
| 2019-03-01 | 305.328 |
| 2019-04-01 | 306.336 |
| 2019-05-01 | 305.585 |
| 2019-06-01 | 305.275 |
| 2019-07-01 | 304.293 |
| 2019-08-01 | 302.468 |
| 2019-09-01 | 303.275 |
| 2019-10-01 | 302.611 |
| 2019-11-01 | 302.264 |
| 2019-12-01 | 302.457 |
| 2020-01-01 | 300.036 |
| 2020-02-01 | 298.877 |
| 2020-03-01 | 297.289 |
| 2020-04-01 | 295.254 |
| 2020-05-01 | 297.374 |
| 2020-06-01 | 301.627 |
| 2020-07-01 | 304.333 |
| 2020-08-01 | 306.524 |
| 2020-09-01 | 309.748 |
| 2020-10-01 | 311.033 |
| 2020-11-01 | 313.851 |
| 2020-12-01 | 315.713 |
| 2021-01-01 | 319.425 |
| 2021-02-01 | 322.636 |
| 2021-03-01 | 325.58 |
| 2021-04-01 | 326.252 |
| 2021-05-01 | 324.63 |
| 2021-06-01 | 323.069 |
| 2021-07-01 | 321.228 |
| 2021-08-01 | 319.876 |
| 2021-09-01 | 317.151 |
| 2021-10-01 | 315.408 |
| 2021-11-01 | 313.373 |
| 2021-12-01 | 311.05 |
| 2022-01-01 | 306.819 |
| 2022-02-01 | 304.123 |
| 2022-03-01 | 301.027 |
| 2022-04-01 | 299.979 |
| 2022-05-01 | 296.934 |
| 2022-06-01 | 294.081 |
| 2022-07-01 | 290.772 |
| 2022-08-01 | 290.781 |
| 2022-09-01 | 289.951 |
| 2022-10-01 | 288.367 |
| 2022-11-01 | 287.261 |
| 2022-12-01 | 286.473 |
| 2023-01-01 | 287.319 |
| 2023-02-01 | 287.973 |
| 2023-03-01 | 286.648 |
| 2023-04-01 | 287.267 |
| 2023-05-01 | 289.213 |
| 2023-06-01 | 288.365 |
| 2023-07-01 | 290.704 |
| 2023-08-01 | 289.007 |
| 2023-09-01 | 286.873 |
| 2023-10-01 | 286.678 |
| 2023-11-01 | 285.476 |
| 2023-12-01 | 282.508 |
| 2024-01-01 | 280.143 |
| 2024-02-01 | 272.458 |
| 2024-03-01 | 254.931 |

Rolling average vasectomies and births per 100,000 male and female population respectively for all included from April 2016 to December 2024

Vasectomies per 100,000 male population

Births per 100,000 female population

| Month and year | Vasectomies |
| --- | --- |
| 2016-04-01 | 24.9134 |
| 2016-05-01 | 25.2262 |
| 2016-06-01 | 25.3894 |
| 2016-07-01 | 25.2302 |
| 2016-08-01 | 25.52 |
| 2016-09-01 | 25.6025 |
| 2016-10-01 | 25.6657 |
| 2016-11-01 | 26.1741 |
| 2016-12-01 | 25.9671 |
| 2017-01-01 | 25.9746 |
| 2017-02-01 | 26.2111 |
| 2017-03-01 | 26.0447 |
| 2017-04-01 | 25.9962 |
| 2017-05-01 | 25.826 |
| 2017-06-01 | 26.1367 |
| 2017-07-01 | 26.2645 |
| 2017-08-01 | 26.2144 |
| 2017-09-01 | 26.503 |
| 2017-10-01 | 26.5746 |
| 2017-11-01 | 26.5677 |
| 2017-12-01 | 26.4506 |
| 2018-01-01 | 26.7826 |
| 2018-02-01 | 26.3021 |
| 2018-03-01 | 26.4055 |
| 2018-04-01 | 26.9045 |
| 2018-05-01 | 27.1309 |
| 2018-06-01 | 26.6744 |
| 2018-07-01 | 26.8675 |
| 2018-08-01 | 27.1482 |
| 2018-09-01 | 27.1714 |
| 2018-10-01 | 27.5663 |
| 2018-11-01 | 27.819 |
| 2018-12-01 | 28.1137 |
| 2019-01-01 | 28.2844 |
| 2019-02-01 | 28.6593 |
| 2019-03-01 | 28.8226 |
| 2019-04-01 | 28.8826 |
| 2019-05-01 | 29.0508 |
| 2019-06-01 | 29.0252 |
| 2019-07-01 | 29.5201 |
| 2019-08-01 | 29.5787 |
| 2019-09-01 | 29.7828 |
| 2019-10-01 | 29.8568 |
| 2019-11-01 | 29.5736 |
| 2019-12-01 | 29.7045 |
| 2020-01-01 | 29.488 |
| 2020-02-01 | 29.4959 |
| 2020-03-01 | 29.1198 |
| 2020-04-01 | 27.6974 |
| 2020-05-01 | 26.3526 |
| 2020-06-01 | 26.9414 |
| 2020-07-01 | 27.5578 |
| 2020-08-01 | 27.9865 |
| 2020-09-01 | 28.623 |
| 2020-10-01 | 28.8891 |
| 2020-11-01 | 29.4357 |
| 2020-12-01 | 29.7971 |
| 2021-01-01 | 30.3464 |
| 2021-02-01 | 30.9414 |
| 2021-03-01 | 31.6426 |
| 2021-04-01 | 33.2762 |
| 2021-05-01 | 34.6804 |
| 2021-06-01 | 35.0471 |
| 2021-07-01 | 34.9134 |
| 2021-08-01 | 34.5922 |
| 2021-09-01 | 35.3655 |
| 2021-10-01 | 35.3648 |
| 2021-11-01 | 35.8351 |
| 2021-12-01 | 35.6534 |
| 2022-01-01 | 35.3357 |
| 2022-02-01 | 35.1327 |
| 2022-03-01 | 34.9513 |
| 2022-04-01 | 34.7328 |
| 2022-05-01 | 35.0481 |
| 2022-06-01 | 34.6793 |
| 2022-07-01 | 34.0838 |
| 2022-08-01 | 34.6109 |
| 2022-09-01 | 35.0693 |
| 2022-10-01 | 35.781 |
| 2022-11-01 | 36.1065 |
| 2022-12-01 | 36.687 |
| 2023-01-01 | 37.0405 |
| 2023-02-01 | 38.0028 |
| 2023-03-01 | 39.8658 |
| 2023-04-01 | 40.6677 |
| 2023-05-01 | 41.3851 |
| 2023-06-01 | 42.4946 |
| 2023-07-01 | 43.0535 |
| 2023-08-01 | 44.0887 |
| 2023-09-01 | 43.6038 |
| 2023-10-01 | 43.7155 |
| 2023-11-01 | 43.9148 |
| 2023-12-01 | 43.9552 |
| 2024-01-01 | 43.9633 |
| 2024-02-01 | 44.2723 |
| 2024-03-01 | 42.9707 |
| 2024-04-01 | 42.9467 |
| 2024-05-01 | 42.9338 |
| 2024-06-01 | 42.312 |
| 2024-07-01 | 42.1741 |
| 2024-08-01 | 41.4792 |
| 2024-09-01 | 40.6875 |
| 2024-10-01 | 40.4846 |
| 2024-11-01 | 39.5535 |
| 2024-12-01 | 39.2185 |

|  | Births |
| --- | --- |
| 2015-07-01 | 539.784 |
| 2015-08-01 | 540.942 |
| 2015-09-01 | 541.786 |
| 2015-10-01 | 541.356 |
| 2015-11-01 | 541.788 |
| 2015-12-01 | 540.971 |
| 2016-01-01 | 540.837 |
| 2016-02-01 | 540.646 |
| 2016-03-01 | 538.862 |
| 2016-04-01 | 539.698 |
| 2016-05-01 | 536.696 |
| 2016-06-01 | 535.036 |
| 2016-07-01 | 531.995 |
| 2016-08-01 | 529.674 |
| 2016-09-01 | 526.884 |
| 2016-10-01 | 525.16 |
| 2016-11-01 | 523.873 |
| 2016-12-01 | 522.457 |
| 2017-01-01 | 520.003 |
| 2017-02-01 | 518.506 |
| 2017-03-01 | 516.066 |
| 2017-04-01 | 515.515 |
| 2017-05-01 | 514.498 |
| 2017-06-01 | 512.788 |
| 2017-07-01 | 512.699 |
| 2017-08-01 | 512.499 |
| 2017-09-01 | 511.66 |
| 2017-10-01 | 511.283 |
| 2017-11-01 | 509.724 |
| 2017-12-01 | 508.429 |
| 2018-01-01 | 508.808 |
| 2018-02-01 | 508.551 |
| 2018-03-01 | 509.567 |
| 2018-04-01 | 507.531 |
| 2018-05-01 | 507.582 |
| 2018-06-01 | 507.169 |
| 2018-07-01 | 507.416 |
| 2018-08-01 | 508.013 |
| 2018-09-01 | 506.775 |
| 2018-10-01 | 505.813 |
| 2018-11-01 | 504.831 |
| 2018-12-01 | 504.23 |
| 2019-01-01 | 504.823 |
| 2019-02-01 | 503.934 |
| 2019-03-01 | 503.556 |
| 2019-04-01 | 503.472 |
| 2019-05-01 | 502.537 |
| 2019-06-01 | 500.463 |
| 2019-07-01 | 498.992 |
| 2019-08-01 | 496.467 |
| 2019-09-01 | 496.175 |
| 2019-10-01 | 496.498 |
| 2019-11-01 | 495.969 |
| 2019-12-01 | 495.525 |
| 2020-01-01 | 492.715 |
| 2020-02-01 | 491.808 |
| 2020-03-01 | 491.975 |
| 2020-04-01 | 493.519 |
| 2020-05-01 | 497.417 |
| 2020-06-01 | 504.604 |
| 2020-07-01 | 509.4 |
| 2020-08-01 | 513.744 |
| 2020-09-01 | 518.34 |
| 2020-10-01 | 520.534 |
| 2020-11-01 | 523.245 |
| 2020-12-01 | 525.365 |
| 2021-01-01 | 529.252 |
| 2021-02-01 | 531.97 |
| 2021-03-01 | 533.16 |
| 2021-04-01 | 531.125 |
| 2021-05-01 | 528.56 |
| 2021-06-01 | 525.236 |
| 2021-07-01 | 522.178 |
| 2021-08-01 | 518.018 |
| 2021-09-01 | 513.111 |
| 2021-10-01 | 507.952 |
| 2021-11-01 | 503.644 |
| 2021-12-01 | 499.015 |
| 2022-01-01 | 492.397 |
| 2022-02-01 | 488.913 |
| 2022-03-01 | 484.054 |
| 2022-04-01 | 479.896 |
| 2022-05-01 | 473.927 |
| 2022-06-01 | 467.451 |
| 2022-07-01 | 461.238 |
| 2022-08-01 | 459.09 |
| 2022-09-01 | 456.646 |
| 2022-10-01 | 454.519 |
| 2022-11-01 | 452.549 |
| 2022-12-01 | 450.781 |
| 2023-01-01 | 449.973 |
| 2023-02-01 | 447.676 |
| 2023-03-01 | 444.321 |
| 2023-04-01 | 443.022 |
| 2023-05-01 | 443.765 |
| 2023-06-01 | 440.943 |
| 2023-07-01 | 441.236 |
| 2023-08-01 | 438.494 |
| 2023-09-01 | 434.749 |
| 2023-10-01 | 433.458 |
| 2023-11-01 | 430.594 |
| 2023-12-01 | 426.069 |
| 2024-01-01 | 422.231 |
| 2024-02-01 | 410.96 |
| 2024-03-01 | 384.876 |

Victoria

Rolling average vasectomies and births per 100,000 male and female population respectively for 18-24 year olds from April 2016 to December 2024

Vasectomies per 100,000 male population

Births per 100,000 female population

| Month and year | Vasectomies |
| --- | --- |
| 2016-04-01 | 0.389778 |
| 2016-05-01 | 0.383611 |
| 2016-06-01 | 0.351643 |
| 2016-07-01 | 0.32366 |
| 2016-08-01 | 0.403636 |
| 2016-09-01 | 0.429633 |
| 2016-10-01 | 0.34794 |
| 2016-11-01 | 0.34794 |
| 2016-12-01 | 0.34794 |
| 2017-01-01 | 0.321064 |
| 2017-02-01 | 0.294188 |
| 2017-03-01 | 0.292837 |
| 2017-04-01 | 0.239181 |
| 2017-05-01 | 0.212353 |
| 2017-06-01 | 0.238581 |
| 2017-07-01 | 0.211922 |
| 2017-08-01 | 0.131946 |
| 2017-09-01 | 0.0786291 |
| 2017-10-01 | 0.130867 |
| 2017-11-01 | 0.156985 |
| 2017-12-01 | 0.156985 |
| 2018-01-01 | 0.156985 |
| 2018-02-01 | 0.182681 |
| 2018-03-01 | 0.181672 |
| 2018-04-01 | 0.207334 |
| 2018-05-01 | 0.28432 |
| 2018-06-01 | 0.309416 |
| 2018-07-01 | 0.309416 |
| 2018-08-01 | 0.385863 |
| 2018-09-01 | 0.411345 |
| 2018-10-01 | 0.38465 |
| 2018-11-01 | 0.358532 |
| 2018-12-01 | 0.409617 |
| 2019-01-01 | 0.459941 |
| 2019-02-01 | 0.484569 |
| 2019-03-01 | 0.458339 |
| 2019-04-01 | 0.432677 |
| 2019-05-01 | 0.431634 |
| 2019-06-01 | 0.405624 |
| 2019-07-01 | 0.405624 |
| 2019-08-01 | 0.354481 |
| 2019-09-01 | 0.328999 |
| 2019-10-01 | 0.303456 |
| 2019-11-01 | 0.354874 |
| 2019-12-01 | 0.303789 |
| 2020-01-01 | 0.279196 |
| 2020-02-01 | 0.228872 |
| 2020-03-01 | 0.306633 |
| 2020-04-01 | 0.306633 |
| 2020-05-01 | 0.335207 |
| 2020-06-01 | 0.362151 |
| 2020-07-01 | 0.362151 |
| 2020-08-01 | 0.39051 |
| 2020-09-01 | 0.417342 |
| 2020-10-01 | 0.472507 |
| 2020-11-01 | 0.503837 |
| 2020-12-01 | 0.559003 |
| 2021-01-01 | 0.561329 |
| 2021-02-01 | 0.589385 |
| 2021-03-01 | 0.514519 |
| 2021-04-01 | 0.628297 |
| 2021-05-01 | 0.552224 |
| 2021-06-01 | 0.585299 |
| 2021-07-01 | 0.642921 |
| 2021-08-01 | 0.704503 |
| 2021-09-01 | 0.735294 |
| 2021-10-01 | 0.795306 |
| 2021-11-01 | 0.798941 |
| 2021-12-01 | 0.743775 |
| 2022-01-01 | 0.743927 |
| 2022-02-01 | 0.744079 |
| 2022-03-01 | 0.800646 |
| 2022-04-01 | 0.799504 |
| 2022-05-01 | 0.883695 |
| 2022-06-01 | 0.882839 |
| 2022-07-01 | 0.908106 |
| 2022-08-01 | 0.903381 |
| 2022-09-01 | 0.873389 |
| 2022-10-01 | 1.00436 |
| 2022-11-01 | 1.05472 |
| 2022-12-01 | 1.19147 |
| 2023-01-01 | 1.21653 |
| 2023-02-01 | 1.26822 |
| 2023-03-01 | 1.26349 |
| 2023-04-01 | 1.15086 |
| 2023-05-01 | 1.09096 |
| 2023-06-01 | 1.00648 |
| 2023-07-01 | 0.975393 |
| 2023-08-01 | 0.864874 |
| 2023-09-01 | 0.966748 |
| 2023-10-01 | 0.798041 |
| 2023-11-01 | 0.790359 |
| 2023-12-01 | 0.80849 |
| 2024-01-01 | 0.755224 |
| 2024-02-01 | 0.700713 |
| 2024-03-01 | 0.671589 |
| 2024-04-01 | 0.747766 |
| 2024-05-01 | 0.720422 |
| 2024-06-01 | 0.796599 |
| 2024-07-01 | 0.770006 |
| 2024-08-01 | 0.820424 |
| 2024-09-01 | 0.816964 |
| 2024-10-01 | 0.739525 |
| 2024-11-01 | 0.6357 |
| 2024-12-01 | 0.506061 |

|  | Births |
| --- | --- |
| 2015-07-01 | 239.08 |
| 2015-08-01 | 237.865 |
| 2015-09-01 | 236.853 |
| 2015-10-01 | 234.512 |
| 2015-11-01 | 231.557 |
| 2015-12-01 | 230.931 |
| 2016-01-01 | 229.139 |
| 2016-02-01 | 227.567 |
| 2016-03-01 | 226.701 |
| 2016-04-01 | 222.431 |
| 2016-05-01 | 219.312 |
| 2016-06-01 | 216.532 |
| 2016-07-01 | 214.974 |
| 2016-08-01 | 213.647 |
| 2016-09-01 | 211.468 |
| 2016-10-01 | 211.07 |
| 2016-11-01 | 210.435 |
| 2016-12-01 | 209.007 |
| 2017-01-01 | 208.462 |
| 2017-02-01 | 207.279 |
| 2017-03-01 | 205.876 |
| 2017-04-01 | 205.75 |
| 2017-05-01 | 204.859 |
| 2017-06-01 | 204.356 |
| 2017-07-01 | 203.984 |
| 2017-08-01 | 202.458 |
| 2017-09-01 | 200.889 |
| 2017-10-01 | 198.457 |
| 2017-11-01 | 196.977 |
| 2017-12-01 | 195.209 |
| 2018-01-01 | 193.694 |
| 2018-02-01 | 193.767 |
| 2018-03-01 | 192.309 |
| 2018-04-01 | 191.147 |
| 2018-05-01 | 190.378 |
| 2018-06-01 | 189.193 |
| 2018-07-01 | 188.591 |
| 2018-08-01 | 189.37 |
| 2018-09-01 | 189.607 |
| 2018-10-01 | 190.53 |
| 2018-11-01 | 187.876 |
| 2018-12-01 | 186.084 |
| 2019-01-01 | 185.892 |
| 2019-02-01 | 185.005 |
| 2019-03-01 | 182.708 |
| 2019-04-01 | 181.257 |
| 2019-05-01 | 181.412 |
| 2019-06-01 | 181.052 |
| 2019-07-01 | 179.118 |
| 2019-08-01 | 177.88 |
| 2019-09-01 | 177.066 |
| 2019-10-01 | 174.024 |
| 2019-11-01 | 174.462 |
| 2019-12-01 | 175.349 |
| 2020-01-01 | 174.233 |
| 2020-02-01 | 172.818 |
| 2020-03-01 | 174.285 |
| 2020-04-01 | 174.916 |
| 2020-05-01 | 174.155 |
| 2020-06-01 | 174.382 |
| 2020-07-01 | 175.605 |
| 2020-08-01 | 174.234 |
| 2020-09-01 | 173.63 |
| 2020-10-01 | 174.08 |
| 2020-11-01 | 172.82 |
| 2020-12-01 | 171.67 |
| 2021-01-01 | 171.547 |
| 2021-02-01 | 170.819 |
| 2021-03-01 | 169.011 |
| 2021-04-01 | 166.892 |
| 2021-05-01 | 165.373 |
| 2021-06-01 | 163.616 |
| 2021-07-01 | 161.226 |
| 2021-08-01 | 159.712 |
| 2021-09-01 | 158.861 |
| 2021-10-01 | 156.399 |
| 2021-11-01 | 156.148 |
| 2021-12-01 | 155.794 |
| 2022-01-01 | 155.118 |
| 2022-02-01 | 154.656 |
| 2022-03-01 | 153.858 |
| 2022-04-01 | 152.697 |
| 2022-05-01 | 150.541 |
| 2022-06-01 | 148.444 |
| 2022-07-01 | 148.267 |
| 2022-08-01 | 148.322 |
| 2022-09-01 | 147.339 |
| 2022-10-01 | 146.703 |
| 2022-11-01 | 144.977 |
| 2022-12-01 | 142.876 |
| 2023-01-01 | 140.716 |
| 2023-02-01 | 138.421 |
| 2023-03-01 | 137.449 |
| 2023-04-01 | 135.273 |
| 2023-05-01 | 135.807 |
| 2023-06-01 | 134.002 |
| 2023-07-01 | 132.622 |
| 2023-08-01 | 129.894 |
| 2023-09-01 | 127.732 |
| 2023-10-01 | 127.476 |
| 2023-11-01 | 126.324 |
| 2023-12-01 | 124.773 |
| 2024-01-01 | 122.503 |
| 2024-02-01 | 119.287 |
| 2024-03-01 | 108.48 |

Rolling average vasectomies and births per 100,000 male and female population respectively for 25-34 year olds from April 2016 to December 2024

Vasectomies per 100,000 male population

Births per 100,000 female population

| Month and year | Vasectomies |
| --- | --- |
| 2016-04-01 | 13.4666 |
| 2016-05-01 | 13.7667 |
| 2016-06-01 | 13.7374 |
| 2016-07-01 | 13.5042 |
| 2016-08-01 | 13.8997 |
| 2016-09-01 | 13.8129 |
| 2016-10-01 | 14.142 |
| 2016-11-01 | 13.8388 |
| 2016-12-01 | 13.8342 |
| 2017-01-01 | 13.8445 |
| 2017-02-01 | 13.9535 |
| 2017-03-01 | 14.1804 |
| 2017-04-01 | 14.1224 |
| 2017-05-01 | 13.7606 |
| 2017-06-01 | 13.9668 |
| 2017-07-01 | 13.9854 |
| 2017-08-01 | 13.8382 |
| 2017-09-01 | 14.2986 |
| 2017-10-01 | 14.5125 |
| 2017-11-01 | 14.6493 |
| 2017-12-01 | 14.8836 |
| 2018-01-01 | 14.947 |
| 2018-02-01 | 15.2047 |
| 2018-03-01 | 15.0586 |
| 2018-04-01 | 15.1051 |
| 2018-05-01 | 15.2617 |
| 2018-06-01 | 15.1131 |
| 2018-07-01 | 15.0658 |
| 2018-08-01 | 15.3384 |
| 2018-09-01 | 15.1507 |
| 2018-10-01 | 14.8296 |
| 2018-11-01 | 15.2368 |
| 2018-12-01 | 15.2622 |
| 2019-01-01 | 15.0938 |
| 2019-02-01 | 14.9956 |
| 2019-03-01 | 15.3738 |
| 2019-04-01 | 16.0153 |
| 2019-05-01 | 16.4223 |
| 2019-06-01 | 16.2804 |
| 2019-07-01 | 16.4317 |
| 2019-08-01 | 16.4074 |
| 2019-09-01 | 16.5864 |
| 2019-10-01 | 17.0235 |
| 2019-11-01 | 17.1322 |
| 2019-12-01 | 16.7798 |
| 2020-01-01 | 17.3934 |
| 2020-02-01 | 17.5843 |
| 2020-03-01 | 17.2442 |
| 2020-04-01 | 15.9826 |
| 2020-05-01 | 15.6015 |
| 2020-06-01 | 15.8658 |
| 2020-07-01 | 16.59 |
| 2020-08-01 | 15.9407 |
| 2020-09-01 | 15.4512 |
| 2020-10-01 | 15.2636 |
| 2020-11-01 | 15.4546 |
| 2020-12-01 | 15.8767 |
| 2021-01-01 | 15.9397 |
| 2021-02-01 | 16.2865 |
| 2021-03-01 | 17.1362 |
| 2021-04-01 | 18.6814 |
| 2021-05-01 | 19.3832 |
| 2021-06-01 | 20.0217 |
| 2021-07-01 | 20.1489 |
| 2021-08-01 | 21.1508 |
| 2021-09-01 | 22.25 |
| 2021-10-01 | 23.3089 |
| 2021-11-01 | 24.0263 |
| 2021-12-01 | 23.8883 |
| 2022-01-01 | 24.0605 |
| 2022-02-01 | 24.1876 |
| 2022-03-01 | 24.1836 |
| 2022-04-01 | 24.1249 |
| 2022-05-01 | 24.3017 |
| 2022-06-01 | 24.6799 |
| 2022-07-01 | 24.6873 |
| 2022-08-01 | 24.5286 |
| 2022-09-01 | 25.0847 |
| 2022-10-01 | 25.9706 |
| 2022-11-01 | 25.5878 |
| 2022-12-01 | 26.9307 |
| 2023-01-01 | 27.3466 |
| 2023-02-01 | 27.4762 |
| 2023-03-01 | 27.992 |
| 2023-04-01 | 27.8062 |
| 2023-05-01 | 28.2779 |
| 2023-06-01 | 28.5316 |
| 2023-07-01 | 28.4197 |
| 2023-08-01 | 28.7924 |
| 2023-09-01 | 28.6566 |
| 2023-10-01 | 27.4558 |
| 2023-11-01 | 27.7379 |
| 2023-12-01 | 26.9754 |
| 2024-01-01 | 26.6551 |
| 2024-02-01 | 26.6626 |
| 2024-03-01 | 25.9497 |
| 2024-04-01 | 26.3918 |
| 2024-05-01 | 25.8686 |
| 2024-06-01 | 25.0176 |
| 2024-07-01 | 25.3366 |
| 2024-08-01 | 25.3072 |
| 2024-09-01 | 24.8221 |
| 2024-10-01 | 24.7732 |
| 2024-11-01 | 24.098 |
| 2024-12-01 | 23.8663 |

|  | Births |
| --- | --- |
| 2015-07-01 | 879.983 |
| 2015-08-01 | 879.486 |
| 2015-09-01 | 876.965 |
| 2015-10-01 | 873.36 |
| 2015-11-01 | 872.013 |
| 2015-12-01 | 869.372 |
| 2016-01-01 | 862.841 |
| 2016-02-01 | 862.379 |
| 2016-03-01 | 859.476 |
| 2016-04-01 | 858.212 |
| 2016-05-01 | 852.952 |
| 2016-06-01 | 848.81 |
| 2016-07-01 | 843.745 |
| 2016-08-01 | 841.031 |
| 2016-09-01 | 836.609 |
| 2016-10-01 | 831.86 |
| 2016-11-01 | 831.435 |
| 2016-12-01 | 827.164 |
| 2017-01-01 | 830.555 |
| 2017-02-01 | 826.491 |
| 2017-03-01 | 822.602 |
| 2017-04-01 | 822.099 |
| 2017-05-01 | 819.375 |
| 2017-06-01 | 817.017 |
| 2017-07-01 | 814.835 |
| 2017-08-01 | 814.179 |
| 2017-09-01 | 811.592 |
| 2017-10-01 | 812.079 |
| 2017-11-01 | 808.141 |
| 2017-12-01 | 802.157 |
| 2018-01-01 | 796.358 |
| 2018-02-01 | 795.334 |
| 2018-03-01 | 794.834 |
| 2018-04-01 | 790.096 |
| 2018-05-01 | 788.003 |
| 2018-06-01 | 786.043 |
| 2018-07-01 | 786.548 |
| 2018-08-01 | 785.248 |
| 2018-09-01 | 785.061 |
| 2018-10-01 | 783.637 |
| 2018-11-01 | 781.309 |
| 2018-12-01 | 785.428 |
| 2019-01-01 | 786.86 |
| 2019-02-01 | 782.788 |
| 2019-03-01 | 778.204 |
| 2019-04-01 | 778.821 |
| 2019-05-01 | 778.676 |
| 2019-06-01 | 778.02 |
| 2019-07-01 | 775.844 |
| 2019-08-01 | 772.068 |
| 2019-09-01 | 769.846 |
| 2019-10-01 | 769.275 |
| 2019-11-01 | 769 |
| 2019-12-01 | 766.453 |
| 2020-01-01 | 764.594 |
| 2020-02-01 | 764.765 |
| 2020-03-01 | 765.261 |
| 2020-04-01 | 768.43 |
| 2020-05-01 | 778.369 |
| 2020-06-01 | 787.513 |
| 2020-07-01 | 794.838 |
| 2020-08-01 | 802.794 |
| 2020-09-01 | 811.358 |
| 2020-10-01 | 817.777 |
| 2020-11-01 | 820.471 |
| 2020-12-01 | 823.562 |
| 2021-01-01 | 827.609 |
| 2021-02-01 | 832.435 |
| 2021-03-01 | 838.245 |
| 2021-04-01 | 833.801 |
| 2021-05-01 | 826.826 |
| 2021-06-01 | 819.815 |
| 2021-07-01 | 816.379 |
| 2021-08-01 | 813.098 |
| 2021-09-01 | 805.985 |
| 2021-10-01 | 796.821 |
| 2021-11-01 | 791.794 |
| 2021-12-01 | 787.453 |
| 2022-01-01 | 779.15 |
| 2022-02-01 | 773.973 |
| 2022-03-01 | 766.286 |
| 2022-04-01 | 762.264 |
| 2022-05-01 | 751.957 |
| 2022-06-01 | 743.219 |
| 2022-07-01 | 732.262 |
| 2022-08-01 | 722.854 |
| 2022-09-01 | 718.419 |
| 2022-10-01 | 716.212 |
| 2022-11-01 | 712.943 |
| 2022-12-01 | 709.693 |
| 2023-01-01 | 704.355 |
| 2023-02-01 | 700.505 |
| 2023-03-01 | 694.968 |
| 2023-04-01 | 691.052 |
| 2023-05-01 | 690.911 |
| 2023-06-01 | 689.035 |
| 2023-07-01 | 689.23 |
| 2023-08-01 | 687.876 |
| 2023-09-01 | 684.427 |
| 2023-10-01 | 683.178 |
| 2023-11-01 | 681.432 |
| 2023-12-01 | 676.924 |
| 2024-01-01 | 673.59 |
| 2024-02-01 | 656.869 |
| 2024-03-01 | 604.582 |

Rolling average vasectomies and births per 100,000 male and female population respectively for 35-44 year olds from April 2016 to December 2024

Vasectomies per 100,000 male population

Births per 100,000 female population

| Month and year | Vasectomies |
| --- | --- |
| 2016-04-01 | 68.6761 |
| 2016-05-01 | 68.6851 |
| 2016-06-01 | 69.0345 |
| 2016-07-01 | 69.0332 |
| 2016-08-01 | 69.5011 |
| 2016-09-01 | 70.3149 |
| 2016-10-01 | 70.3102 |
| 2016-11-01 | 70.2296 |
| 2016-12-01 | 69.4708 |
| 2017-01-01 | 69.4027 |
| 2017-02-01 | 69.2434 |
| 2017-03-01 | 70.4214 |
| 2017-04-01 | 69.1469 |
| 2017-05-01 | 69.2836 |
| 2017-06-01 | 70.007 |
| 2017-07-01 | 68.706 |
| 2017-08-01 | 69.2926 |
| 2017-09-01 | 68.6808 |
| 2017-10-01 | 68.3789 |
| 2017-11-01 | 68.5519 |
| 2017-12-01 | 68.9726 |
| 2018-01-01 | 69.3174 |
| 2018-02-01 | 69.431 |
| 2018-03-01 | 69.2735 |
| 2018-04-01 | 69.7599 |
| 2018-05-01 | 71.0168 |
| 2018-06-01 | 70.4763 |
| 2018-07-01 | 71.6613 |
| 2018-08-01 | 71.5568 |
| 2018-09-01 | 71.4611 |
| 2018-10-01 | 71.4147 |
| 2018-11-01 | 72.2982 |
| 2018-12-01 | 72.7722 |
| 2019-01-01 | 73.4689 |
| 2019-02-01 | 74.0544 |
| 2019-03-01 | 75.2319 |
| 2019-04-01 | 75.856 |
| 2019-05-01 | 77.0803 |
| 2019-06-01 | 77.1191 |
| 2019-07-01 | 77.0922 |
| 2019-08-01 | 77.7447 |
| 2019-09-01 | 78.6701 |
| 2019-10-01 | 79.5772 |
| 2019-11-01 | 80.3815 |
| 2019-12-01 | 79.9439 |
| 2020-01-01 | 81.642 |
| 2020-02-01 | 82.4917 |
| 2020-03-01 | 81.2802 |
| 2020-04-01 | 76.4428 |
| 2020-05-01 | 72.8701 |
| 2020-06-01 | 72.748 |
| 2020-07-01 | 74.2553 |
| 2020-08-01 | 70.5118 |
| 2020-09-01 | 67.3917 |
| 2020-10-01 | 67.1699 |
| 2020-11-01 | 65.4688 |
| 2020-12-01 | 66.2873 |
| 2021-01-01 | 65.8024 |
| 2021-02-01 | 66.719 |
| 2021-03-01 | 69.2518 |
| 2021-04-01 | 75.325 |
| 2021-05-01 | 78.2714 |
| 2021-06-01 | 79.8397 |
| 2021-07-01 | 81.2438 |
| 2021-08-01 | 86.5196 |
| 2021-09-01 | 91.3449 |
| 2021-10-01 | 92.5332 |
| 2021-11-01 | 94.4143 |
| 2021-12-01 | 93.4362 |
| 2022-01-01 | 92.1952 |
| 2022-02-01 | 92.1205 |
| 2022-03-01 | 91.3506 |
| 2022-04-01 | 90.3866 |
| 2022-05-01 | 91.3718 |
| 2022-06-01 | 92.1962 |
| 2022-07-01 | 89.8815 |
| 2022-08-01 | 88.313 |
| 2022-09-01 | 88.7066 |
| 2022-10-01 | 93.0216 |
| 2022-11-01 | 94.4456 |
| 2022-12-01 | 98.2709 |
| 2023-01-01 | 100.036 |
| 2023-02-01 | 101.085 |
| 2023-03-01 | 103.689 |
| 2023-04-01 | 102.934 |
| 2023-05-01 | 104.571 |
| 2023-06-01 | 104.603 |
| 2023-07-01 | 105.421 |
| 2023-08-01 | 107.574 |
| 2023-09-01 | 107.525 |
| 2023-10-01 | 102.935 |
| 2023-11-01 | 103.172 |
| 2023-12-01 | 100.785 |
| 2024-01-01 | 99.6166 |
| 2024-02-01 | 99.0976 |
| 2024-03-01 | 95.8854 |
| 2024-04-01 | 97.8081 |
| 2024-05-01 | 96.5927 |
| 2024-06-01 | 94.9536 |
| 2024-07-01 | 96.5226 |
| 2024-08-01 | 95.1537 |
| 2024-09-01 | 92.9243 |
| 2024-10-01 | 92.8015 |
| 2024-11-01 | 90.4924 |
| 2024-12-01 | 89.417 |

|  | Births |
| --- | --- |
| 2015-07-01 | 312.48 |
| 2015-08-01 | 313.034 |
| 2015-09-01 | 313.318 |
| 2015-10-01 | 313.463 |
| 2015-11-01 | 313.161 |
| 2015-12-01 | 312.875 |
| 2016-01-01 | 312.296 |
| 2016-02-01 | 311.51 |
| 2016-03-01 | 310.943 |
| 2016-04-01 | 311.814 |
| 2016-05-01 | 310.906 |
| 2016-06-01 | 309.844 |
| 2016-07-01 | 309.595 |
| 2016-08-01 | 310.193 |
| 2016-09-01 | 309.545 |
| 2016-10-01 | 308.166 |
| 2016-11-01 | 310.186 |
| 2016-12-01 | 307.869 |
| 2017-01-01 | 307.971 |
| 2017-02-01 | 310.188 |
| 2017-03-01 | 309.941 |
| 2017-04-01 | 309.461 |
| 2017-05-01 | 309.005 |
| 2017-06-01 | 309.303 |
| 2017-07-01 | 308.469 |
| 2017-08-01 | 307.464 |
| 2017-09-01 | 307.845 |
| 2017-10-01 | 310.133 |
| 2017-11-01 | 309.395 |
| 2017-12-01 | 311.058 |
| 2018-01-01 | 312.98 |
| 2018-02-01 | 311.997 |
| 2018-03-01 | 310.599 |
| 2018-04-01 | 311.726 |
| 2018-05-01 | 313.013 |
| 2018-06-01 | 312.597 |
| 2018-07-01 | 312.733 |
| 2018-08-01 | 313.577 |
| 2018-09-01 | 312.637 |
| 2018-10-01 | 311.763 |
| 2018-11-01 | 311.888 |
| 2018-12-01 | 311.017 |
| 2019-01-01 | 310.105 |
| 2019-02-01 | 310.183 |
| 2019-03-01 | 310.549 |
| 2019-04-01 | 310.212 |
| 2019-05-01 | 308.743 |
| 2019-06-01 | 308.435 |
| 2019-07-01 | 308.859 |
| 2019-08-01 | 307.307 |
| 2019-09-01 | 307.342 |
| 2019-10-01 | 306.715 |
| 2019-11-01 | 306.77 |
| 2019-12-01 | 307.167 |
| 2020-01-01 | 306.356 |
| 2020-02-01 | 303.928 |
| 2020-03-01 | 302.524 |
| 2020-04-01 | 301.259 |
| 2020-05-01 | 304.59 |
| 2020-06-01 | 308.659 |
| 2020-07-01 | 310.703 |
| 2020-08-01 | 314.426 |
| 2020-09-01 | 314.362 |
| 2020-10-01 | 316.648 |
| 2020-11-01 | 316.874 |
| 2020-12-01 | 319.906 |
| 2021-01-01 | 319.988 |
| 2021-02-01 | 322.562 |
| 2021-03-01 | 327.756 |
| 2021-04-01 | 330.426 |
| 2021-05-01 | 329.967 |
| 2021-06-01 | 327.158 |
| 2021-07-01 | 324.99 |
| 2021-08-01 | 322.238 |
| 2021-09-01 | 323.055 |
| 2021-10-01 | 321.295 |
| 2021-11-01 | 319.602 |
| 2021-12-01 | 316.885 |
| 2022-01-01 | 314.688 |
| 2022-02-01 | 313.654 |
| 2022-03-01 | 310.664 |
| 2022-04-01 | 307.715 |
| 2022-05-01 | 304.206 |
| 2022-06-01 | 303.093 |
| 2022-07-01 | 301.875 |
| 2022-08-01 | 300.509 |
| 2022-09-01 | 299.076 |
| 2022-10-01 | 297.629 |
| 2022-11-01 | 297.126 |
| 2022-12-01 | 294.734 |
| 2023-01-01 | 295.388 |
| 2023-02-01 | 295.973 |
| 2023-03-01 | 295.339 |
| 2023-04-01 | 294.489 |
| 2023-05-01 | 294.613 |
| 2023-06-01 | 292.486 |
| 2023-07-01 | 291.967 |
| 2023-08-01 | 291.848 |
| 2023-09-01 | 291.568 |
| 2023-10-01 | 291.824 |
| 2023-11-01 | 291.345 |
| 2023-12-01 | 289.143 |
| 2024-01-01 | 286.106 |
| 2024-02-01 | 275.237 |
| 2024-03-01 | 250.899 |

Rolling average vasectomies and births per 100,000 male and female population respectively for all included from April 2016 to December 2024

Vasectomies per 100,000 male population

Births per 100,000 female population

| Month and year | Vasectomies |
| --- | --- |
| 2016-04-01 | 29.1952 |
| 2016-05-01 | 29.309 |
| 2016-06-01 | 29.4038 |
| 2016-07-01 | 29.2891 |
| 2016-08-01 | 29.6147 |
| 2016-09-01 | 29.8546 |
| 2016-10-01 | 29.9509 |
| 2016-11-01 | 29.7921 |
| 2016-12-01 | 29.518 |
| 2017-01-01 | 29.4841 |
| 2017-02-01 | 29.4539 |
| 2017-03-01 | 29.9367 |
| 2017-04-01 | 29.4527 |
| 2017-05-01 | 29.3401 |
| 2017-06-01 | 29.6679 |
| 2017-07-01 | 29.2146 |
| 2017-08-01 | 29.3294 |
| 2017-09-01 | 29.2816 |
| 2017-10-01 | 29.2746 |
| 2017-11-01 | 29.393 |
| 2017-12-01 | 29.6294 |
| 2018-01-01 | 29.7726 |
| 2018-02-01 | 29.9206 |
| 2018-03-01 | 29.8077 |
| 2018-04-01 | 29.9992 |
| 2018-05-01 | 30.5118 |
| 2018-06-01 | 30.2733 |
| 2018-07-01 | 30.6597 |
| 2018-08-01 | 30.7518 |
| 2018-09-01 | 30.6502 |
| 2018-10-01 | 30.4992 |
| 2018-11-01 | 30.959 |
| 2018-12-01 | 31.1453 |
| 2019-01-01 | 31.33 |
| 2019-02-01 | 31.4983 |
| 2019-03-01 | 32.0464 |
| 2019-04-01 | 32.517 |
| 2019-05-01 | 33.1077 |
| 2019-06-01 | 33.0642 |
| 2019-07-01 | 33.1293 |
| 2019-08-01 | 33.3466 |
| 2019-09-01 | 33.7432 |
| 2019-10-01 | 34.2511 |
| 2019-11-01 | 34.6153 |
| 2019-12-01 | 34.3339 |
| 2020-01-01 | 35.1961 |
| 2020-02-01 | 35.5959 |
| 2020-03-01 | 35.1041 |
| 2020-04-01 | 32.9463 |
| 2020-05-01 | 31.6103 |
| 2020-06-01 | 31.7287 |
| 2020-07-01 | 32.625 |
| 2020-08-01 | 31.124 |
| 2020-09-01 | 29.8984 |
| 2020-10-01 | 29.8406 |
| 2020-11-01 | 29.4093 |
| 2020-12-01 | 29.9575 |
| 2021-01-01 | 29.9019 |
| 2021-02-01 | 30.4829 |
| 2021-03-01 | 31.814 |
| 2021-04-01 | 34.6991 |
| 2021-05-01 | 36.108 |
| 2021-06-01 | 37.0342 |
| 2021-07-01 | 37.719 |
| 2021-08-01 | 40.1285 |
| 2021-09-01 | 42.3998 |
| 2021-10-01 | 43.34 |
| 2021-11-01 | 44.3808 |
| 2021-12-01 | 44.0156 |
| 2022-01-01 | 43.6588 |
| 2022-02-01 | 43.7272 |
| 2022-03-01 | 43.5037 |
| 2022-04-01 | 43.15 |
| 2022-05-01 | 43.634 |
| 2022-06-01 | 44.1177 |
| 2022-07-01 | 43.2681 |
| 2022-08-01 | 42.6226 |
| 2022-09-01 | 42.9777 |
| 2022-10-01 | 44.9402 |
| 2022-11-01 | 45.3141 |
| 2022-12-01 | 47.2815 |
| 2023-01-01 | 48.0888 |
| 2023-02-01 | 48.5203 |
| 2023-03-01 | 49.6624 |
| 2023-04-01 | 49.2633 |
| 2023-05-01 | 50.0079 |
| 2023-06-01 | 50.0704 |
| 2023-07-01 | 50.2963 |
| 2023-08-01 | 51.1822 |
| 2023-09-01 | 51.1064 |
| 2023-10-01 | 48.8764 |
| 2023-11-01 | 49.0495 |
| 2023-12-01 | 47.8531 |
| 2024-01-01 | 47.2767 |
| 2024-02-01 | 47.0644 |
| 2024-03-01 | 45.5851 |
| 2024-04-01 | 46.4843 |
| 2024-05-01 | 45.8247 |
| 2024-06-01 | 44.9051 |
| 2024-07-01 | 45.6107 |
| 2024-08-01 | 45.1177 |
| 2024-09-01 | 44.1146 |
| 2024-10-01 | 44.0432 |
| 2024-11-01 | 42.9163 |
| 2024-12-01 | 42.408 |

|  | Births |
| --- | --- |
| 2015-07-01 | 520.874 |
| 2015-08-01 | 520.707 |
| 2015-09-01 | 519.659 |
| 2015-10-01 | 517.894 |
| 2015-11-01 | 516.729 |
| 2015-12-01 | 515.639 |
| 2016-01-01 | 512.571 |
| 2016-02-01 | 511.915 |
| 2016-03-01 | 510.531 |
| 2016-04-01 | 509.421 |
| 2016-05-01 | 506.369 |
| 2016-06-01 | 503.804 |
| 2016-07-01 | 501.405 |
| 2016-08-01 | 500.313 |
| 2016-09-01 | 497.879 |
| 2016-10-01 | 495.48 |
| 2016-11-01 | 495.957 |
| 2016-12-01 | 493.178 |
| 2017-01-01 | 494.532 |
| 2017-02-01 | 493.45 |
| 2017-03-01 | 491.522 |
| 2017-04-01 | 491.185 |
| 2017-05-01 | 489.764 |
| 2017-06-01 | 488.856 |
| 2017-07-01 | 487.637 |
| 2017-08-01 | 486.698 |
| 2017-09-01 | 485.446 |
| 2017-10-01 | 485.906 |
| 2017-11-01 | 483.758 |
| 2017-12-01 | 481.546 |
| 2018-01-01 | 479.577 |
| 2018-02-01 | 478.911 |
| 2018-03-01 | 477.935 |
| 2018-04-01 | 476.189 |
| 2018-05-01 | 475.664 |
| 2018-06-01 | 474.503 |
| 2018-07-01 | 474.697 |
| 2018-08-01 | 474.751 |
| 2018-09-01 | 474.496 |
| 2018-10-01 | 473.925 |
| 2018-11-01 | 472.476 |
| 2018-12-01 | 473.5 |
| 2019-01-01 | 473.87 |
| 2019-02-01 | 472.162 |
| 2019-03-01 | 470.01 |
| 2019-04-01 | 469.956 |
| 2019-05-01 | 469.561 |
| 2019-06-01 | 469.26 |
| 2019-07-01 | 468.18 |
| 2019-08-01 | 465.905 |
| 2019-09-01 | 464.93 |
| 2019-10-01 | 463.87 |
| 2019-11-01 | 464.017 |
| 2019-12-01 | 463.44 |
| 2020-01-01 | 462.179 |
| 2020-02-01 | 461.073 |
| 2020-03-01 | 461.128 |
| 2020-04-01 | 462.097 |
| 2020-05-01 | 467.175 |
| 2020-06-01 | 472.442 |
| 2020-07-01 | 476.404 |
| 2020-08-01 | 480.647 |
| 2020-09-01 | 483.901 |
| 2020-10-01 | 487.357 |
| 2020-11-01 | 488.168 |
| 2020-12-01 | 490.194 |
| 2021-01-01 | 491.677 |
| 2021-02-01 | 494.284 |
| 2021-03-01 | 498.035 |
| 2021-04-01 | 496.549 |
| 2021-05-01 | 492.997 |
| 2021-06-01 | 488.478 |
| 2021-07-01 | 485.534 |
| 2021-08-01 | 482.616 |
| 2021-09-01 | 479.673 |
| 2021-10-01 | 474.572 |
| 2021-11-01 | 471.64 |
| 2021-12-01 | 468.567 |
| 2022-01-01 | 464.092 |
| 2022-02-01 | 461.36 |
| 2022-03-01 | 456.805 |
| 2022-04-01 | 453.712 |
| 2022-05-01 | 447.69 |
| 2022-06-01 | 443.203 |
| 2022-07-01 | 438.27 |
| 2022-08-01 | 433.942 |
| 2022-09-01 | 431.336 |
| 2022-10-01 | 429.698 |
| 2022-11-01 | 427.752 |
| 2022-12-01 | 425.018 |
| 2023-01-01 | 422.595 |
| 2023-02-01 | 420.704 |
| 2023-03-01 | 417.983 |
| 2023-04-01 | 415.552 |
| 2023-05-01 | 415.603 |
| 2023-06-01 | 413.581 |
| 2023-07-01 | 413.064 |
| 2023-08-01 | 411.772 |
| 2023-09-01 | 409.723 |
| 2023-10-01 | 409.162 |
| 2023-11-01 | 407.922 |
| 2023-12-01 | 404.845 |
| 2024-01-01 | 401.753 |
| 2024-02-01 | 390.211 |
| 2024-03-01 | 357.858 |

Queensland:

Rolling average vasectomies and births per 100,000 male and female population respectively for 18-24 year olds from April 2016 to December 2024

Vasectomies per 100,000 male population

Births per 100,000 female population

| Month and year | Vasectomies |
| --- | --- |
| 2016-04-01 | 0.679933 |
| 2016-05-01 | 0.618121 |
| 2016-06-01 | 0.70825 |
| 2016-07-01 | 0.708245 |
| 2016-08-01 | 0.743582 |
| 2016-09-01 | 0.849633 |
| 2016-10-01 | 0.885017 |
| 2016-11-01 | 0.884778 |
| 2016-12-01 | 0.955189 |
| 2017-01-01 | 0.955002 |
| 2017-02-01 | 1.06039 |
| 2017-03-01 | 1.02445 |
| 2017-04-01 | 1.16478 |
| 2017-05-01 | 1.16478 |
| 2017-06-01 | 1.05829 |
| 2017-07-01 | 1.1638 |
| 2017-08-01 | 1.09283 |
| 2017-09-01 | 1.022 |
| 2017-10-01 | 1.09273 |
| 2017-11-01 | 1.09271 |
| 2017-12-01 | 0.951165 |
| 2018-01-01 | 0.915975 |
| 2018-02-01 | 0.810322 |
| 2018-03-01 | 0.774968 |
| 2018-04-01 | 0.669345 |
| 2018-05-01 | 0.739461 |
| 2018-06-01 | 0.774429 |
| 2018-07-01 | 0.703415 |
| 2018-08-01 | 0.667908 |
| 2018-09-01 | 0.667617 |
| 2018-10-01 | 0.736624 |
| 2018-11-01 | 0.805979 |
| 2018-12-01 | 0.840657 |
| 2019-01-01 | 0.875419 |
| 2019-02-01 | 0.84031 |
| 2019-03-01 | 0.978665 |
| 2019-04-01 | 0.978099 |
| 2019-05-01 | 0.977534 |
| 2019-06-01 | 0.942193 |
| 2019-07-01 | 1.01128 |
| 2019-08-01 | 1.15002 |
| 2019-09-01 | 1.25403 |
| 2019-10-01 | 1.2192 |
| 2019-11-01 | 1.25447 |
| 2019-12-01 | 1.39488 |
| 2020-01-01 | 1.46501 |
| 2020-02-01 | 1.6748 |
| 2020-03-01 | 1.53616 |
| 2020-04-01 | 1.46661 |
| 2020-05-01 | 1.53767 |
| 2020-06-01 | 1.5732 |
| 2020-07-01 | 1.6821 |
| 2020-08-01 | 1.75626 |
| 2020-09-01 | 1.72354 |
| 2020-10-01 | 1.69019 |
| 2020-11-01 | 1.51481 |
| 2020-12-01 | 1.48261 |
| 2021-01-01 | 1.44931 |
| 2021-02-01 | 1.34691 |
| 2021-03-01 | 1.45598 |
| 2021-04-01 | 1.59964 |
| 2021-05-01 | 1.63861 |
| 2021-06-01 | 1.64014 |
| 2021-07-01 | 1.71648 |
| 2021-08-01 | 1.61272 |
| 2021-09-01 | 1.65059 |
| 2021-10-01 | 1.79555 |
| 2021-11-01 | 1.86752 |
| 2021-12-01 | 1.79723 |
| 2022-01-01 | 1.83218 |
| 2022-02-01 | 1.79581 |
| 2022-03-01 | 1.72336 |
| 2022-04-01 | 1.68571 |
| 2022-05-01 | 1.5768 |
| 2022-06-01 | 1.75232 |
| 2022-07-01 | 1.49799 |
| 2022-08-01 | 1.49361 |
| 2022-09-01 | 1.66383 |
| 2022-10-01 | 1.65373 |
| 2022-11-01 | 1.82357 |
| 2022-12-01 | 1.99196 |
| 2023-01-01 | 2.08909 |
| 2023-02-01 | 2.18778 |
| 2023-03-01 | 2.28491 |
| 2023-04-01 | 2.21252 |
| 2023-05-01 | 2.20908 |
| 2023-06-01 | 2.12981 |
| 2023-07-01 | 2.19249 |
| 2023-08-01 | 2.18533 |
| 2023-09-01 | 2.00356 |
| 2023-10-01 | 2.058 |
| 2023-11-01 | 2.01369 |
| 2023-12-01 | 1.96776 |
| 2024-01-01 | 1.86173 |
| 2024-02-01 | 1.75709 |
| 2024-03-01 | 1.61851 |
| 2024-04-01 | 1.74682 |
| 2024-05-01 | 1.71198 |
| 2024-06-01 | 1.60867 |
| 2024-07-01 | 1.57247 |
| 2024-08-01 | 1.56838 |
| 2024-09-01 | 1.50007 |
| 2024-10-01 | 1.36343 |
| 2024-11-01 | 1.35746 |
| 2024-12-01 | 1.25473 |

|  | Births |
| --- | --- |
| 2015-07-01 | 400.135 |
| 2015-08-01 | 404.153 |
| 2015-09-01 | 405.618 |
| 2015-10-01 | 404.068 |
| 2015-11-01 | 402.879 |
| 2015-12-01 | 403.904 |
| 2016-01-01 | 401.375 |
| 2016-02-01 | 401.682 |
| 2016-03-01 | 399.951 |
| 2016-04-01 | 396.502 |
| 2016-05-01 | 392.98 |
| 2016-06-01 | 389.523 |
| 2016-07-01 | 388.736 |
| 2016-08-01 | 385.155 |
| 2016-09-01 | 381.217 |
| 2016-10-01 | 379.107 |
| 2016-11-01 | 379.16 |
| 2016-12-01 | 376.868 |
| 2017-01-01 | 374.519 |
| 2017-02-01 | 371.012 |
| 2017-03-01 | 366.384 |
| 2017-04-01 | 369.299 |
| 2017-05-01 | 367.868 |
| 2017-06-01 | 367.356 |
| 2017-07-01 | 366.926 |
| 2017-08-01 | 364.004 |
| 2017-09-01 | 363.935 |
| 2017-10-01 | 362.318 |
| 2017-11-01 | 358.964 |
| 2017-12-01 | 357.522 |
| 2018-01-01 | 358.878 |
| 2018-02-01 | 359.737 |
| 2018-03-01 | 360.745 |
| 2018-04-01 | 358.23 |
| 2018-05-01 | 358.603 |
| 2018-06-01 | 354.344 |
| 2018-07-01 | 354.635 |
| 2018-08-01 | 357.612 |
| 2018-09-01 | 355.651 |
| 2018-10-01 | 355.685 |
| 2018-11-01 | 354.682 |
| 2018-12-01 | 351.68 |
| 2019-01-01 | 349.402 |
| 2019-02-01 | 347.443 |
| 2019-03-01 | 344.58 |
| 2019-04-01 | 340.594 |
| 2019-05-01 | 338.643 |
| 2019-06-01 | 340.571 |
| 2019-07-01 | 339.039 |
| 2019-08-01 | 335.036 |
| 2019-09-01 | 333.549 |
| 2019-10-01 | 332.191 |
| 2019-11-01 | 331.011 |
| 2019-12-01 | 331.205 |
| 2020-01-01 | 330.999 |
| 2020-02-01 | 328.454 |
| 2020-03-01 | 330.883 |
| 2020-04-01 | 333.936 |
| 2020-05-01 | 332.405 |
| 2020-06-01 | 333.529 |
| 2020-07-01 | 332.096 |
| 2020-08-01 | 331.955 |
| 2020-09-01 | 333.772 |
| 2020-10-01 | 334.99 |
| 2020-11-01 | 336.096 |
| 2020-12-01 | 336.475 |
| 2021-01-01 | 336.765 |
| 2021-02-01 | 335.824 |
| 2021-03-01 | 335.185 |
| 2021-04-01 | 332.193 |
| 2021-05-01 | 333.531 |
| 2021-06-01 | 330.488 |
| 2021-07-01 | 329.076 |
| 2021-08-01 | 328.888 |
| 2021-09-01 | 327.2 |
| 2021-10-01 | 326.66 |
| 2021-11-01 | 323.966 |
| 2021-12-01 | 323.124 |
| 2022-01-01 | 319.68 |
| 2022-02-01 | 317.957 |
| 2022-03-01 | 314.327 |
| 2022-04-01 | 312.03 |
| 2022-05-01 | 307.205 |
| 2022-06-01 | 303.643 |
| 2022-07-01 | 301.482 |
| 2022-08-01 | 297.97 |
| 2022-09-01 | 294.728 |
| 2022-10-01 | 289.675 |
| 2022-11-01 | 288.885 |
| 2022-12-01 | 285.398 |
| 2023-01-01 | 283.496 |
| 2023-02-01 | 282.426 |
| 2023-03-01 | 280.868 |
| 2023-04-01 | 278.485 |
| 2023-05-01 | 277.515 |
| 2023-06-01 | 274.544 |
| 2023-07-01 | 272.379 |
| 2023-08-01 | 269.176 |
| 2023-09-01 | 266.885 |
| 2023-10-01 | 265.64 |
| 2023-11-01 | 261.633 |
| 2023-12-01 | 258.063 |
| 2024-01-01 | 254.331 |
| 2024-02-01 | 251.447 |
| 2024-03-01 | 236.121 |

Rolling average vasectomies and births per 100,000 male and female population respectively for 25-34 year olds from April 2016 to December 2024

Vasectomies per 100,000 male population

Births per 100,000 female population

| Month and year | Vasectomies |
| --- | --- |
| 2016-04-01 | 39.5612 |
| 2016-05-01 | 39.3131 |
| 2016-06-01 | 39.3986 |
| 2016-07-01 | 39.2142 |
| 2016-08-01 | 39.3401 |
| 2016-09-01 | 39.3755 |
| 2016-10-01 | 38.7 |
| 2016-11-01 | 39.0217 |
| 2016-12-01 | 40.0498 |
| 2017-01-01 | 40.301 |
| 2017-02-01 | 40.4541 |
| 2017-03-01 | 40.7769 |
| 2017-04-01 | 40.2034 |
| 2017-05-01 | 40.0383 |
| 2017-06-01 | 40.5889 |
| 2017-07-01 | 40.951 |
| 2017-08-01 | 41.4222 |
| 2017-09-01 | 41.4477 |
| 2017-10-01 | 42.0576 |
| 2017-11-01 | 41.9892 |
| 2017-12-01 | 41.6769 |
| 2018-01-01 | 41.6915 |
| 2018-02-01 | 42.4448 |
| 2018-03-01 | 43.1256 |
| 2018-04-01 | 43.9059 |
| 2018-05-01 | 44.7058 |
| 2018-06-01 | 44.2128 |
| 2018-07-01 | 44.2172 |
| 2018-08-01 | 44.3296 |
| 2018-09-01 | 44.4789 |
| 2018-10-01 | 45.0002 |
| 2018-11-01 | 45.6161 |
| 2018-12-01 | 45.4264 |
| 2019-01-01 | 45.3169 |
| 2019-02-01 | 44.5386 |
| 2019-03-01 | 44.2778 |
| 2019-04-01 | 44.6828 |
| 2019-05-01 | 45.0125 |
| 2019-06-01 | 44.9001 |
| 2019-07-01 | 45.7865 |
| 2019-08-01 | 46.426 |
| 2019-09-01 | 47.1265 |
| 2019-10-01 | 47.8134 |
| 2019-11-01 | 48.5688 |
| 2019-12-01 | 48.4711 |
| 2020-01-01 | 49.0852 |
| 2020-02-01 | 49.8376 |
| 2020-03-01 | 49.3749 |
| 2020-04-01 | 47.6225 |
| 2020-05-01 | 47.5181 |
| 2020-06-01 | 48.5592 |
| 2020-07-01 | 50.7373 |
| 2020-08-01 | 51.2582 |
| 2020-09-01 | 52.1317 |
| 2020-10-01 | 53.3742 |
| 2020-11-01 | 52.4934 |
| 2020-12-01 | 52.8242 |
| 2021-01-01 | 53.377 |
| 2021-02-01 | 53.6274 |
| 2021-03-01 | 54.505 |
| 2021-04-01 | 56.7283 |
| 2021-05-01 | 57.1852 |
| 2021-06-01 | 56.8737 |
| 2021-07-01 | 54.4085 |
| 2021-08-01 | 52.759 |
| 2021-09-01 | 52.4857 |
| 2021-10-01 | 52.0355 |
| 2021-11-01 | 52.3568 |
| 2021-12-01 | 53.8205 |
| 2022-01-01 | 53.4159 |
| 2022-02-01 | 52.7341 |
| 2022-03-01 | 52.6776 |
| 2022-04-01 | 52.1537 |
| 2022-05-01 | 52.2239 |
| 2022-06-01 | 52.6899 |
| 2022-07-01 | 52.6688 |
| 2022-08-01 | 54.1958 |
| 2022-09-01 | 55.1597 |
| 2022-10-01 | 55.2486 |
| 2022-11-01 | 56.4812 |
| 2022-12-01 | 55.9084 |
| 2023-01-01 | 56.6125 |
| 2023-02-01 | 57.398 |
| 2023-03-01 | 59.1106 |
| 2023-04-01 | 59.2394 |
| 2023-05-01 | 59.7008 |
| 2023-06-01 | 59.3949 |
| 2023-07-01 | 60.0196 |
| 2023-08-01 | 59.6846 |
| 2023-09-01 | 58.6527 |
| 2023-10-01 | 58.4022 |
| 2023-11-01 | 58.3894 |
| 2023-12-01 | 57.0301 |
| 2024-01-01 | 56.2126 |
| 2024-02-01 | 56.0218 |
| 2024-03-01 | 54.2403 |
| 2024-04-01 | 53.5629 |
| 2024-05-01 | 53.2058 |
| 2024-06-01 | 52.6831 |
| 2024-07-01 | 52.2339 |
| 2024-08-01 | 51.4974 |
| 2024-09-01 | 50.9868 |
| 2024-10-01 | 51.0027 |
| 2024-11-01 | 51.3485 |
| 2024-12-01 | 52 |

|  | Births |
| --- | --- |
| 2015-07-01 | 910.97 |
| 2015-08-01 | 911.744 |
| 2015-09-01 | 913.972 |
| 2015-10-01 | 907.11 |
| 2015-11-01 | 907.719 |
| 2015-12-01 | 910.28 |
| 2016-01-01 | 912.083 |
| 2016-02-01 | 910.613 |
| 2016-03-01 | 908.313 |
| 2016-04-01 | 906.08 |
| 2016-05-01 | 897.452 |
| 2016-06-01 | 892.003 |
| 2016-07-01 | 886.859 |
| 2016-08-01 | 883.577 |
| 2016-09-01 | 880.194 |
| 2016-10-01 | 877.096 |
| 2016-11-01 | 875.407 |
| 2016-12-01 | 866.687 |
| 2017-01-01 | 859.902 |
| 2017-02-01 | 858.9 |
| 2017-03-01 | 856.399 |
| 2017-04-01 | 857.734 |
| 2017-05-01 | 860.951 |
| 2017-06-01 | 859.192 |
| 2017-07-01 | 855.182 |
| 2017-08-01 | 855.139 |
| 2017-09-01 | 851.964 |
| 2017-10-01 | 849.627 |
| 2017-11-01 | 845.335 |
| 2017-12-01 | 845.363 |
| 2018-01-01 | 845.156 |
| 2018-02-01 | 844.688 |
| 2018-03-01 | 842.44 |
| 2018-04-01 | 838.151 |
| 2018-05-01 | 833.568 |
| 2018-06-01 | 833.452 |
| 2018-07-01 | 837.071 |
| 2018-08-01 | 839.122 |
| 2018-09-01 | 835.64 |
| 2018-10-01 | 836.992 |
| 2018-11-01 | 835.799 |
| 2018-12-01 | 833.72 |
| 2019-01-01 | 833.978 |
| 2019-02-01 | 831.974 |
| 2019-03-01 | 831.337 |
| 2019-04-01 | 831.49 |
| 2019-05-01 | 830.442 |
| 2019-06-01 | 828.689 |
| 2019-07-01 | 823.238 |
| 2019-08-01 | 819.408 |
| 2019-09-01 | 822.622 |
| 2019-10-01 | 822.903 |
| 2019-11-01 | 822.59 |
| 2019-12-01 | 822.87 |
| 2020-01-01 | 819.459 |
| 2020-02-01 | 817.608 |
| 2020-03-01 | 818.777 |
| 2020-04-01 | 821.615 |
| 2020-05-01 | 831.722 |
| 2020-06-01 | 840.502 |
| 2020-07-01 | 850.419 |
| 2020-08-01 | 855.226 |
| 2020-09-01 | 859.758 |
| 2020-10-01 | 864.779 |
| 2020-11-01 | 869.006 |
| 2020-12-01 | 873.574 |
| 2021-01-01 | 876.045 |
| 2021-02-01 | 881.719 |
| 2021-03-01 | 885.587 |
| 2021-04-01 | 882.246 |
| 2021-05-01 | 874.312 |
| 2021-06-01 | 871.468 |
| 2021-07-01 | 866.548 |
| 2021-08-01 | 863.425 |
| 2021-09-01 | 859.583 |
| 2021-10-01 | 851.56 |
| 2021-11-01 | 847.945 |
| 2021-12-01 | 842.632 |
| 2022-01-01 | 839.743 |
| 2022-02-01 | 835.463 |
| 2022-03-01 | 827.869 |
| 2022-04-01 | 823.061 |
| 2022-05-01 | 813.661 |
| 2022-06-01 | 800.837 |
| 2022-07-01 | 791.067 |
| 2022-08-01 | 785.052 |
| 2022-09-01 | 779.261 |
| 2022-10-01 | 776.068 |
| 2022-11-01 | 770.532 |
| 2022-12-01 | 765.023 |
| 2023-01-01 | 761.277 |
| 2023-02-01 | 755.923 |
| 2023-03-01 | 750.755 |
| 2023-04-01 | 746.769 |
| 2023-05-01 | 748.48 |
| 2023-06-01 | 745.624 |
| 2023-07-01 | 744.131 |
| 2023-08-01 | 740.561 |
| 2023-09-01 | 735.087 |
| 2023-10-01 | 731.423 |
| 2023-11-01 | 726.701 |
| 2023-12-01 | 721.067 |
| 2024-01-01 | 717.054 |
| 2024-02-01 | 706.423 |
| 2024-03-01 | 669.871 |

Rolling average vasectomies and births per 100,000 male and female population respectively for 35-44 year olds from April 2016 to December 2024

Vasectomies per 100,000 male population

Births per 100,000 female population

| Month and year | Vasectomies |
| --- | --- |
| 2016-04-01 | 96.8103 |
| 2016-05-01 | 97.2815 |
| 2016-06-01 | 97.8578 |
| 2016-07-01 | 98.1332 |
| 2016-08-01 | 99.6134 |
| 2016-09-01 | 99.2582 |
| 2016-10-01 | 99.6677 |
| 2016-11-01 | 100.888 |
| 2016-12-01 | 100.563 |
| 2017-01-01 | 102.371 |
| 2017-02-01 | 102.838 |
| 2017-03-01 | 104.299 |
| 2017-04-01 | 103.132 |
| 2017-05-01 | 102.618 |
| 2017-06-01 | 102.573 |
| 2017-07-01 | 102.201 |
| 2017-08-01 | 101.486 |
| 2017-09-01 | 103.209 |
| 2017-10-01 | 104.083 |
| 2017-11-01 | 103.388 |
| 2017-12-01 | 104.311 |
| 2018-01-01 | 104.368 |
| 2018-02-01 | 104.984 |
| 2018-03-01 | 105.336 |
| 2018-04-01 | 107.84 |
| 2018-05-01 | 109.873 |
| 2018-06-01 | 112.106 |
| 2018-07-01 | 113.599 |
| 2018-08-01 | 115.807 |
| 2018-09-01 | 115.289 |
| 2018-10-01 | 116.437 |
| 2018-11-01 | 117.869 |
| 2018-12-01 | 116.268 |
| 2019-01-01 | 117.21 |
| 2019-02-01 | 118.234 |
| 2019-03-01 | 119.739 |
| 2019-04-01 | 118.896 |
| 2019-05-01 | 119.554 |
| 2019-06-01 | 117.423 |
| 2019-07-01 | 117.631 |
| 2019-08-01 | 118.99 |
| 2019-09-01 | 121.556 |
| 2019-10-01 | 122.73 |
| 2019-11-01 | 123.801 |
| 2019-12-01 | 125.266 |
| 2020-01-01 | 126.483 |
| 2020-02-01 | 127.066 |
| 2020-03-01 | 124.975 |
| 2020-04-01 | 120.207 |
| 2020-05-01 | 118.285 |
| 2020-06-01 | 121.95 |
| 2020-07-01 | 125.947 |
| 2020-08-01 | 126.94 |
| 2020-09-01 | 128.284 |
| 2020-10-01 | 129.426 |
| 2020-11-01 | 131.187 |
| 2020-12-01 | 132.786 |
| 2021-01-01 | 133.707 |
| 2021-02-01 | 135.395 |
| 2021-03-01 | 139.213 |
| 2021-04-01 | 145.777 |
| 2021-05-01 | 148.059 |
| 2021-06-01 | 146.029 |
| 2021-07-01 | 143.682 |
| 2021-08-01 | 140.28 |
| 2021-09-01 | 139.901 |
| 2021-10-01 | 139.354 |
| 2021-11-01 | 138.14 |
| 2021-12-01 | 138.331 |
| 2022-01-01 | 134.717 |
| 2022-02-01 | 131.417 |
| 2022-03-01 | 127.751 |
| 2022-04-01 | 126.099 |
| 2022-05-01 | 126.423 |
| 2022-06-01 | 126.142 |
| 2022-07-01 | 124.577 |
| 2022-08-01 | 126.873 |
| 2022-09-01 | 128.481 |
| 2022-10-01 | 129.793 |
| 2022-11-01 | 130.473 |
| 2022-12-01 | 130.427 |
| 2023-01-01 | 133.413 |
| 2023-02-01 | 136.167 |
| 2023-03-01 | 141.087 |
| 2023-04-01 | 141.529 |
| 2023-05-01 | 141.113 |
| 2023-06-01 | 142.122 |
| 2023-07-01 | 143.115 |
| 2023-08-01 | 141.704 |
| 2023-09-01 | 139.296 |
| 2023-10-01 | 138.565 |
| 2023-11-01 | 139.986 |
| 2023-12-01 | 139.447 |
| 2024-01-01 | 137.648 |
| 2024-02-01 | 137.445 |
| 2024-03-01 | 134.252 |
| 2024-04-01 | 133.963 |
| 2024-05-01 | 134.812 |
| 2024-06-01 | 133.204 |
| 2024-07-01 | 132.058 |
| 2024-08-01 | 131.809 |
| 2024-09-01 | 129.542 |
| 2024-10-01 | 128.442 |
| 2024-11-01 | 126.53 |
| 2024-12-01 | 126.148 |

|  | Births |
| --- | --- |
| 2015-07-01 | 251.241 |
| 2015-08-01 | 251.509 |
| 2015-09-01 | 251.4 |
| 2015-10-01 | 251.21 |
| 2015-11-01 | 250.485 |
| 2015-12-01 | 249.682 |
| 2016-01-01 | 248.075 |
| 2016-02-01 | 248.965 |
| 2016-03-01 | 246.572 |
| 2016-04-01 | 245.132 |
| 2016-05-01 | 242.12 |
| 2016-06-01 | 239.091 |
| 2016-07-01 | 237.001 |
| 2016-08-01 | 237.679 |
| 2016-09-01 | 238.157 |
| 2016-10-01 | 238.041 |
| 2016-11-01 | 239.761 |
| 2016-12-01 | 240.034 |
| 2017-01-01 | 240.124 |
| 2017-02-01 | 239.194 |
| 2017-03-01 | 241.589 |
| 2017-04-01 | 242.091 |
| 2017-05-01 | 243.775 |
| 2017-06-01 | 245.657 |
| 2017-07-01 | 246.41 |
| 2017-08-01 | 246.081 |
| 2017-09-01 | 246.134 |
| 2017-10-01 | 246.328 |
| 2017-11-01 | 247.32 |
| 2017-12-01 | 247.452 |
| 2018-01-01 | 249.858 |
| 2018-02-01 | 250.68 |
| 2018-03-01 | 250.846 |
| 2018-04-01 | 251.555 |
| 2018-05-01 | 253.117 |
| 2018-06-01 | 252.245 |
| 2018-07-01 | 252.456 |
| 2018-08-01 | 253.751 |
| 2018-09-01 | 252.5 |
| 2018-10-01 | 252.667 |
| 2018-11-01 | 251.385 |
| 2018-12-01 | 251.421 |
| 2019-01-01 | 251.818 |
| 2019-02-01 | 251.156 |
| 2019-03-01 | 250.008 |
| 2019-04-01 | 249.492 |
| 2019-05-01 | 248.472 |
| 2019-06-01 | 247.322 |
| 2019-07-01 | 248.253 |
| 2019-08-01 | 247.15 |
| 2019-09-01 | 248.38 |
| 2019-10-01 | 248.283 |
| 2019-11-01 | 248.06 |
| 2019-12-01 | 248.935 |
| 2020-01-01 | 246.676 |
| 2020-02-01 | 247.833 |
| 2020-03-01 | 247.479 |
| 2020-04-01 | 246.727 |
| 2020-05-01 | 248.132 |
| 2020-06-01 | 250.645 |
| 2020-07-01 | 252.222 |
| 2020-08-01 | 252.226 |
| 2020-09-01 | 252.916 |
| 2020-10-01 | 254.625 |
| 2020-11-01 | 256.751 |
| 2020-12-01 | 256.263 |
| 2021-01-01 | 259.863 |
| 2021-02-01 | 259.797 |
| 2021-03-01 | 260.772 |
| 2021-04-01 | 261.712 |
| 2021-05-01 | 260.771 |
| 2021-06-01 | 260.048 |
| 2021-07-01 | 258.56 |
| 2021-08-01 | 259.854 |
| 2021-09-01 | 259.584 |
| 2021-10-01 | 257.642 |
| 2021-11-01 | 256.533 |
| 2021-12-01 | 255.967 |
| 2022-01-01 | 252.517 |
| 2022-02-01 | 252.414 |
| 2022-03-01 | 252.743 |
| 2022-04-01 | 252.42 |
| 2022-05-01 | 250.249 |
| 2022-06-01 | 247.997 |
| 2022-07-01 | 245.374 |
| 2022-08-01 | 242.333 |
| 2022-09-01 | 241.414 |
| 2022-10-01 | 241.093 |
| 2022-11-01 | 240.41 |
| 2022-12-01 | 238.397 |
| 2023-01-01 | 240.224 |
| 2023-02-01 | 240.045 |
| 2023-03-01 | 238.745 |
| 2023-04-01 | 238.729 |
| 2023-05-01 | 239.319 |
| 2023-06-01 | 238.658 |
| 2023-07-01 | 239.008 |
| 2023-08-01 | 240.215 |
| 2023-09-01 | 237.477 |
| 2023-10-01 | 235.717 |
| 2023-11-01 | 233.744 |
| 2023-12-01 | 232.971 |
| 2024-01-01 | 229.889 |
| 2024-02-01 | 225.673 |
| 2024-03-01 | 213.099 |

South Australia:

Rolling average vasectomies and births per 100,000 male and female population respectively for 18-24 year olds from April 2016 to December 2024

Vasectomies per 100,000 male population

Births per 100,000 female population

| Month and year | Vasectomies |
| --- | --- |
| 2016-04-01 | 0 |
| 2016-05-01 | 0 |
| 2016-06-01 | 0 |
| 2016-07-01 | 0.102477 |
| 2016-08-01 | 0.102477 |
| 2016-09-01 | 0.102477 |
| 2016-10-01 | 0.102477 |
| 2016-11-01 | 0.102477 |
| 2016-12-01 | 0.102477 |
| 2017-01-01 | 0.102477 |
| 2017-02-01 | 0.20469 |
| 2017-03-01 | 0.20469 |
| 2017-04-01 | 0.20469 |
| 2017-05-01 | 0.306972 |
| 2017-06-01 | 0.409254 |
| 2017-07-01 | 0.306777 |
| 2017-08-01 | 0.306777 |
| 2017-09-01 | 0.306777 |
| 2017-10-01 | 0.306777 |
| 2017-11-01 | 0.306777 |
| 2017-12-01 | 0.306777 |
| 2018-01-01 | 0.306777 |
| 2018-02-01 | 0.204564 |
| 2018-03-01 | 0.204564 |
| 2018-04-01 | 0.204564 |
| 2018-05-01 | 0.102282 |
| 2018-06-01 | 0.101653 |
| 2018-07-01 | 0.101653 |
| 2018-08-01 | 0.101653 |
| 2018-09-01 | 0.202906 |
| 2018-10-01 | 0.406317 |
| 2018-11-01 | 0.406317 |
| 2018-12-01 | 0.406317 |
| 2019-01-01 | 0.406317 |
| 2019-02-01 | 0.406317 |
| 2019-03-01 | 0.406317 |
| 2019-04-01 | 0.507306 |
| 2019-05-01 | 0.709285 |
| 2019-06-01 | 0.708621 |
| 2019-07-01 | 0.708621 |
| 2019-08-01 | 0.708621 |
| 2019-09-01 | 0.708373 |
| 2019-10-01 | 0.504962 |
| 2019-11-01 | 0.504962 |
| 2019-12-01 | 0.504962 |
| 2020-01-01 | 0.504962 |
| 2020-02-01 | 0.504962 |
| 2020-03-01 | 0.504962 |
| 2020-04-01 | 0.506265 |
| 2020-05-01 | 0.304287 |
| 2020-06-01 | 0.30559 |
| 2020-07-01 | 0.408839 |
| 2020-08-01 | 0.408839 |
| 2020-09-01 | 0.307833 |
| 2020-10-01 | 0.307833 |
| 2020-11-01 | 0.307833 |
| 2020-12-01 | 0.516535 |
| 2021-01-01 | 0.516535 |
| 2021-02-01 | 0.621622 |
| 2021-03-01 | 0.72671 |
| 2021-04-01 | 0.624417 |
| 2021-05-01 | 0.624417 |
| 2021-06-01 | 0.522125 |
| 2021-07-01 | 0.525413 |
| 2021-08-01 | 0.63195 |
| 2021-09-01 | 0.63195 |
| 2021-10-01 | 0.63195 |
| 2021-11-01 | 0.845134 |
| 2021-12-01 | 0.743024 |
| 2022-01-01 | 0.743024 |
| 2022-02-01 | 0.637937 |
| 2022-03-01 | 0.637807 |
| 2022-04-01 | 0.637807 |
| 2022-05-01 | 0.637807 |
| 2022-06-01 | 0.637807 |
| 2022-07-01 | 0.634556 |
| 2022-08-01 | 0.631305 |
| 2022-09-01 | 0.734591 |
| 2022-10-01 | 1.04339 |
| 2022-11-01 | 0.933144 |
| 2022-12-01 | 1.03242 |
| 2023-01-01 | 1.03242 |
| 2023-02-01 | 1.03242 |
| 2023-03-01 | 0.927463 |
| 2023-04-01 | 0.927463 |
| 2023-05-01 | 0.927463 |
| 2023-06-01 | 0.927463 |
| 2023-07-01 | 1.12374 |
| 2023-08-01 | 1.12031 |
| 2023-09-01 | 1.11688 |
| 2023-10-01 | 0.808079 |
| 2023-11-01 | 0.705145 |
| 2023-12-01 | 0.599244 |
| 2024-01-01 | 0.698265 |
| 2024-02-01 | 0.698265 |
| 2024-03-01 | 0.698265 |
| 2024-04-01 | 0.896801 |
| 2024-05-01 | 1.09534 |
| 2024-06-01 | 1.09534 |
| 2024-07-01 | 0.894445 |
| 2024-08-01 | 0.893265 |
| 2024-09-01 | 0.79341 |
| 2024-10-01 | 0.79341 |
| 2024-11-01 | 0.892293 |
| 2024-12-01 | 0.891208 |

|  | Births |
| --- | --- |
| 2015-07-01 | 308.135 |
| 2015-08-01 | 306.6 |
| 2015-09-01 | 305.644 |
| 2015-10-01 | 302.353 |
| 2015-11-01 | 303.266 |
| 2015-12-01 | 301.588 |
| 2016-01-01 | 300.689 |
| 2016-02-01 | 296.324 |
| 2016-03-01 | 293.259 |
| 2016-04-01 | 291.753 |
| 2016-05-01 | 292.076 |
| 2016-06-01 | 290.893 |
| 2016-07-01 | 286.243 |
| 2016-08-01 | 285.153 |
| 2016-09-01 | 286.865 |
| 2016-10-01 | 285.634 |
| 2016-11-01 | 279.983 |
| 2016-12-01 | 278.861 |
| 2017-01-01 | 275.46 |
| 2017-02-01 | 277.145 |
| 2017-03-01 | 275.693 |
| 2017-04-01 | 276.741 |
| 2017-05-01 | 274.455 |
| 2017-06-01 | 272.919 |
| 2017-07-01 | 275.379 |
| 2017-08-01 | 272.56 |
| 2017-09-01 | 268.338 |
| 2017-10-01 | 267.54 |
| 2017-11-01 | 269.214 |
| 2017-12-01 | 265.839 |
| 2018-01-01 | 269.488 |
| 2018-02-01 | 268.712 |
| 2018-03-01 | 270.311 |
| 2018-04-01 | 266.942 |
| 2018-05-01 | 265.402 |
| 2018-06-01 | 261.71 |
| 2018-07-01 | 256.963 |
| 2018-08-01 | 256.957 |
| 2018-09-01 | 254.149 |
| 2018-10-01 | 252.617 |
| 2018-11-01 | 249.148 |
| 2018-12-01 | 250.731 |
| 2019-01-01 | 244.125 |
| 2019-02-01 | 240.534 |
| 2019-03-01 | 238.809 |
| 2019-04-01 | 237.996 |
| 2019-05-01 | 237.929 |
| 2019-06-01 | 238.098 |
| 2019-07-01 | 238.779 |
| 2019-08-01 | 237.052 |
| 2019-09-01 | 236.293 |
| 2019-10-01 | 234.485 |
| 2019-11-01 | 233.097 |
| 2019-12-01 | 230.842 |
| 2020-01-01 | 231.606 |
| 2020-02-01 | 230.572 |
| 2020-03-01 | 228.045 |
| 2020-04-01 | 231.812 |
| 2020-05-01 | 232.758 |
| 2020-06-01 | 233.083 |
| 2020-07-01 | 233.204 |
| 2020-08-01 | 230.875 |
| 2020-09-01 | 229.788 |
| 2020-10-01 | 229.488 |
| 2020-11-01 | 229.972 |
| 2020-12-01 | 229.309 |
| 2021-01-01 | 227.75 |
| 2021-02-01 | 228.189 |
| 2021-03-01 | 228.895 |
| 2021-04-01 | 225.156 |
| 2021-05-01 | 220.389 |
| 2021-06-01 | 218.559 |
| 2021-07-01 | 216.108 |
| 2021-08-01 | 217.735 |
| 2021-09-01 | 219.358 |
| 2021-10-01 | 220.097 |
| 2021-11-01 | 219.823 |
| 2021-12-01 | 218.348 |
| 2022-01-01 | 217.705 |
| 2022-02-01 | 215.283 |
| 2022-03-01 | 210.922 |
| 2022-04-01 | 208.925 |
| 2022-05-01 | 206.698 |
| 2022-06-01 | 205.26 |
| 2022-07-01 | 199.632 |
| 2022-08-01 | 197.425 |
| 2022-09-01 | 193.982 |
| 2022-10-01 | 190.015 |
| 2022-11-01 | 185.737 |
| 2022-12-01 | 183.049 |
| 2023-01-01 | 180.113 |
| 2023-02-01 | 178.974 |
| 2023-03-01 | 177.186 |
| 2023-04-01 | 174.455 |
| 2023-05-01 | 174.837 |
| 2023-06-01 | 169.73 |
| 2023-07-01 | 172.76 |
| 2023-08-01 | 170.013 |
| 2023-09-01 | 167.052 |
| 2023-10-01 | 165.89 |
| 2023-11-01 | 164.956 |
| 2023-12-01 | 165.217 |
| 2024-01-01 | 161.935 |
| 2024-02-01 | 157.255 |
| 2024-03-01 | 148.707 |

Rolling average vasectomies and births per 100,000 male and female population respectively for 25-34 year olds from April 2016 to December 2024

Vasectomies per 100,000 male population

Births per 100,000 female population

| Month and year | Vasectomies |
| --- | --- |
| 2016-04-01 | 17.9462 |
| 2016-05-01 | 17.8886 |
| 2016-06-01 | 18.2013 |
| 2016-07-01 | 17.9774 |
| 2016-08-01 | 17.6833 |
| 2016-09-01 | 17.1717 |
| 2016-10-01 | 16.73 |
| 2016-11-01 | 17.2246 |
| 2016-12-01 | 17.2866 |
| 2017-01-01 | 18.0013 |
| 2017-02-01 | 17.8504 |
| 2017-03-01 | 18.8486 |
| 2017-04-01 | 19.0543 |
| 2017-05-01 | 19.5461 |
| 2017-06-01 | 18.6741 |
| 2017-07-01 | 18.7333 |
| 2017-08-01 | 19.7967 |
| 2017-09-01 | 20.4303 |
| 2017-10-01 | 21.0639 |
| 2017-11-01 | 20.9076 |
| 2017-12-01 | 20.6813 |
| 2018-01-01 | 20.243 |
| 2018-02-01 | 20.2308 |
| 2018-03-01 | 19.7124 |
| 2018-04-01 | 19.7684 |
| 2018-05-01 | 19.6791 |
| 2018-06-01 | 19.8832 |
| 2018-07-01 | 20.0063 |
| 2018-08-01 | 20.4069 |
| 2018-09-01 | 19.8915 |
| 2018-10-01 | 20.5035 |
| 2018-11-01 | 20.8349 |
| 2018-12-01 | 20.4637 |
| 2019-01-01 | 20.8749 |
| 2019-02-01 | 20.8585 |
| 2019-03-01 | 20.6262 |
| 2019-04-01 | 20.3961 |
| 2019-05-01 | 20.0932 |
| 2019-06-01 | 20.7105 |
| 2019-07-01 | 20.2671 |
| 2019-08-01 | 19.5342 |
| 2019-09-01 | 20.0031 |
| 2019-10-01 | 19.6169 |
| 2019-11-01 | 19.2368 |
| 2019-12-01 | 19.6364 |
| 2020-01-01 | 19.616 |
| 2020-02-01 | 19.9401 |
| 2020-03-01 | 19.9843 |
| 2020-04-01 | 18.8583 |
| 2020-05-01 | 17.6612 |
| 2020-06-01 | 17.1515 |
| 2020-07-01 | 17.6209 |
| 2020-08-01 | 17.533 |
| 2020-09-01 | 17.7212 |
| 2020-10-01 | 18.1156 |
| 2020-11-01 | 18.1688 |
| 2020-12-01 | 18.4307 |
| 2021-01-01 | 18.4927 |
| 2021-02-01 | 18.0712 |
| 2021-03-01 | 19.85 |
| 2021-04-01 | 21.6947 |
| 2021-05-01 | 23.0598 |
| 2021-06-01 | 23.5956 |
| 2021-07-01 | 22.4838 |
| 2021-08-01 | 21.5779 |
| 2021-09-01 | 21.3535 |
| 2021-10-01 | 19.9601 |
| 2021-11-01 | 20.3472 |
| 2021-12-01 | 21.1431 |
| 2022-01-01 | 20.9824 |
| 2022-02-01 | 21.8338 |
| 2022-03-01 | 21.2279 |
| 2022-04-01 | 21.9127 |
| 2022-05-01 | 22.7408 |
| 2022-06-01 | 23.634 |
| 2022-07-01 | 24.8657 |
| 2022-08-01 | 26.9526 |
| 2022-09-01 | 30.3305 |
| 2022-10-01 | 31.4659 |
| 2022-11-01 | 31.1857 |
| 2022-12-01 | 30.431 |
| 2023-01-01 | 31.0358 |
| 2023-02-01 | 31.2159 |
| 2023-03-01 | 30.6573 |
| 2023-04-01 | 30.6798 |
| 2023-05-01 | 31.1623 |
| 2023-06-01 | 30.8042 |
| 2023-07-01 | 31.563 |
| 2023-08-01 | 31.9038 |
| 2023-09-01 | 29.7409 |
| 2023-10-01 | 29.7962 |
| 2023-11-01 | 30.3088 |
| 2023-12-01 | 31.2686 |
| 2024-01-01 | 31.856 |
| 2024-02-01 | 31.7352 |
| 2024-03-01 | 32.5006 |
| 2024-04-01 | 31.5018 |
| 2024-05-01 | 31.7611 |
| 2024-06-01 | 31.0872 |
| 2024-07-01 | 31.2979 |
| 2024-08-01 | 30.9344 |
| 2024-09-01 | 30.8262 |
| 2024-10-01 | 30.79 |
| 2024-11-01 | 31.6301 |
| 2024-12-01 | 30.898 |

|  | Births |
| --- | --- |
| 2015-07-01 | 919.136 |
| 2015-08-01 | 919.127 |
| 2015-09-01 | 918.758 |
| 2015-10-01 | 915.491 |
| 2015-11-01 | 918.862 |
| 2015-12-01 | 919.577 |
| 2016-01-01 | 913.629 |
| 2016-02-01 | 916.986 |
| 2016-03-01 | 909.238 |
| 2016-04-01 | 909.79 |
| 2016-05-01 | 906.169 |
| 2016-06-01 | 904.264 |
| 2016-07-01 | 900.081 |
| 2016-08-01 | 894.744 |
| 2016-09-01 | 889.839 |
| 2016-10-01 | 885.164 |
| 2016-11-01 | 877.936 |
| 2016-12-01 | 874.743 |
| 2017-01-01 | 875.945 |
| 2017-02-01 | 871.664 |
| 2017-03-01 | 869.886 |
| 2017-04-01 | 869.412 |
| 2017-05-01 | 869.393 |
| 2017-06-01 | 864.168 |
| 2017-07-01 | 862.062 |
| 2017-08-01 | 860.03 |
| 2017-09-01 | 859.556 |
| 2017-10-01 | 857.092 |
| 2017-11-01 | 855.018 |
| 2017-12-01 | 850.466 |
| 2018-01-01 | 845.371 |
| 2018-02-01 | 845.497 |
| 2018-03-01 | 844.756 |
| 2018-04-01 | 838.39 |
| 2018-05-01 | 835.492 |
| 2018-06-01 | 838.248 |
| 2018-07-01 | 840.608 |
| 2018-08-01 | 841.565 |
| 2018-09-01 | 839.347 |
| 2018-10-01 | 841.208 |
| 2018-11-01 | 842.46 |
| 2018-12-01 | 844.509 |
| 2019-01-01 | 852.777 |
| 2019-02-01 | 847.901 |
| 2019-03-01 | 844.404 |
| 2019-04-01 | 843.694 |
| 2019-05-01 | 840.652 |
| 2019-06-01 | 838.5 |
| 2019-07-01 | 834.494 |
| 2019-08-01 | 833.349 |
| 2019-09-01 | 830.005 |
| 2019-10-01 | 826.375 |
| 2019-11-01 | 820.645 |
| 2019-12-01 | 816.033 |
| 2020-01-01 | 804.59 |
| 2020-02-01 | 806.313 |
| 2020-03-01 | 809.281 |
| 2020-04-01 | 817.665 |
| 2020-05-01 | 829.5 |
| 2020-06-01 | 837.265 |
| 2020-07-01 | 842.841 |
| 2020-08-01 | 851.685 |
| 2020-09-01 | 857.742 |
| 2020-10-01 | 862.028 |
| 2020-11-01 | 865.205 |
| 2020-12-01 | 870.882 |
| 2021-01-01 | 877.836 |
| 2021-02-01 | 880.429 |
| 2021-03-01 | 882.458 |
| 2021-04-01 | 873.831 |
| 2021-05-01 | 870.549 |
| 2021-06-01 | 865.058 |
| 2021-07-01 | 854.637 |
| 2021-08-01 | 847.191 |
| 2021-09-01 | 844.845 |
| 2021-10-01 | 839.525 |
| 2021-11-01 | 834.342 |
| 2021-12-01 | 833.822 |
| 2022-01-01 | 827.021 |
| 2022-02-01 | 824.161 |
| 2022-03-01 | 819.925 |
| 2022-04-01 | 813.129 |
| 2022-05-01 | 802.682 |
| 2022-06-01 | 791.675 |
| 2022-07-01 | 786.981 |
| 2022-08-01 | 780.189 |
| 2022-09-01 | 772.327 |
| 2022-10-01 | 770.88 |
| 2022-11-01 | 767.34 |
| 2022-12-01 | 760.049 |
| 2023-01-01 | 762.5 |
| 2023-02-01 | 757.402 |
| 2023-03-01 | 746.726 |
| 2023-04-01 | 743.914 |
| 2023-05-01 | 745.772 |
| 2023-06-01 | 743.459 |
| 2023-07-01 | 744.074 |
| 2023-08-01 | 742.45 |
| 2023-09-01 | 740.361 |
| 2023-10-01 | 738.189 |
| 2023-11-01 | 737.528 |
| 2023-12-01 | 730.453 |
| 2024-01-01 | 721.307 |
| 2024-02-01 | 714.782 |
| 2024-03-01 | 684.433 |

Rolling average vasectomies and births per 100,000 male and female population respectively for 35-44 year olds from April 2016 to December 2024

Vasectomies per 100,000 male population

Births per 100,000 female population

| Month and year | Vasectomies |
| --- | --- |
| 2016-04-01 | 66.8229 |
| 2016-05-01 | 66.7143 |
| 2016-06-01 | 68.4989 |
| 2016-07-01 | 67.6213 |
| 2016-08-01 | 67.1299 |
| 2016-09-01 | 66.9498 |
| 2016-10-01 | 68.4683 |
| 2016-11-01 | 68.5119 |
| 2016-12-01 | 69.4874 |
| 2017-01-01 | 69.7417 |
| 2017-02-01 | 69.4576 |
| 2017-03-01 | 70.5784 |
| 2017-04-01 | 70.124 |
| 2017-05-01 | 70.9244 |
| 2017-06-01 | 70.2416 |
| 2017-07-01 | 68.444 |
| 2017-08-01 | 69.6962 |
| 2017-09-01 | 69.8537 |
| 2017-10-01 | 70.6149 |
| 2017-11-01 | 72.1552 |
| 2017-12-01 | 71.276 |
| 2018-01-01 | 71.9446 |
| 2018-02-01 | 73.0683 |
| 2018-03-01 | 74.2676 |
| 2018-04-01 | 75.4615 |
| 2018-05-01 | 76.019 |
| 2018-06-01 | 76.9604 |
| 2018-07-01 | 78.6831 |
| 2018-08-01 | 78.5936 |
| 2018-09-01 | 77.5117 |
| 2018-10-01 | 77.2656 |
| 2018-11-01 | 77.4637 |
| 2018-12-01 | 78.6229 |
| 2019-01-01 | 78.3988 |
| 2019-02-01 | 78.1459 |
| 2019-03-01 | 77.3509 |
| 2019-04-01 | 77.5614 |
| 2019-05-01 | 78.5967 |
| 2019-06-01 | 78.2455 |
| 2019-07-01 | 78.5289 |
| 2019-08-01 | 78.0351 |
| 2019-09-01 | 78.7881 |
| 2019-10-01 | 78.6756 |
| 2019-11-01 | 78.4573 |
| 2019-12-01 | 77.0595 |
| 2020-01-01 | 77.9406 |
| 2020-02-01 | 77.9552 |
| 2020-03-01 | 77.8979 |
| 2020-04-01 | 74.1126 |
| 2020-05-01 | 68.7213 |
| 2020-06-01 | 66.5615 |
| 2020-07-01 | 67.638 |
| 2020-08-01 | 66.6951 |
| 2020-09-01 | 67.9278 |
| 2020-10-01 | 70.4864 |
| 2020-11-01 | 70.3439 |
| 2020-12-01 | 72.5548 |
| 2021-01-01 | 71.0557 |
| 2021-02-01 | 70.0543 |
| 2021-03-01 | 74.447 |
| 2021-04-01 | 78.9508 |
| 2021-05-01 | 83.5984 |
| 2021-06-01 | 86.4325 |
| 2021-07-01 | 85.1236 |
| 2021-08-01 | 85.9692 |
| 2021-09-01 | 86.2015 |
| 2021-10-01 | 82.1054 |
| 2021-11-01 | 80.7251 |
| 2021-12-01 | 81.6126 |
| 2022-01-01 | 82.2468 |
| 2022-02-01 | 83.2799 |
| 2022-03-01 | 80.3003 |
| 2022-04-01 | 80.6566 |
| 2022-05-01 | 81.4376 |
| 2022-06-01 | 80.5495 |
| 2022-07-01 | 81.2443 |
| 2022-08-01 | 83.7803 |
| 2022-09-01 | 87.2907 |
| 2022-10-01 | 91.6848 |
| 2022-11-01 | 91.8585 |
| 2022-12-01 | 90.3548 |
| 2023-01-01 | 92.1096 |
| 2023-02-01 | 94.0157 |
| 2023-03-01 | 95.6074 |
| 2023-04-01 | 95.1756 |
| 2023-05-01 | 96.1132 |
| 2023-06-01 | 99.7914 |
| 2023-07-01 | 102.689 |
| 2023-08-01 | 103.167 |
| 2023-09-01 | 103.381 |
| 2023-10-01 | 101.595 |
| 2023-11-01 | 104.278 |
| 2023-12-01 | 104.49 |
| 2024-01-01 | 105.123 |
| 2024-02-01 | 103.377 |
| 2024-03-01 | 101.606 |
| 2024-04-01 | 102.981 |
| 2024-05-01 | 103.419 |
| 2024-06-01 | 98.5932 |
| 2024-07-01 | 98.1461 |
| 2024-08-01 | 98.2409 |
| 2024-09-01 | 94.7287 |
| 2024-10-01 | 94.8173 |
| 2024-11-01 | 95.1279 |
| 2024-12-01 | 95.6313 |

|  | Births |
| --- | --- |
| 2015-07-01 | 261.192 |
| 2015-08-01 | 259.825 |
| 2015-09-01 | 257.198 |
| 2015-10-01 | 256.961 |
| 2015-11-01 | 257.723 |
| 2015-12-01 | 258.099 |
| 2016-01-01 | 261.855 |
| 2016-02-01 | 261.163 |
| 2016-03-01 | 260.014 |
| 2016-04-01 | 259.866 |
| 2016-05-01 | 256.359 |
| 2016-06-01 | 255.118 |
| 2016-07-01 | 255.125 |
| 2016-08-01 | 258.491 |
| 2016-09-01 | 260.457 |
| 2016-10-01 | 259.34 |
| 2016-11-01 | 261.422 |
| 2016-12-01 | 263.502 |
| 2017-01-01 | 261.771 |
| 2017-02-01 | 261.682 |
| 2017-03-01 | 258.316 |
| 2017-04-01 | 259.108 |
| 2017-05-01 | 258.669 |
| 2017-06-01 | 256.422 |
| 2017-07-01 | 255.773 |
| 2017-08-01 | 252.555 |
| 2017-09-01 | 253.92 |
| 2017-10-01 | 252.216 |
| 2017-11-01 | 250.41 |
| 2017-12-01 | 247.065 |
| 2018-01-01 | 251.424 |
| 2018-02-01 | 251.896 |
| 2018-03-01 | 254.063 |
| 2018-04-01 | 254.074 |
| 2018-05-01 | 254.274 |
| 2018-06-01 | 257.179 |
| 2018-07-01 | 257.008 |
| 2018-08-01 | 258.732 |
| 2018-09-01 | 258.082 |
| 2018-10-01 | 260.649 |
| 2018-11-01 | 260.86 |
| 2018-12-01 | 263.194 |
| 2019-01-01 | 262.465 |
| 2019-02-01 | 263.688 |
| 2019-03-01 | 261.397 |
| 2019-04-01 | 261.797 |
| 2019-05-01 | 261.722 |
| 2019-06-01 | 258.521 |
| 2019-07-01 | 258.662 |
| 2019-08-01 | 257.592 |
| 2019-09-01 | 256.612 |
| 2019-10-01 | 257.345 |
| 2019-11-01 | 256.927 |
| 2019-12-01 | 253.487 |
| 2020-01-01 | 251.576 |
| 2020-02-01 | 249.852 |
| 2020-03-01 | 247.464 |
| 2020-04-01 | 246.871 |
| 2020-05-01 | 247.117 |
| 2020-06-01 | 249.085 |
| 2020-07-01 | 250.296 |
| 2020-08-01 | 252.869 |
| 2020-09-01 | 255.095 |
| 2020-10-01 | 253.508 |
| 2020-11-01 | 256.334 |
| 2020-12-01 | 257.479 |
| 2021-01-01 | 259.128 |
| 2021-02-01 | 261.739 |
| 2021-03-01 | 268.486 |
| 2021-04-01 | 268.516 |
| 2021-05-01 | 269.494 |
| 2021-06-01 | 271.129 |
| 2021-07-01 | 271.757 |
| 2021-08-01 | 271.436 |
| 2021-09-01 | 271.91 |
| 2021-10-01 | 273.248 |
| 2021-11-01 | 273.265 |
| 2021-12-01 | 276.174 |
| 2022-01-01 | 273.815 |
| 2022-02-01 | 273.834 |
| 2022-03-01 | 271.894 |
| 2022-04-01 | 269.952 |
| 2022-05-01 | 269.051 |
| 2022-06-01 | 266.135 |
| 2022-07-01 | 264.474 |
| 2022-08-01 | 262.317 |
| 2022-09-01 | 258.869 |
| 2022-10-01 | 258.725 |
| 2022-11-01 | 256.245 |
| 2022-12-01 | 255.01 |
| 2023-01-01 | 258.098 |
| 2023-02-01 | 254.142 |
| 2023-03-01 | 251.637 |
| 2023-04-01 | 253.364 |
| 2023-05-01 | 251.395 |
| 2023-06-01 | 250.599 |
| 2023-07-01 | 251.657 |
| 2023-08-01 | 250.718 |
| 2023-09-01 | 250.081 |
| 2023-10-01 | 249.023 |
| 2023-11-01 | 249.184 |
| 2023-12-01 | 246.097 |
| 2024-01-01 | 241.176 |
| 2024-02-01 | 239.538 |
| 2024-03-01 | 228.308 |

Western Australia

Rolling average vasectomies and births per 100,000 male and female population respectively for 18-24 year olds from April 2016 to December 2024

Vasectomies per 100,000 male population

Births per 100,000 female population

| Month and year | Vasectomies |
| --- | --- |
| 2016-04-01 | 0.244885 |
| 2016-05-01 | 0.222622 |
| 2016-06-01 | 0.204071 |
| 2016-07-01 | 0.204071 |
| 2016-08-01 | 0.204071 |
| 2016-09-01 | 0.204071 |
| 2016-10-01 | 0.204071 |
| 2016-11-01 | 0.136023 |
| 2016-12-01 | 0.205008 |
| 2017-01-01 | 0.0689851 |
| 2017-02-01 | 0.0689851 |
| 2017-03-01 | 0.137977 |
| 2017-04-01 | 0.207379 |
| 2017-05-01 | 0.207379 |
| 2017-06-01 | 0.276782 |
| 2017-07-01 | 0.276782 |
| 2017-08-01 | 0.346345 |
| 2017-09-01 | 0.346345 |
| 2017-10-01 | 0.346345 |
| 2017-11-01 | 0.346345 |
| 2017-12-01 | 0.27736 |
| 2018-01-01 | 0.27736 |
| 2018-02-01 | 0.27736 |
| 2018-03-01 | 0.208369 |
| 2018-04-01 | 0.138966 |
| 2018-05-01 | 0.138966 |
| 2018-06-01 | 0.0695633 |
| 2018-07-01 | 0.0695633 |
| 2018-08-01 | 0.139044 |
| 2018-09-01 | 0.139044 |
| 2018-10-01 | 0.278407 |
| 2018-11-01 | 0.41777 |
| 2018-12-01 | 0.487451 |
| 2019-01-01 | 0.487451 |
| 2019-02-01 | 0.487451 |
| 2019-03-01 | 0.556662 |
| 2019-04-01 | 0.556662 |
| 2019-05-01 | 0.833844 |
| 2019-06-01 | 0.833844 |
| 2019-07-01 | 0.833844 |
| 2019-08-01 | 0.694799 |
| 2019-09-01 | 0.763872 |
| 2019-10-01 | 0.624509 |
| 2019-11-01 | 0.554518 |
| 2019-12-01 | 0.554209 |
| 2020-01-01 | 0.554209 |
| 2020-02-01 | 0.554209 |
| 2020-03-01 | 0.553978 |
| 2020-04-01 | 0.553978 |
| 2020-05-01 | 0.276796 |
| 2020-06-01 | 0.346271 |
| 2020-07-01 | 0.346271 |
| 2020-08-01 | 0.346271 |
| 2020-09-01 | 0.277198 |
| 2020-10-01 | 0.277198 |
| 2020-11-01 | 0.278216 |
| 2020-12-01 | 0.279233 |
| 2021-01-01 | 0.349909 |
| 2021-02-01 | 0.420585 |
| 2021-03-01 | 0.351605 |
| 2021-04-01 | 0.351605 |
| 2021-05-01 | 0.422596 |
| 2021-06-01 | 0.424113 |
| 2021-07-01 | 0.424113 |
| 2021-08-01 | 0.495196 |
| 2021-09-01 | 0.495196 |
| 2021-10-01 | 0.566267 |
| 2021-11-01 | 0.70909 |
| 2021-12-01 | 0.780843 |
| 2022-01-01 | 0.78054 |
| 2022-02-01 | 0.709864 |
| 2022-03-01 | 0.709864 |
| 2022-04-01 | 0.709864 |
| 2022-05-01 | 0.638872 |
| 2022-06-01 | 0.567881 |
| 2022-07-01 | 0.636504 |
| 2022-08-01 | 0.56542 |
| 2022-09-01 | 0.634043 |
| 2022-10-01 | 0.698104 |
| 2022-11-01 | 0.620024 |
| 2022-12-01 | 0.613015 |
| 2023-01-01 | 0.608573 |
| 2023-02-01 | 0.608573 |
| 2023-03-01 | 0.608573 |
| 2023-04-01 | 0.608573 |
| 2023-05-01 | 0.608573 |
| 2023-06-01 | 0.608573 |
| 2023-07-01 | 0.53995 |
| 2023-08-01 | 0.53995 |
| 2023-09-01 | 0.599026 |
| 2023-10-01 | 0.653764 |
| 2023-11-01 | 0.518631 |
| 2023-12-01 | 0.446789 |
| 2024-01-01 | 0.380858 |
| 2024-02-01 | 0.380858 |
| 2024-03-01 | 0.50529 |
| 2024-04-01 | 0.50529 |
| 2024-05-01 | 0.50529 |
| 2024-06-01 | 0.50529 |
| 2024-07-01 | 0.50529 |
| 2024-08-01 | 0.627719 |
| 2024-09-01 | 0.561235 |
| 2024-10-01 | 0.371365 |
| 2024-11-01 | 0.432332 |
| 2024-12-01 | 0.490978 |

|  | Births |
| --- | --- |
| 2015-07-01 | 362.73 |
| 2015-08-01 | 362.957 |
| 2015-09-01 | 363.218 |
| 2015-10-01 | 361.564 |
| 2015-11-01 | 359.334 |
| 2015-12-01 | 358.744 |
| 2016-01-01 | 359.106 |
| 2016-02-01 | 358.499 |
| 2016-03-01 | 360.841 |
| 2016-04-01 | 359.34 |
| 2016-05-01 | 358.276 |
| 2016-06-01 | 356.994 |
| 2016-07-01 | 351.395 |
| 2016-08-01 | 352.165 |
| 2016-09-01 | 349.412 |
| 2016-10-01 | 347.339 |
| 2016-11-01 | 345.338 |
| 2016-12-01 | 344.653 |
| 2017-01-01 | 341.85 |
| 2017-02-01 | 336.884 |
| 2017-03-01 | 336.417 |
| 2017-04-01 | 334.23 |
| 2017-05-01 | 331.824 |
| 2017-06-01 | 329.933 |
| 2017-07-01 | 327.111 |
| 2017-08-01 | 322.4 |
| 2017-09-01 | 321.213 |
| 2017-10-01 | 320.035 |
| 2017-11-01 | 318.415 |
| 2017-12-01 | 316.062 |
| 2018-01-01 | 313.906 |
| 2018-02-01 | 314.255 |
| 2018-03-01 | 310.026 |
| 2018-04-01 | 308.601 |
| 2018-05-01 | 303.862 |
| 2018-06-01 | 301.111 |
| 2018-07-01 | 302.836 |
| 2018-08-01 | 300.272 |
| 2018-09-01 | 298.299 |
| 2018-10-01 | 298.142 |
| 2018-11-01 | 298.428 |
| 2018-12-01 | 295.846 |
| 2019-01-01 | 294.574 |
| 2019-02-01 | 296.406 |
| 2019-03-01 | 295.438 |
| 2019-04-01 | 293.374 |
| 2019-05-01 | 291.393 |
| 2019-06-01 | 290.286 |
| 2019-07-01 | 287.512 |
| 2019-08-01 | 287.547 |
| 2019-09-01 | 285.807 |
| 2019-10-01 | 286.038 |
| 2019-11-01 | 283.962 |
| 2019-12-01 | 283.266 |
| 2020-01-01 | 281.079 |
| 2020-02-01 | 276.067 |
| 2020-03-01 | 276.241 |
| 2020-04-01 | 276.845 |
| 2020-05-01 | 278.656 |
| 2020-06-01 | 280.239 |
| 2020-07-01 | 281.086 |
| 2020-08-01 | 279.913 |
| 2020-09-01 | 280.302 |
| 2020-10-01 | 277.977 |
| 2020-11-01 | 278.032 |
| 2020-12-01 | 278.969 |
| 2021-01-01 | 281.759 |
| 2021-02-01 | 282.243 |
| 2021-03-01 | 283.515 |
| 2021-04-01 | 281.249 |
| 2021-05-01 | 277.547 |
| 2021-06-01 | 275.209 |
| 2021-07-01 | 273.676 |
| 2021-08-01 | 273.799 |
| 2021-09-01 | 270.474 |
| 2021-10-01 | 266.957 |
| 2021-11-01 | 264.472 |
| 2021-12-01 | 263.257 |
| 2022-01-01 | 257.991 |
| 2022-02-01 | 256.311 |
| 2022-03-01 | 252.923 |
| 2022-04-01 | 248.803 |
| 2022-05-01 | 247.853 |
| 2022-06-01 | 243.821 |
| 2022-07-01 | 238.703 |
| 2022-08-01 | 234.051 |
| 2022-09-01 | 233.103 |
| 2022-10-01 | 232.4 |
| 2022-11-01 | 229.067 |
| 2022-12-01 | 224.933 |
| 2023-01-01 | 222.996 |
| 2023-02-01 | 220.872 |
| 2023-03-01 | 216.841 |
| 2023-04-01 | 216.879 |
| 2023-05-01 | 216.246 |
| 2023-06-01 | 216.698 |
| 2023-07-01 | 215.834 |
| 2023-08-01 | 214.114 |
| 2023-09-01 | 211.753 |
| 2023-10-01 | 209.471 |
| 2023-11-01 | 205.779 |
| 2023-12-01 | 200.956 |
| 2024-01-01 | 194.954 |
| 2024-02-01 | 184.912 |
| 2024-03-01 | 168.321 |

Rolling average vasectomies and births per 100,000 male and female population respectively for 25-34 year olds from April 2016 to December 2024

Vasectomies per 100,000 male population

Births per 100,000 female population

| Month and year | Vasectomies |
| --- | --- |
| 2016-04-01 | 21.8592 |
| 2016-05-01 | 21.1984 |
| 2016-06-01 | 21.0936 |
| 2016-07-01 | 21.3968 |
| 2016-08-01 | 20.9363 |
| 2016-09-01 | 20.7955 |
| 2016-10-01 | 21.0606 |
| 2016-11-01 | 21.3271 |
| 2016-12-01 | 21.0243 |
| 2017-01-01 | 20.7973 |
| 2017-02-01 | 21.1398 |
| 2017-03-01 | 20.7993 |
| 2017-04-01 | 20.535 |
| 2017-05-01 | 20.9608 |
| 2017-06-01 | 21.6794 |
| 2017-07-01 | 21.1315 |
| 2017-08-01 | 21.2835 |
| 2017-09-01 | 21.8453 |
| 2017-10-01 | 21.8326 |
| 2017-11-01 | 21.5733 |
| 2017-12-01 | 21.4736 |
| 2018-01-01 | 22.1566 |
| 2018-02-01 | 22.0153 |
| 2018-03-01 | 22.0438 |
| 2018-04-01 | 22.1953 |
| 2018-05-01 | 22.1812 |
| 2018-06-01 | 21.4285 |
| 2018-07-01 | 22.2438 |
| 2018-08-01 | 22.4834 |
| 2018-09-01 | 21.1378 |
| 2018-10-01 | 21.9195 |
| 2018-11-01 | 22.3218 |
| 2018-12-01 | 22.6368 |
| 2019-01-01 | 22.617 |
| 2019-02-01 | 23.1377 |
| 2019-03-01 | 22.7816 |
| 2019-04-01 | 23.3014 |
| 2019-05-01 | 23.9046 |
| 2019-06-01 | 23.7923 |
| 2019-07-01 | 23.6811 |
| 2019-08-01 | 23.3601 |
| 2019-09-01 | 24.5874 |
| 2019-10-01 | 24.0905 |
| 2019-11-01 | 24.8127 |
| 2019-12-01 | 24.65 |
| 2020-01-01 | 24.3142 |
| 2020-02-01 | 24.0203 |
| 2020-03-01 | 24.7339 |
| 2020-04-01 | 23.5569 |
| 2020-05-01 | 22.8858 |
| 2020-06-01 | 23.8576 |
| 2020-07-01 | 24.8031 |
| 2020-08-01 | 26.002 |
| 2020-09-01 | 26.8209 |
| 2020-10-01 | 27.3526 |
| 2020-11-01 | 28.1888 |
| 2020-12-01 | 29.0555 |
| 2021-01-01 | 29.546 |
| 2021-02-01 | 29.8253 |
| 2021-03-01 | 30.5382 |
| 2021-04-01 | 31.5732 |
| 2021-05-01 | 31.7188 |
| 2021-06-01 | 31.9175 |
| 2021-07-01 | 31.1273 |
| 2021-08-01 | 30.5085 |
| 2021-09-01 | 30.4001 |
| 2021-10-01 | 31.2568 |
| 2021-11-01 | 30.3697 |
| 2021-12-01 | 29.6932 |
| 2022-01-01 | 29.5969 |
| 2022-02-01 | 30.0514 |
| 2022-03-01 | 29.2304 |
| 2022-04-01 | 28.8208 |
| 2022-05-01 | 29.2148 |
| 2022-06-01 | 28.8802 |
| 2022-07-01 | 28.6431 |
| 2022-08-01 | 29.1814 |
| 2022-09-01 | 28.9657 |
| 2022-10-01 | 29.1154 |
| 2022-11-01 | 29.7458 |
| 2022-12-01 | 30.1802 |
| 2023-01-01 | 30.249 |
| 2023-02-01 | 29.9311 |
| 2023-03-01 | 31.1745 |
| 2023-04-01 | 31.8602 |
| 2023-05-01 | 33.0566 |
| 2023-06-01 | 33.5746 |
| 2023-07-01 | 35.121 |
| 2023-08-01 | 34.9855 |
| 2023-09-01 | 35.415 |
| 2023-10-01 | 34.8304 |
| 2023-11-01 | 34.7137 |
| 2023-12-01 | 34.6484 |
| 2024-01-01 | 34.6585 |
| 2024-02-01 | 34.7737 |
| 2024-03-01 | 32.9977 |
| 2024-04-01 | 32.275 |
| 2024-05-01 | 30.8043 |
| 2024-06-01 | 31.1835 |
| 2024-07-01 | 29.8838 |
| 2024-08-01 | 29.8424 |
| 2024-09-01 | 29.3961 |
| 2024-10-01 | 28.6806 |
| 2024-11-01 | 28.3485 |
| 2024-12-01 | 28.9614 |

|  | Births |
| --- | --- |
| 2015-07-01 | 943.972 |
| 2015-08-01 | 944.423 |
| 2015-09-01 | 944.923 |
| 2015-10-01 | 944.367 |
| 2015-11-01 | 947.439 |
| 2015-12-01 | 946.998 |
| 2016-01-01 | 944.534 |
| 2016-02-01 | 943.524 |
| 2016-03-01 | 941.138 |
| 2016-04-01 | 939.075 |
| 2016-05-01 | 928.128 |
| 2016-06-01 | 925.466 |
| 2016-07-01 | 919.324 |
| 2016-08-01 | 916.157 |
| 2016-09-01 | 917.72 |
| 2016-10-01 | 913.177 |
| 2016-11-01 | 910.697 |
| 2016-12-01 | 903.584 |
| 2017-01-01 | 899.888 |
| 2017-02-01 | 902.382 |
| 2017-03-01 | 903.035 |
| 2017-04-01 | 904.472 |
| 2017-05-01 | 904.164 |
| 2017-06-01 | 900.513 |
| 2017-07-01 | 905.862 |
| 2017-08-01 | 907.361 |
| 2017-09-01 | 897.363 |
| 2017-10-01 | 899.942 |
| 2017-11-01 | 897.538 |
| 2017-12-01 | 896.724 |
| 2018-01-01 | 896.59 |
| 2018-02-01 | 893.807 |
| 2018-03-01 | 888.111 |
| 2018-04-01 | 886.551 |
| 2018-05-01 | 885.432 |
| 2018-06-01 | 883.625 |
| 2018-07-01 | 876.764 |
| 2018-08-01 | 875.761 |
| 2018-09-01 | 878.198 |
| 2018-10-01 | 876.487 |
| 2018-11-01 | 873.008 |
| 2018-12-01 | 874.748 |
| 2019-01-01 | 877.459 |
| 2019-02-01 | 873.401 |
| 2019-03-01 | 873.391 |
| 2019-04-01 | 872.895 |
| 2019-05-01 | 874.438 |
| 2019-06-01 | 868.819 |
| 2019-07-01 | 864.913 |
| 2019-08-01 | 860.122 |
| 2019-09-01 | 855.627 |
| 2019-10-01 | 853.575 |
| 2019-11-01 | 850.45 |
| 2019-12-01 | 846.891 |
| 2020-01-01 | 838.214 |
| 2020-02-01 | 838.252 |
| 2020-03-01 | 837.571 |
| 2020-04-01 | 840.8 |
| 2020-05-01 | 846.776 |
| 2020-06-01 | 858.25 |
| 2020-07-01 | 869.289 |
| 2020-08-01 | 874.034 |
| 2020-09-01 | 881.09 |
| 2020-10-01 | 884.498 |
| 2020-11-01 | 889.754 |
| 2020-12-01 | 895.956 |
| 2021-01-01 | 903.863 |
| 2021-02-01 | 908.219 |
| 2021-03-01 | 912.828 |
| 2021-04-01 | 908.537 |
| 2021-05-01 | 901.974 |
| 2021-06-01 | 895.182 |
| 2021-07-01 | 885.995 |
| 2021-08-01 | 879.405 |
| 2021-09-01 | 873.284 |
| 2021-10-01 | 865.777 |
| 2021-11-01 | 864.894 |
| 2021-12-01 | 855.842 |
| 2022-01-01 | 847.497 |
| 2022-02-01 | 839.328 |
| 2022-03-01 | 830.498 |
| 2022-04-01 | 823.323 |
| 2022-05-01 | 813.132 |
| 2022-06-01 | 799.513 |
| 2022-07-01 | 790.812 |
| 2022-08-01 | 786.634 |
| 2022-09-01 | 776.14 |
| 2022-10-01 | 771.082 |
| 2022-11-01 | 760.586 |
| 2022-12-01 | 753.079 |
| 2023-01-01 | 747.996 |
| 2023-02-01 | 742.085 |
| 2023-03-01 | 732.746 |
| 2023-04-01 | 726.615 |
| 2023-05-01 | 726.115 |
| 2023-06-01 | 723.207 |
| 2023-07-01 | 720.505 |
| 2023-08-01 | 719.901 |
| 2023-09-01 | 719.363 |
| 2023-10-01 | 715.902 |
| 2023-11-01 | 710.559 |
| 2023-12-01 | 704.664 |
| 2024-01-01 | 697.721 |
| 2024-02-01 | 669.957 |
| 2024-03-01 | 616.28 |

Rolling average vasectomies and births per 100,000 male and female population respectively for 35-44 year olds from April 2016 to December 2024

Vasectomies per 100,000 male population

Births per 100,000 female population

| Month and year | Vasectomies |
| --- | --- |
| 2016-04-01 | 66.1233 |
| 2016-05-01 | 66.2155 |
| 2016-06-01 | 67.8309 |
| 2016-07-01 | 68.1312 |
| 2016-08-01 | 69.9249 |
| 2016-09-01 | 72.0357 |
| 2016-10-01 | 71.6649 |
| 2016-11-01 | 72.9669 |
| 2016-12-01 | 73.3395 |
| 2017-01-01 | 73.7902 |
| 2017-02-01 | 74.9305 |
| 2017-03-01 | 75.8857 |
| 2017-04-01 | 76.2669 |
| 2017-05-01 | 76.8333 |
| 2017-06-01 | 76.3708 |
| 2017-07-01 | 77.0621 |
| 2017-08-01 | 75.4707 |
| 2017-09-01 | 75.0028 |
| 2017-10-01 | 75.5514 |
| 2017-11-01 | 74.4226 |
| 2017-12-01 | 73.3088 |
| 2018-01-01 | 75.0085 |
| 2018-02-01 | 74.4801 |
| 2018-03-01 | 75.1482 |
| 2018-04-01 | 76.6286 |
| 2018-05-01 | 76.4599 |
| 2018-06-01 | 77.5656 |
| 2018-07-01 | 77.8451 |
| 2018-08-01 | 78.7128 |
| 2018-09-01 | 78.4527 |
| 2018-10-01 | 78.6275 |
| 2018-11-01 | 79.0661 |
| 2018-12-01 | 79.8935 |
| 2019-01-01 | 80.6365 |
| 2019-02-01 | 81.3745 |
| 2019-03-01 | 82.0064 |
| 2019-04-01 | 81.5858 |
| 2019-05-01 | 82.5842 |
| 2019-06-01 | 81.1712 |
| 2019-07-01 | 82.1785 |
| 2019-08-01 | 82.3653 |
| 2019-09-01 | 82.9513 |
| 2019-10-01 | 83.8095 |
| 2019-11-01 | 85.1342 |
| 2019-12-01 | 85.9607 |
| 2020-01-01 | 84.8107 |
| 2020-02-01 | 85.5388 |
| 2020-03-01 | 85.2378 |
| 2020-04-01 | 81.0365 |
| 2020-05-01 | 77.447 |
| 2020-06-01 | 78.3225 |
| 2020-07-01 | 80.1634 |
| 2020-08-01 | 80.6159 |
| 2020-09-01 | 82.0179 |
| 2020-10-01 | 84.0336 |
| 2020-11-01 | 85.6305 |
| 2020-12-01 | 88.0902 |
| 2021-01-01 | 88.9724 |
| 2021-02-01 | 88.2618 |
| 2021-03-01 | 90.1655 |
| 2021-04-01 | 93.527 |
| 2021-05-01 | 98.9952 |
| 2021-06-01 | 100.414 |
| 2021-07-01 | 97.8435 |
| 2021-08-01 | 98.9388 |
| 2021-09-01 | 99.9223 |
| 2021-10-01 | 98.845 |
| 2021-11-01 | 97.1173 |
| 2021-12-01 | 95.0355 |
| 2022-01-01 | 94.9877 |
| 2022-02-01 | 95.2169 |
| 2022-03-01 | 93.8443 |
| 2022-04-01 | 92.5156 |
| 2022-05-01 | 89.1732 |
| 2022-06-01 | 87.6516 |
| 2022-07-01 | 88.0755 |
| 2022-08-01 | 87.0756 |
| 2022-09-01 | 86.9142 |
| 2022-10-01 | 87.407 |
| 2022-11-01 | 88.4982 |
| 2022-12-01 | 87.2591 |
| 2023-01-01 | 86.2077 |
| 2023-02-01 | 86.1924 |
| 2023-03-01 | 87.4156 |
| 2023-04-01 | 90.1231 |
| 2023-05-01 | 94.1329 |
| 2023-06-01 | 96.7303 |
| 2023-07-01 | 98.0313 |
| 2023-08-01 | 100.101 |
| 2023-09-01 | 101.698 |
| 2023-10-01 | 101.963 |
| 2023-11-01 | 103.225 |
| 2023-12-01 | 104.818 |
| 2024-01-01 | 107.589 |
| 2024-02-01 | 108.15 |
| 2024-03-01 | 106.004 |
| 2024-04-01 | 105.001 |
| 2024-05-01 | 103.537 |
| 2024-06-01 | 102.966 |
| 2024-07-01 | 102.855 |
| 2024-08-01 | 102.716 |
| 2024-09-01 | 102.143 |
| 2024-10-01 | 101.796 |
| 2024-11-01 | 100.129 |
| 2024-12-01 | 100.13 |

|  | Births |
| --- | --- |
| 2015-07-01 | 285.107 |
| 2015-08-01 | 286.814 |
| 2015-09-01 | 287.185 |
| 2015-10-01 | 286.348 |
| 2015-11-01 | 285.655 |
| 2015-12-01 | 285.772 |
| 2016-01-01 | 288.29 |
| 2016-02-01 | 290.713 |
| 2016-03-01 | 290.943 |
| 2016-04-01 | 289.667 |
| 2016-05-01 | 288.726 |
| 2016-06-01 | 290 |
| 2016-07-01 | 289.058 |
| 2016-08-01 | 287.122 |
| 2016-09-01 | 286.805 |
| 2016-10-01 | 288.764 |
| 2016-11-01 | 288.175 |
| 2016-12-01 | 287.242 |
| 2017-01-01 | 288.171 |
| 2017-02-01 | 287.415 |
| 2017-03-01 | 284.878 |
| 2017-04-01 | 287.162 |
| 2017-05-01 | 287.445 |
| 2017-06-01 | 281.863 |
| 2017-07-01 | 278.455 |
| 2017-08-01 | 278.826 |
| 2017-09-01 | 277.004 |
| 2017-10-01 | 274.692 |
| 2017-11-01 | 275.192 |
| 2017-12-01 | 274.426 |
| 2018-01-01 | 274.143 |
| 2018-02-01 | 275.233 |
| 2018-03-01 | 275.631 |
| 2018-04-01 | 273.252 |
| 2018-05-01 | 273.25 |
| 2018-06-01 | 274.729 |
| 2018-07-01 | 277.916 |
| 2018-08-01 | 279.949 |
| 2018-09-01 | 281.253 |
| 2018-10-01 | 284.295 |
| 2018-11-01 | 283.549 |
| 2018-12-01 | 283.214 |
| 2019-01-01 | 284.008 |
| 2019-02-01 | 282.768 |
| 2019-03-01 | 282.628 |
| 2019-04-01 | 282.345 |
| 2019-05-01 | 281.797 |
| 2019-06-01 | 282.91 |
| 2019-07-01 | 282.807 |
| 2019-08-01 | 280.636 |
| 2019-09-01 | 280.467 |
| 2019-10-01 | 276.688 |
| 2019-11-01 | 277.532 |
| 2019-12-01 | 276.891 |
| 2020-01-01 | 276.297 |
| 2020-02-01 | 274.866 |
| 2020-03-01 | 273.433 |
| 2020-04-01 | 273.421 |
| 2020-05-01 | 276.55 |
| 2020-06-01 | 279.189 |
| 2020-07-01 | 280.194 |
| 2020-08-01 | 282.745 |
| 2020-09-01 | 285.094 |
| 2020-10-01 | 287.209 |
| 2020-11-01 | 286.78 |
| 2020-12-01 | 289.273 |
| 2021-01-01 | 289.46 |
| 2021-02-01 | 291.211 |
| 2021-03-01 | 294.772 |
| 2021-04-01 | 295.684 |
| 2021-05-01 | 294.136 |
| 2021-06-01 | 292.04 |
| 2021-07-01 | 289.897 |
| 2021-08-01 | 289.382 |
| 2021-09-01 | 285.194 |
| 2021-10-01 | 283.698 |
| 2021-11-01 | 285.231 |
| 2021-12-01 | 284.499 |
| 2022-01-01 | 281.256 |
| 2022-02-01 | 281.736 |
| 2022-03-01 | 278.752 |
| 2022-04-01 | 277.581 |
| 2022-05-01 | 271.694 |
| 2022-06-01 | 268.33 |
| 2022-07-01 | 265.79 |
| 2022-08-01 | 262.619 |
| 2022-09-01 | 260.419 |
| 2022-10-01 | 259.881 |
| 2022-11-01 | 256.624 |
| 2022-12-01 | 254.892 |
| 2023-01-01 | 256.193 |
| 2023-02-01 | 253.874 |
| 2023-03-01 | 254.099 |
| 2023-04-01 | 253.932 |
| 2023-05-01 | 256.711 |
| 2023-06-01 | 257.145 |
| 2023-07-01 | 257.619 |
| 2023-08-01 | 257.707 |
| 2023-09-01 | 259.065 |
| 2023-10-01 | 259.235 |
| 2023-11-01 | 259.847 |
| 2023-12-01 | 256.488 |
| 2024-01-01 | 252.062 |
| 2024-02-01 | 242.613 |
| 2024-03-01 | 221.856 |

Tasmania:

Rolling average vasectomies and births per 100,000 male and female population respectively for 18-24 year olds from April 2016 to December 2024

Vasectomies per 100,000 male population

Births per 100,000 female population

| Month and year | Vasectomies |
| --- | --- |
| 2016-04-01 | 0 |
| 2016-05-01 | 0 |
| 2016-06-01 | 0 |
| 2016-07-01 | 0 |
| 2016-08-01 | 0.359211 |
| 2016-09-01 | 0.359211 |
| 2016-10-01 | 0.359211 |
| 2016-11-01 | 0.717172 |
| 2016-12-01 | 0.717172 |
| 2017-01-01 | 0.717172 |
| 2017-02-01 | 0.717172 |
| 2017-03-01 | 0.717172 |
| 2017-04-01 | 0.717172 |
| 2017-05-01 | 0.717172 |
| 2017-06-01 | 1.06967 |
| 2017-07-01 | 1.06967 |
| 2017-08-01 | 0.710456 |
| 2017-09-01 | 0.710456 |
| 2017-10-01 | 1.05909 |
| 2017-11-01 | 0.701126 |
| 2017-12-01 | 0.701126 |
| 2018-01-01 | 1.04839 |
| 2018-02-01 | 1.04839 |
| 2018-03-01 | 1.04839 |
| 2018-04-01 | 1.04839 |
| 2018-05-01 | 1.04839 |
| 2018-06-01 | 1.04257 |
| 2018-07-01 | 1.04257 |
| 2018-08-01 | 1.04257 |
| 2018-09-01 | 1.04257 |
| 2018-10-01 | 0.693939 |
| 2018-11-01 | 1.38931 |
| 2018-12-01 | 1.38931 |
| 2019-01-01 | 1.04204 |
| 2019-02-01 | 1.04204 |
| 2019-03-01 | 1.04204 |
| 2019-04-01 | 1.04204 |
| 2019-05-01 | 1.04204 |
| 2019-06-01 | 0.695372 |
| 2019-07-01 | 0.695372 |
| 2019-08-01 | 0.695372 |
| 2019-09-01 | 1.04754 |
| 2019-10-01 | 1.40366 |
| 2019-11-01 | 0.708293 |
| 2019-12-01 | 1.06442 |
| 2020-01-01 | 1.06442 |
| 2020-02-01 | 1.06442 |
| 2020-03-01 | 1.42164 |
| 2020-04-01 | 1.42164 |
| 2020-05-01 | 1.42164 |
| 2020-06-01 | 1.42164 |
| 2020-07-01 | 1.42164 |
| 2020-08-01 | 1.42164 |
| 2020-09-01 | 1.06948 |
| 2020-10-01 | 0.71335 |
| 2020-11-01 | 1.07714 |
| 2020-12-01 | 0.721014 |
| 2021-01-01 | 0.721014 |
| 2021-02-01 | 0.721014 |
| 2021-03-01 | 0.36379 |
| 2021-04-01 | 0.36379 |
| 2021-05-01 | 0.36379 |
| 2021-06-01 | 0.36379 |
| 2021-07-01 | 0.36379 |
| 2021-08-01 | 0.36379 |
| 2021-09-01 | 0.36379 |
| 2021-10-01 | 0.36379 |
| 2021-11-01 | 0 |
| 2021-12-01 | 0.372107 |
| 2022-01-01 | 0.372107 |
| 2022-02-01 | 0.372107 |
| 2022-03-01 | 0.372107 |
| 2022-04-01 | 0.372107 |
| 2022-05-01 | 0.747313 |
| 2022-06-01 | 0.747313 |
| 2022-07-01 | 0.747313 |
| 2022-08-01 | 0.747313 |
| 2022-09-01 | 0.747313 |
| 2022-10-01 | 1.1248 |
| 2022-11-01 | 1.50228 |
| 2022-12-01 | 1.13017 |
| 2023-01-01 | 1.13017 |
| 2023-02-01 | 1.13017 |
| 2023-03-01 | 1.50769 |
| 2023-04-01 | 1.88703 |
| 2023-05-01 | 1.89117 |
| 2023-06-01 | 1.89117 |
| 2023-07-01 | 2.27002 |
| 2023-08-01 | 2.27002 |
| 2023-09-01 | 2.27002 |
| 2023-10-01 | 1.89254 |
| 2023-11-01 | 1.89479 |
| 2023-12-01 | 1.89479 |
| 2024-01-01 | 1.89479 |
| 2024-02-01 | 1.89479 |
| 2024-03-01 | 1.51727 |
| 2024-04-01 | 1.13793 |
| 2024-05-01 | 0.758594 |
| 2024-06-01 | 0.758594 |
| 2024-07-01 | 0.379737 |
| 2024-08-01 | 0.379737 |
| 2024-09-01 | 0.757495 |
| 2024-10-01 | 0.757495 |
| 2024-11-01 | 0.377758 |
| 2024-12-01 | 0.377758 |

|  | Births |
| --- | --- |
| 2015-07-01 | 476.372 |
| 2015-08-01 | 476.943 |
| 2015-09-01 | 478.997 |
| 2015-10-01 | 468.676 |
| 2015-11-01 | 471.647 |
| 2015-12-01 | 474.633 |
| 2016-01-01 | 473.379 |
| 2016-02-01 | 476.427 |
| 2016-03-01 | 473.624 |
| 2016-04-01 | 466.961 |
| 2016-05-01 | 449.937 |
| 2016-06-01 | 437.16 |
| 2016-07-01 | 438.068 |
| 2016-08-01 | 434.701 |
| 2016-09-01 | 429.373 |
| 2016-10-01 | 421.207 |
| 2016-11-01 | 410.331 |
| 2016-12-01 | 406.779 |
| 2017-01-01 | 405.724 |
| 2017-02-01 | 398.527 |
| 2017-03-01 | 403.988 |
| 2017-04-01 | 398.414 |
| 2017-05-01 | 406.27 |
| 2017-06-01 | 414.441 |
| 2017-07-01 | 402.826 |
| 2017-08-01 | 396.155 |
| 2017-09-01 | 395.528 |
| 2017-10-01 | 396.171 |
| 2017-11-01 | 402.824 |
| 2017-12-01 | 394.803 |
| 2018-01-01 | 395.051 |
| 2018-02-01 | 398.359 |
| 2018-03-01 | 388.028 |
| 2018-04-01 | 389.142 |
| 2018-05-01 | 380.154 |
| 2018-06-01 | 381.946 |
| 2018-07-01 | 383.651 |
| 2018-08-01 | 390.59 |
| 2018-09-01 | 383.619 |
| 2018-10-01 | 380.405 |
| 2018-11-01 | 372.696 |
| 2018-12-01 | 374.349 |
| 2019-01-01 | 365.997 |
| 2019-02-01 | 357.636 |
| 2019-03-01 | 355.669 |
| 2019-04-01 | 356.988 |
| 2019-05-01 | 358.167 |
| 2019-06-01 | 349.723 |
| 2019-07-01 | 346.55 |
| 2019-08-01 | 335.885 |
| 2019-09-01 | 330.019 |
| 2019-10-01 | 338.399 |
| 2019-11-01 | 332.531 |
| 2019-12-01 | 335.103 |
| 2020-01-01 | 340.689 |
| 2020-02-01 | 346.647 |
| 2020-03-01 | 354.568 |
| 2020-04-01 | 350.793 |
| 2020-05-01 | 353.032 |
| 2020-06-01 | 351.505 |
| 2020-07-01 | 358.561 |
| 2020-08-01 | 360.106 |
| 2020-09-01 | 362.342 |
| 2020-10-01 | 349.194 |
| 2020-11-01 | 353.911 |
| 2020-12-01 | 349.292 |
| 2021-01-01 | 348.263 |
| 2021-02-01 | 348.821 |
| 2021-03-01 | 343.079 |
| 2021-04-01 | 337.896 |
| 2021-05-01 | 331.848 |
| 2021-06-01 | 327.834 |
| 2021-07-01 | 325.768 |
| 2021-08-01 | 321.972 |
| 2021-09-01 | 324.97 |
| 2021-10-01 | 332.358 |
| 2021-11-01 | 329.699 |
| 2021-12-01 | 331.443 |
| 2022-01-01 | 329.187 |
| 2022-02-01 | 319.615 |
| 2022-03-01 | 319.766 |
| 2022-04-01 | 323.524 |
| 2022-05-01 | 323.139 |
| 2022-06-01 | 326.489 |
| 2022-07-01 | 324.427 |
| 2022-08-01 | 323.097 |
| 2022-09-01 | 318.55 |
| 2022-10-01 | 306.176 |
| 2022-11-01 | 310.939 |
| 2022-12-01 | 307.94 |
| 2023-01-01 | 316.324 |
| 2023-02-01 | 320.565 |
| 2023-03-01 | 314.178 |
| 2023-04-01 | 309.012 |
| 2023-05-01 | 306.266 |
| 2023-06-01 | 299.457 |
| 2023-07-01 | 291.938 |
| 2023-08-01 | 293.44 |
| 2023-09-01 | 295.364 |
| 2023-10-01 | 300.85 |
| 2023-11-01 | 296.522 |
| 2023-12-01 | 290.505 |
| 2024-01-01 | 277.504 |
| 2024-02-01 | 268.984 |
| 2024-03-01 | 255.488 |

Rolling average vasectomies and births per 100,000 male and female population respectively for 25-34 year olds from April 2016 to December 2024

Vasectomies per 100,000 male population

Births per 100,000 female population

| Month and year | Vasectomies |
| --- | --- |
| 2016-04-01 | 25.5756 |
| 2016-05-01 | 28.6595 |
| 2016-06-01 | 29.0258 |
| 2016-07-01 | 29.8206 |
| 2016-08-01 | 30.5777 |
| 2016-09-01 | 32.1782 |
| 2016-10-01 | 35.0826 |
| 2016-11-01 | 36.0476 |
| 2016-12-01 | 35.9717 |
| 2017-01-01 | 36.1697 |
| 2017-02-01 | 36.6439 |
| 2017-03-01 | 36.5657 |
| 2017-04-01 | 35.9078 |
| 2017-05-01 | 33.5705 |
| 2017-06-01 | 34.4852 |
| 2017-07-01 | 34.6489 |
| 2017-08-01 | 34.459 |
| 2017-09-01 | 34.5548 |
| 2017-10-01 | 34.3231 |
| 2017-11-01 | 33.5814 |
| 2017-12-01 | 34.2238 |
| 2018-01-01 | 34.8779 |
| 2018-02-01 | 34.7792 |
| 2018-03-01 | 34.9314 |
| 2018-04-01 | 36.0464 |
| 2018-05-01 | 38.1195 |
| 2018-06-01 | 37.4149 |
| 2018-07-01 | 37.0473 |
| 2018-08-01 | 36.1002 |
| 2018-09-01 | 36.6267 |
| 2018-10-01 | 34.4462 |
| 2018-11-01 | 35.8882 |
| 2018-12-01 | 36.1798 |
| 2019-01-01 | 35.3254 |
| 2019-02-01 | 36.1698 |
| 2019-03-01 | 36.7632 |
| 2019-04-01 | 36.8205 |
| 2019-05-01 | 36.096 |
| 2019-06-01 | 36.6338 |
| 2019-07-01 | 37.2403 |
| 2019-08-01 | 37.3363 |
| 2019-09-01 | 37.8325 |
| 2019-10-01 | 39.5185 |
| 2019-11-01 | 37.6405 |
| 2019-12-01 | 38.3554 |
| 2020-01-01 | 37.3704 |
| 2020-02-01 | 37.0031 |
| 2020-03-01 | 37.0827 |
| 2020-04-01 | 34.9202 |
| 2020-05-01 | 32.2785 |
| 2020-06-01 | 29.8825 |
| 2020-07-01 | 30.2021 |
| 2020-08-01 | 30.7171 |
| 2020-09-01 | 30.2735 |
| 2020-10-01 | 30.0893 |
| 2020-11-01 | 29.2815 |
| 2020-12-01 | 27.796 |
| 2021-01-01 | 29.2377 |
| 2021-02-01 | 28.9046 |
| 2021-03-01 | 30.4478 |
| 2021-04-01 | 32.451 |
| 2021-05-01 | 33.4038 |
| 2021-06-01 | 36.6509 |
| 2021-07-01 | 37.5373 |
| 2021-08-01 | 37.3536 |
| 2021-09-01 | 35.273 |
| 2021-10-01 | 34.0738 |
| 2021-11-01 | 33.9502 |
| 2021-12-01 | 36.6935 |
| 2022-01-01 | 36.8212 |
| 2022-02-01 | 37.5674 |
| 2022-03-01 | 36.194 |
| 2022-04-01 | 34.1019 |
| 2022-05-01 | 34.6707 |
| 2022-06-01 | 33.1663 |
| 2022-07-01 | 31.6974 |
| 2022-08-01 | 34.9355 |
| 2022-09-01 | 36.9602 |
| 2022-10-01 | 36.9579 |
| 2022-11-01 | 37.774 |
| 2022-12-01 | 35.1093 |
| 2023-01-01 | 34.7189 |
| 2023-02-01 | 35.7704 |
| 2023-03-01 | 37.8512 |
| 2023-04-01 | 38.9002 |
| 2023-05-01 | 39.1508 |
| 2023-06-01 | 39.8147 |
| 2023-07-01 | 41.5158 |
| 2023-08-01 | 39.9636 |
| 2023-09-01 | 38.5759 |
| 2023-10-01 | 41.5541 |
| 2023-11-01 | 42.8781 |
| 2023-12-01 | 42.7261 |
| 2024-01-01 | 44.0261 |
| 2024-02-01 | 44.9466 |
| 2024-03-01 | 43.3746 |
| 2024-04-01 | 44.2697 |
| 2024-05-01 | 44.3432 |
| 2024-06-01 | 44.4264 |
| 2024-07-01 | 44.3003 |
| 2024-08-01 | 42.7079 |
| 2024-09-01 | 48.7274 |
| 2024-10-01 | 48.423 |
| 2024-11-01 | 50.0231 |
| 2024-12-01 | 51.7895 |

|  | Births |
| --- | --- |
| 2015-07-01 | 946.668 |
| 2015-08-01 | 956.194 |
| 2015-09-01 | 952.396 |
| 2015-10-01 | 950.601 |
| 2015-11-01 | 951.821 |
| 2015-12-01 | 951.293 |
| 2016-01-01 | 937.072 |
| 2016-02-01 | 945.257 |
| 2016-03-01 | 929.048 |
| 2016-04-01 | 928.801 |
| 2016-05-01 | 920.856 |
| 2016-06-01 | 912.073 |
| 2016-07-01 | 897.787 |
| 2016-08-01 | 887.598 |
| 2016-09-01 | 870.637 |
| 2016-10-01 | 865.153 |
| 2016-11-01 | 864.554 |
| 2016-12-01 | 864.336 |
| 2017-01-01 | 860.358 |
| 2017-02-01 | 853.694 |
| 2017-03-01 | 862.798 |
| 2017-04-01 | 855.918 |
| 2017-05-01 | 847.646 |
| 2017-06-01 | 841.68 |
| 2017-07-01 | 833.729 |
| 2017-08-01 | 820.535 |
| 2017-09-01 | 829.543 |
| 2017-10-01 | 826.37 |
| 2017-11-01 | 825.586 |
| 2017-12-01 | 811.242 |
| 2018-01-01 | 817.363 |
| 2018-02-01 | 815.096 |
| 2018-03-01 | 811.336 |
| 2018-04-01 | 810.931 |
| 2018-05-01 | 814.043 |
| 2018-06-01 | 813.26 |
| 2018-07-01 | 818.8 |
| 2018-08-01 | 826.181 |
| 2018-09-01 | 819.839 |
| 2018-10-01 | 825.114 |
| 2018-11-01 | 830.281 |
| 2018-12-01 | 831.305 |
| 2019-01-01 | 827.039 |
| 2019-02-01 | 829.319 |
| 2019-03-01 | 821.889 |
| 2019-04-01 | 818.721 |
| 2019-05-01 | 813.285 |
| 2019-06-01 | 810.02 |
| 2019-07-01 | 810.526 |
| 2019-08-01 | 803.554 |
| 2019-09-01 | 809.075 |
| 2019-10-01 | 806.084 |
| 2019-11-01 | 796.593 |
| 2019-12-01 | 790.583 |
| 2020-01-01 | 782.26 |
| 2020-02-01 | 777.72 |
| 2020-03-01 | 779.742 |
| 2020-04-01 | 786.26 |
| 2020-05-01 | 791.503 |
| 2020-06-01 | 796.863 |
| 2020-07-01 | 789.798 |
| 2020-08-01 | 794.086 |
| 2020-09-01 | 801.479 |
| 2020-10-01 | 804.417 |
| 2020-11-01 | 801.762 |
| 2020-12-01 | 812.602 |
| 2021-01-01 | 814.395 |
| 2021-02-01 | 821.02 |
| 2021-03-01 | 825.289 |
| 2021-04-01 | 812.616 |
| 2021-05-01 | 807.117 |
| 2021-06-01 | 800.355 |
| 2021-07-01 | 805.548 |
| 2021-08-01 | 796.939 |
| 2021-09-01 | 787.404 |
| 2021-10-01 | 783.448 |
| 2021-11-01 | 789.276 |
| 2021-12-01 | 781.904 |
| 2022-01-01 | 789.98 |
| 2022-02-01 | 784.679 |
| 2022-03-01 | 780.236 |
| 2022-04-01 | 789.675 |
| 2022-05-01 | 797.416 |
| 2022-06-01 | 791.802 |
| 2022-07-01 | 782.432 |
| 2022-08-01 | 782.785 |
| 2022-09-01 | 774.026 |
| 2022-10-01 | 770.499 |
| 2022-11-01 | 762.115 |
| 2022-12-01 | 757.724 |
| 2023-01-01 | 753.301 |
| 2023-02-01 | 752.02 |
| 2023-03-01 | 751.584 |
| 2023-04-01 | 740.886 |
| 2023-05-01 | 738.517 |
| 2023-06-01 | 743.591 |
| 2023-07-01 | 736.975 |
| 2023-08-01 | 737.28 |
| 2023-09-01 | 738.177 |
| 2023-10-01 | 739.227 |
| 2023-11-01 | 742.44 |
| 2023-12-01 | 737.843 |
| 2024-01-01 | 737.884 |
| 2024-02-01 | 730.262 |
| 2024-03-01 | 683.282 |

Rolling average vasectomies and births per 100,000 male and female population respectively for 35-44 year olds from April 2016 to December 2024

Vasectomies per 100,000 male population

Births per 100,000 female population

| Month and year | Vasectomies |
| --- | --- |
| 2016-04-01 | 87.6936 |
| 2016-05-01 | 92.3876 |
| 2016-06-01 | 95.1666 |
| 2016-07-01 | 95.2854 |
| 2016-08-01 | 100.234 |
| 2016-09-01 | 102.669 |
| 2016-10-01 | 104.16 |
| 2016-11-01 | 99.7636 |
| 2016-12-01 | 96.7667 |
| 2017-01-01 | 95.3851 |
| 2017-02-01 | 95.7008 |
| 2017-03-01 | 98.2946 |
| 2017-04-01 | 96.87 |
| 2017-05-01 | 94.3075 |
| 2017-06-01 | 88.0709 |
| 2017-07-01 | 93.0991 |
| 2017-08-01 | 88.2354 |
| 2017-09-01 | 85.91 |
| 2017-10-01 | 86.6617 |
| 2017-11-01 | 88.2629 |
| 2017-12-01 | 90.7039 |
| 2018-01-01 | 94.5323 |
| 2018-02-01 | 97.7797 |
| 2018-03-01 | 97.6145 |
| 2018-04-01 | 98.0502 |
| 2018-05-01 | 98.1383 |
| 2018-06-01 | 101.095 |
| 2018-07-01 | 98.91 |
| 2018-08-01 | 102.606 |
| 2018-09-01 | 105.707 |
| 2018-10-01 | 107.429 |
| 2018-11-01 | 114.347 |
| 2018-12-01 | 111.127 |
| 2019-01-01 | 109.041 |
| 2019-02-01 | 108.567 |
| 2019-03-01 | 105.627 |
| 2019-04-01 | 108.199 |
| 2019-05-01 | 112.328 |
| 2019-06-01 | 112.71 |
| 2019-07-01 | 114.385 |
| 2019-08-01 | 112.803 |
| 2019-09-01 | 108.769 |
| 2019-10-01 | 109.81 |
| 2019-11-01 | 105.124 |
| 2019-12-01 | 111.597 |
| 2020-01-01 | 108.526 |
| 2020-02-01 | 106.701 |
| 2020-03-01 | 109.405 |
| 2020-04-01 | 104.148 |
| 2020-05-01 | 94.5501 |
| 2020-06-01 | 92.2092 |
| 2020-07-01 | 89.2631 |
| 2020-08-01 | 91.0621 |
| 2020-09-01 | 94.9777 |
| 2020-10-01 | 89.7567 |
| 2020-11-01 | 88.9413 |
| 2020-12-01 | 87.2988 |
| 2021-01-01 | 91.5729 |
| 2021-02-01 | 90.6202 |
| 2021-03-01 | 94.4458 |
| 2021-04-01 | 101.196 |
| 2021-05-01 | 105.145 |
| 2021-06-01 | 109.046 |
| 2021-07-01 | 112.328 |
| 2021-08-01 | 110.227 |
| 2021-09-01 | 106.076 |
| 2021-10-01 | 107.365 |
| 2021-11-01 | 105.527 |
| 2021-12-01 | 104.65 |
| 2022-01-01 | 104.41 |
| 2022-02-01 | 108.423 |
| 2022-03-01 | 112.169 |
| 2022-04-01 | 111.089 |
| 2022-05-01 | 114.018 |
| 2022-06-01 | 114.934 |
| 2022-07-01 | 112.853 |
| 2022-08-01 | 117.919 |
| 2022-09-01 | 119.367 |
| 2022-10-01 | 122.29 |
| 2022-11-01 | 126.882 |
| 2022-12-01 | 129.722 |
| 2023-01-01 | 129.788 |
| 2023-02-01 | 130.744 |
| 2023-03-01 | 126.213 |
| 2023-04-01 | 126.011 |
| 2023-05-01 | 127.934 |
| 2023-06-01 | 128.909 |
| 2023-07-01 | 131.363 |
| 2023-08-01 | 127.04 |
| 2023-09-01 | 128.061 |
| 2023-10-01 | 129.072 |
| 2023-11-01 | 123.629 |
| 2023-12-01 | 116.993 |
| 2024-01-01 | 118.999 |
| 2024-02-01 | 119.285 |
| 2024-03-01 | 116.939 |
| 2024-04-01 | 115.591 |
| 2024-05-01 | 114.408 |
| 2024-06-01 | 114.429 |
| 2024-07-01 | 112.547 |
| 2024-08-01 | 114.684 |
| 2024-09-01 | 117.507 |
| 2024-10-01 | 115.859 |
| 2024-11-01 | 120.801 |
| 2024-12-01 | 122.516 |

|  | Births |
| --- | --- |
| 2015-07-01 | 224.666 |
| 2015-08-01 | 229.32 |
| 2015-09-01 | 228.655 |
| 2015-10-01 | 229.716 |
| 2015-11-01 | 233.991 |
| 2015-12-01 | 241.687 |
| 2016-01-01 | 238.944 |
| 2016-02-01 | 244.201 |
| 2016-03-01 | 245.188 |
| 2016-04-01 | 245.807 |
| 2016-05-01 | 239.481 |
| 2016-06-01 | 239.598 |
| 2016-07-01 | 239.349 |
| 2016-08-01 | 233.486 |
| 2016-09-01 | 238.855 |
| 2016-10-01 | 233.454 |
| 2016-11-01 | 230.445 |
| 2016-12-01 | 225.304 |
| 2017-01-01 | 230.011 |
| 2017-02-01 | 227.768 |
| 2017-03-01 | 226.069 |
| 2017-04-01 | 228.868 |
| 2017-05-01 | 233.818 |
| 2017-06-01 | 233.915 |
| 2017-07-01 | 230.305 |
| 2017-08-01 | 231.485 |
| 2017-09-01 | 222.263 |
| 2017-10-01 | 222.614 |
| 2017-11-01 | 224.218 |
| 2017-12-01 | 227.179 |
| 2018-01-01 | 223.117 |
| 2018-02-01 | 221.988 |
| 2018-03-01 | 221.415 |
| 2018-04-01 | 223.349 |
| 2018-05-01 | 225.05 |
| 2018-06-01 | 217.551 |
| 2018-07-01 | 219.757 |
| 2018-08-01 | 222.409 |
| 2018-09-01 | 224.894 |
| 2018-10-01 | 232.985 |
| 2018-11-01 | 229.845 |
| 2018-12-01 | 230.352 |
| 2019-01-01 | 230.555 |
| 2019-02-01 | 231.774 |
| 2019-03-01 | 235.321 |
| 2019-04-01 | 232.901 |
| 2019-05-01 | 229.268 |
| 2019-06-01 | 236.119 |
| 2019-07-01 | 236.564 |
| 2019-08-01 | 237.403 |
| 2019-09-01 | 240.37 |
| 2019-10-01 | 237.387 |
| 2019-11-01 | 237.778 |
| 2019-12-01 | 227.409 |
| 2020-01-01 | 226.377 |
| 2020-02-01 | 222.598 |
| 2020-03-01 | 216.567 |
| 2020-04-01 | 212.832 |
| 2020-05-01 | 214.074 |
| 2020-06-01 | 212.699 |
| 2020-07-01 | 214.334 |
| 2020-08-01 | 213.425 |
| 2020-09-01 | 210.627 |
| 2020-10-01 | 208.848 |
| 2020-11-01 | 210.001 |
| 2020-12-01 | 215.033 |
| 2021-01-01 | 218.62 |
| 2021-02-01 | 224.898 |
| 2021-03-01 | 229.768 |
| 2021-04-01 | 237.769 |
| 2021-05-01 | 238.547 |
| 2021-06-01 | 238.772 |
| 2021-07-01 | 243.357 |
| 2021-08-01 | 241.971 |
| 2021-09-01 | 246.607 |
| 2021-10-01 | 247.379 |
| 2021-11-01 | 244.087 |
| 2021-12-01 | 246.112 |
| 2022-01-01 | 244.5 |
| 2022-02-01 | 240.515 |
| 2022-03-01 | 237.76 |
| 2022-04-01 | 231.459 |
| 2022-05-01 | 229.473 |
| 2022-06-01 | 227.203 |
| 2022-07-01 | 222.346 |
| 2022-08-01 | 221.544 |
| 2022-09-01 | 217.429 |
| 2022-10-01 | 219.249 |
| 2022-11-01 | 223.199 |
| 2022-12-01 | 221.315 |
| 2023-01-01 | 222.474 |
| 2023-02-01 | 222.253 |
| 2023-03-01 | 220.886 |
| 2023-04-01 | 224.652 |
| 2023-05-01 | 224.482 |
| 2023-06-01 | 224.519 |
| 2023-07-01 | 226.391 |
| 2023-08-01 | 227.808 |
| 2023-09-01 | 227.617 |
| 2023-10-01 | 225.73 |
| 2023-11-01 | 224.073 |
| 2023-12-01 | 223.18 |
| 2024-01-01 | 219.887 |
| 2024-02-01 | 217.093 |
| 2024-03-01 | 207.233 |

Australian Capital Territory

Rolling average vasectomies and births per 100,000 male and female population respectively for 18-24 year olds from April 2016 to December 2024

Vasectomies per 100,000 male population

Births per 100,000 female population

| Month and year | Vasectomies |
| --- | --- |
| 2016-04-01 | 0.894136 |
| 2016-05-01 | 0.812851 |
| 2016-06-01 | 0.745114 |
| 2016-07-01 | 0.745114 |
| 2016-08-01 | 0.745114 |
| 2016-09-01 | 1.11246 |
| 2016-10-01 | 1.11246 |
| 2016-11-01 | 1.11246 |
| 2016-12-01 | 1.11246 |
| 2017-01-01 | 1.11246 |
| 2017-02-01 | 1.11246 |
| 2017-03-01 | 0.74044 |
| 2017-04-01 | 0.36735 |
| 2017-05-01 | 0.36735 |
| 2017-06-01 | 0.36735 |
| 2017-07-01 | 0.36735 |
| 2017-08-01 | 0.724529 |
| 2017-09-01 | 0.357179 |
| 2017-10-01 | 0.357179 |
| 2017-11-01 | 0.357179 |
| 2017-12-01 | 0.357179 |
| 2018-01-01 | 0.357179 |
| 2018-02-01 | 0.357179 |
| 2018-03-01 | 0.710885 |
| 2018-04-01 | 0.710885 |
| 2018-05-01 | 0.710885 |
| 2018-06-01 | 0.710885 |
| 2018-07-01 | 0.710885 |
| 2018-08-01 | 0.353707 |
| 2018-09-01 | 0.353707 |
| 2018-10-01 | 0.353707 |
| 2018-11-01 | 0.353707 |
| 2018-12-01 | 0.353707 |
| 2019-01-01 | 0.353707 |
| 2019-02-01 | 0.353707 |
| 2019-03-01 | 0.347991 |
| 2019-04-01 | 0.347991 |
| 2019-05-01 | 0.347991 |
| 2019-06-01 | 0.347991 |
| 2019-07-01 | 0.347991 |
| 2019-08-01 | 0.347991 |
| 2019-09-01 | 0.347991 |
| 2019-10-01 | 0.347991 |
| 2019-11-01 | 0.347991 |
| 2019-12-01 | 0.347991 |
| 2020-01-01 | 0.347991 |
| 2020-02-01 | 0.347991 |
| 2020-03-01 | 0 |
| 2020-04-01 | 0 |
| 2020-05-01 | 0 |
| 2020-06-01 | 0 |
| 2020-07-01 | 0 |
| 2020-08-01 | 0 |
| 2020-09-01 | 0 |
| 2020-10-01 | 0 |
| 2020-11-01 | 0 |
| 2020-12-01 | 0.363615 |
| 2021-01-01 | 0.363615 |
| 2021-02-01 | 0.363615 |
| 2021-03-01 | 0.363615 |
| 2021-04-01 | 0.363615 |
| 2021-05-01 | 0.363615 |
| 2021-06-01 | 0.363615 |
| 2021-07-01 | 0.363615 |
| 2021-08-01 | 0.363615 |
| 2021-09-01 | 0.722517 |
| 2021-10-01 | 1.07705 |
| 2021-11-01 | 1.43159 |
| 2021-12-01 | 1.06797 |
| 2022-01-01 | 1.4117 |
| 2022-02-01 | 1.4117 |
| 2022-03-01 | 1.4117 |
| 2022-04-01 | 1.4117 |
| 2022-05-01 | 1.4117 |
| 2022-06-01 | 1.4117 |
| 2022-07-01 | 1.74679 |
| 2022-08-01 | 2.08188 |
| 2022-09-01 | 1.72298 |
| 2022-10-01 | 1.36844 |
| 2022-11-01 | 1.34567 |
| 2022-12-01 | 1.34567 |
| 2023-01-01 | 1.00195 |
| 2023-02-01 | 1.00195 |
| 2023-03-01 | 1.00195 |
| 2023-04-01 | 1.00195 |
| 2023-05-01 | 1.00195 |
| 2023-06-01 | 1.00195 |
| 2023-07-01 | 0.666857 |
| 2023-08-01 | 0.331767 |
| 2023-09-01 | 0.331767 |
| 2023-10-01 | 0.331767 |
| 2023-11-01 | 0.321502 |
| 2023-12-01 | 0.321502 |
| 2024-01-01 | 0.321502 |
| 2024-02-01 | 0.321502 |
| 2024-03-01 | 0.640262 |
| 2024-04-01 | 0.640262 |
| 2024-05-01 | 0.640262 |
| 2024-06-01 | 0.960196 |
| 2024-07-01 | 0.960196 |
| 2024-08-01 | 0.960196 |
| 2024-09-01 | 1.28251 |
| 2024-10-01 | 1.60696 |
| 2024-11-01 | 1.28546 |
| 2024-12-01 | 1.28546 |

|  | Births |
| --- | --- |
| 2015-07-01 | 193.666 |
| 2015-08-01 | 192.334 |
| 2015-09-01 | 192.372 |
| 2015-10-01 | 194.176 |
| 2015-11-01 | 191.465 |
| 2015-12-01 | 193.356 |
| 2016-01-01 | 200.169 |
| 2016-02-01 | 196.333 |
| 2016-03-01 | 193.985 |
| 2016-04-01 | 191.473 |
| 2016-05-01 | 189.679 |
| 2016-06-01 | 190.055 |
| 2016-07-01 | 185.653 |
| 2016-08-01 | 182.477 |
| 2016-09-01 | 183.658 |
| 2016-10-01 | 175.559 |
| 2016-11-01 | 174.56 |
| 2016-12-01 | 172.757 |
| 2017-01-01 | 168.519 |
| 2017-02-01 | 172.091 |
| 2017-03-01 | 170.534 |
| 2017-04-01 | 170.833 |
| 2017-05-01 | 167.538 |
| 2017-06-01 | 165.583 |
| 2017-07-01 | 167.322 |
| 2017-08-01 | 168.758 |
| 2017-09-01 | 164.334 |
| 2017-10-01 | 161.222 |
| 2017-11-01 | 162.666 |
| 2017-12-01 | 159.489 |
| 2018-01-01 | 155.743 |
| 2018-02-01 | 151.97 |
| 2018-03-01 | 152.161 |
| 2018-04-01 | 146.015 |
| 2018-05-01 | 143.772 |
| 2018-06-01 | 145.361 |
| 2018-07-01 | 146.062 |
| 2018-08-01 | 148.189 |
| 2018-09-01 | 146.045 |
| 2018-10-01 | 143.246 |
| 2018-11-01 | 142.964 |
| 2018-12-01 | 147.995 |
| 2019-01-01 | 151.829 |
| 2019-02-01 | 152.791 |
| 2019-03-01 | 151.587 |
| 2019-04-01 | 155.11 |
| 2019-05-01 | 153.229 |
| 2019-06-01 | 144.694 |
| 2019-07-01 | 138.838 |
| 2019-08-01 | 138.774 |
| 2019-09-01 | 139.345 |
| 2019-10-01 | 145.385 |
| 2019-11-01 | 144.006 |
| 2019-12-01 | 136.097 |
| 2020-01-01 | 134.336 |
| 2020-02-01 | 127.815 |
| 2020-03-01 | 130.063 |
| 2020-04-01 | 132.284 |
| 2020-05-01 | 134.128 |
| 2020-06-01 | 135.613 |
| 2020-07-01 | 135.845 |
| 2020-08-01 | 128.489 |
| 2020-09-01 | 127.991 |
| 2020-10-01 | 126.688 |
| 2020-11-01 | 121.05 |
| 2020-12-01 | 121.962 |
| 2021-01-01 | 121.597 |
| 2021-02-01 | 124.254 |
| 2021-03-01 | 123.133 |
| 2021-04-01 | 117.551 |
| 2021-05-01 | 117.719 |
| 2021-06-01 | 115.07 |
| 2021-07-01 | 113.596 |
| 2021-08-01 | 115.66 |
| 2021-09-01 | 113.142 |
| 2021-10-01 | 108.07 |
| 2021-11-01 | 112.943 |
| 2021-12-01 | 112.315 |
| 2022-01-01 | 109.409 |
| 2022-02-01 | 109.665 |
| 2022-03-01 | 104.712 |
| 2022-04-01 | 102.63 |
| 2022-05-01 | 99.4953 |
| 2022-06-01 | 102.008 |
| 2022-07-01 | 97.9695 |
| 2022-08-01 | 92.9017 |
| 2022-09-01 | 93.4527 |
| 2022-10-01 | 94.4334 |
| 2022-11-01 | 88.9832 |
| 2022-12-01 | 90.1176 |
| 2023-01-01 | 91.1106 |
| 2023-02-01 | 89.5152 |
| 2023-03-01 | 89.5936 |
| 2023-04-01 | 89.405 |
| 2023-05-01 | 89.2069 |
| 2023-06-01 | 85.403 |
| 2023-07-01 | 88.4595 |
| 2023-08-01 | 91.8333 |
| 2023-09-01 | 92.2558 |
| 2023-10-01 | 92.1624 |
| 2023-11-01 | 90.4813 |
| 2023-12-01 | 89.0779 |
| 2024-01-01 | 86.0671 |
| 2024-02-01 | 85.31 |
| 2024-03-01 | 82.3432 |

Rolling average vasectomies and births per 100,000 male and female population respectively for 25-34 year olds from April 2016 to December 2024

Vasectomies per 100,000 male population

Births per 100,000 female population

| Month and year | Vasectomies |
| --- | --- |
| 2016-04-01 | 22.7267 |
| 2016-05-01 | 22.2625 |
| 2016-06-01 | 22.6099 |
| 2016-07-01 | 21.6169 |
| 2016-08-01 | 22.5775 |
| 2016-09-01 | 23.0547 |
| 2016-10-01 | 21.7974 |
| 2016-11-01 | 21.0247 |
| 2016-12-01 | 20.2521 |
| 2017-01-01 | 20.2153 |
| 2017-02-01 | 20.1837 |
| 2017-03-01 | 21.1118 |
| 2017-04-01 | 21.0718 |
| 2017-05-01 | 20.5595 |
| 2017-06-01 | 19.791 |
| 2017-07-01 | 20.4688 |
| 2017-08-01 | 19.4474 |
| 2017-09-01 | 19.8749 |
| 2017-10-01 | 21.7316 |
| 2017-11-01 | 22.1594 |
| 2017-12-01 | 22.1152 |
| 2018-01-01 | 21.838 |
| 2018-02-01 | 22.2681 |
| 2018-03-01 | 22.4397 |
| 2018-04-01 | 23.3178 |
| 2018-05-01 | 23.9852 |
| 2018-06-01 | 25.1023 |
| 2018-07-01 | 25.0533 |
| 2018-08-01 | 27.3195 |
| 2018-09-01 | 27.2644 |
| 2018-10-01 | 25.8016 |
| 2018-11-01 | 26.2005 |
| 2018-12-01 | 27.0715 |
| 2019-01-01 | 28.172 |
| 2019-02-01 | 27.44 |
| 2019-03-01 | 26.9169 |
| 2019-04-01 | 26.6266 |
| 2019-05-01 | 25.6814 |
| 2019-06-01 | 24.9389 |
| 2019-07-01 | 24.8876 |
| 2019-08-01 | 23.8807 |
| 2019-09-01 | 24.9455 |
| 2019-10-01 | 26.0175 |
| 2019-11-01 | 26.395 |
| 2019-12-01 | 24.5411 |
| 2020-01-01 | 24.2492 |
| 2020-02-01 | 24.6592 |
| 2020-03-01 | 24.1591 |
| 2020-04-01 | 23.8731 |
| 2020-05-01 | 25.1697 |
| 2020-06-01 | 25.9941 |
| 2020-07-01 | 26.8263 |
| 2020-08-01 | 25.2219 |
| 2020-09-01 | 24.4881 |
| 2020-10-01 | 24.635 |
| 2020-11-01 | 24.9933 |
| 2020-12-01 | 24.9757 |
| 2021-01-01 | 24.4913 |
| 2021-02-01 | 24.6701 |
| 2021-03-01 | 25.2805 |
| 2021-04-01 | 25.0057 |
| 2021-05-01 | 26.431 |
| 2021-06-01 | 27.6229 |
| 2021-07-01 | 27.536 |
| 2021-08-01 | 28.7631 |
| 2021-09-01 | 29.3187 |
| 2021-10-01 | 29.4428 |
| 2021-11-01 | 29.3429 |
| 2021-12-01 | 32.061 |
| 2022-01-01 | 32.8328 |
| 2022-02-01 | 35.2699 |
| 2022-03-01 | 35.4018 |
| 2022-04-01 | 36.6002 |
| 2022-05-01 | 35.4775 |
| 2022-06-01 | 35.1719 |
| 2022-07-01 | 35.5115 |
| 2022-08-01 | 35.2339 |
| 2022-09-01 | 35.15 |
| 2022-10-01 | 34.6606 |
| 2022-11-01 | 34.7804 |
| 2022-12-01 | 34.0747 |
| 2023-01-01 | 33.6077 |
| 2023-02-01 | 33.1001 |
| 2023-03-01 | 33.0443 |
| 2023-04-01 | 33.1771 |
| 2023-05-01 | 32.9206 |
| 2023-06-01 | 30.4115 |
| 2023-07-01 | 29.1385 |
| 2023-08-01 | 30.6936 |
| 2023-09-01 | 31.2294 |
| 2023-10-01 | 33.9952 |
| 2023-11-01 | 35.1373 |
| 2023-12-01 | 34.8846 |
| 2024-01-01 | 34.2541 |
| 2024-02-01 | 32.3989 |
| 2024-03-01 | 32.9659 |
| 2024-04-01 | 31.336 |
| 2024-05-01 | 32.7184 |
| 2024-06-01 | 35.3114 |
| 2024-07-01 | 34.7069 |
| 2024-08-01 | 33.8986 |
| 2024-09-01 | 32.4887 |
| 2024-10-01 | 29.2831 |
| 2024-11-01 | 27.689 |
| 2024-12-01 | 26.2871 |

|  | Births |
| --- | --- |
| 2015-07-01 | 908.202 |
| 2015-08-01 | 911.145 |
| 2015-09-01 | 912.878 |
| 2015-10-01 | 920.25 |
| 2015-11-01 | 921.989 |
| 2015-12-01 | 914.323 |
| 2016-01-01 | 911.188 |
| 2016-02-01 | 911.806 |
| 2016-03-01 | 906.985 |
| 2016-04-01 | 906.157 |
| 2016-05-01 | 895.808 |
| 2016-06-01 | 890.029 |
| 2016-07-01 | 882.502 |
| 2016-08-01 | 873.401 |
| 2016-09-01 | 862.679 |
| 2016-10-01 | 848.374 |
| 2016-11-01 | 844.848 |
| 2016-12-01 | 844.903 |
| 2017-01-01 | 838.984 |
| 2017-02-01 | 834.39 |
| 2017-03-01 | 818.772 |
| 2017-04-01 | 805.495 |
| 2017-05-01 | 809.784 |
| 2017-06-01 | 802.117 |
| 2017-07-01 | 796.167 |
| 2017-08-01 | 788.9 |
| 2017-09-01 | 788.047 |
| 2017-10-01 | 775.304 |
| 2017-11-01 | 771.597 |
| 2017-12-01 | 764.515 |
| 2018-01-01 | 759.254 |
| 2018-02-01 | 754.166 |
| 2018-03-01 | 761.535 |
| 2018-04-01 | 759.559 |
| 2018-05-01 | 753.303 |
| 2018-06-01 | 752.619 |
| 2018-07-01 | 755.294 |
| 2018-08-01 | 757.182 |
| 2018-09-01 | 753.459 |
| 2018-10-01 | 757.011 |
| 2018-11-01 | 762.302 |
| 2018-12-01 | 764.851 |
| 2019-01-01 | 772.876 |
| 2019-02-01 | 772.919 |
| 2019-03-01 | 765.591 |
| 2019-04-01 | 766.383 |
| 2019-05-01 | 767.425 |
| 2019-06-01 | 762.962 |
| 2019-07-01 | 762.498 |
| 2019-08-01 | 756.357 |
| 2019-09-01 | 755.863 |
| 2019-10-01 | 756.86 |
| 2019-11-01 | 746.728 |
| 2019-12-01 | 743.146 |
| 2020-01-01 | 734.431 |
| 2020-02-01 | 733.866 |
| 2020-03-01 | 735.458 |
| 2020-04-01 | 735.499 |
| 2020-05-01 | 738.778 |
| 2020-06-01 | 750.24 |
| 2020-07-01 | 745.841 |
| 2020-08-01 | 744.668 |
| 2020-09-01 | 743.427 |
| 2020-10-01 | 738.843 |
| 2020-11-01 | 742.792 |
| 2020-12-01 | 745.032 |
| 2021-01-01 | 738.179 |
| 2021-02-01 | 743.881 |
| 2021-03-01 | 749.114 |
| 2021-04-01 | 749.172 |
| 2021-05-01 | 745.422 |
| 2021-06-01 | 742.309 |
| 2021-07-01 | 751.226 |
| 2021-08-01 | 754.522 |
| 2021-09-01 | 753.154 |
| 2021-10-01 | 748.823 |
| 2021-11-01 | 740.565 |
| 2021-12-01 | 733.583 |
| 2022-01-01 | 731.027 |
| 2022-02-01 | 721.686 |
| 2022-03-01 | 710.656 |
| 2022-04-01 | 701.428 |
| 2022-05-01 | 693.074 |
| 2022-06-01 | 677.405 |
| 2022-07-01 | 668.521 |
| 2022-08-01 | 664.396 |
| 2022-09-01 | 657.885 |
| 2022-10-01 | 660.988 |
| 2022-11-01 | 662.4 |
| 2022-12-01 | 656.828 |
| 2023-01-01 | 657.013 |
| 2023-02-01 | 657.763 |
| 2023-03-01 | 650.375 |
| 2023-04-01 | 649.967 |
| 2023-05-01 | 648.605 |
| 2023-06-01 | 648.223 |
| 2023-07-01 | 640.495 |
| 2023-08-01 | 635.323 |
| 2023-09-01 | 636.308 |
| 2023-10-01 | 630.904 |
| 2023-11-01 | 629.015 |
| 2023-12-01 | 629.591 |
| 2024-01-01 | 633.017 |
| 2024-02-01 | 624.86 |
| 2024-03-01 | 597.993 |

Rolling average vasectomies and births per 100,000 male and female population respectively for 35-44 year olds from April 2016 to December 2024

Vasectomies per 100,000 male population

Births per 100,000 female population

| Month and year | Vasectomies |
| --- | --- |
| 2016-04-01 | 85.9802 |
| 2016-05-01 | 86.2007 |
| 2016-06-01 | 87.5178 |
| 2016-07-01 | 88.7871 |
| 2016-08-01 | 91.767 |
| 2016-09-01 | 92.7641 |
| 2016-10-01 | 90.9059 |
| 2016-11-01 | 92.4439 |
| 2016-12-01 | 92.8669 |
| 2017-01-01 | 92.718 |
| 2017-02-01 | 98.4052 |
| 2017-03-01 | 104.661 |
| 2017-04-01 | 100.812 |
| 2017-05-01 | 98.9464 |
| 2017-06-01 | 98.9732 |
| 2017-07-01 | 95.9395 |
| 2017-08-01 | 95.3442 |
| 2017-09-01 | 94.5208 |
| 2017-10-01 | 95.3856 |
| 2017-11-01 | 97.5541 |
| 2017-12-01 | 98.1388 |
| 2018-01-01 | 99.8533 |
| 2018-02-01 | 97.9152 |
| 2018-03-01 | 98.3997 |
| 2018-04-01 | 98.2365 |
| 2018-05-01 | 102.344 |
| 2018-06-01 | 105.294 |
| 2018-07-01 | 107.219 |
| 2018-08-01 | 111.969 |
| 2018-09-01 | 111.205 |
| 2018-10-01 | 110.696 |
| 2018-11-01 | 109.567 |
| 2018-12-01 | 109.058 |
| 2019-01-01 | 105.945 |
| 2019-02-01 | 105.381 |
| 2019-03-01 | 101.874 |
| 2019-04-01 | 104.543 |
| 2019-05-01 | 101.908 |
| 2019-06-01 | 97.9211 |
| 2019-07-01 | 95.8604 |
| 2019-08-01 | 92.848 |
| 2019-09-01 | 98.013 |
| 2019-10-01 | 101.344 |
| 2019-11-01 | 103.328 |
| 2019-12-01 | 102.332 |
| 2020-01-01 | 107.502 |
| 2020-02-01 | 109.013 |
| 2020-03-01 | 110.791 |
| 2020-04-01 | 107.798 |
| 2020-05-01 | 110.101 |
| 2020-06-01 | 109.881 |
| 2020-07-01 | 115.186 |
| 2020-08-01 | 111.876 |
| 2020-09-01 | 111.03 |
| 2020-10-01 | 114.216 |
| 2020-11-01 | 113.163 |
| 2020-12-01 | 111.012 |
| 2021-01-01 | 112.003 |
| 2021-02-01 | 117.166 |
| 2021-03-01 | 118.151 |
| 2021-04-01 | 124.374 |
| 2021-05-01 | 125.601 |
| 2021-06-01 | 128.346 |
| 2021-07-01 | 128.286 |
| 2021-08-01 | 133.18 |
| 2021-09-01 | 133.126 |
| 2021-10-01 | 131.313 |
| 2021-11-01 | 131.997 |
| 2021-12-01 | 133.574 |
| 2022-01-01 | 131.377 |
| 2022-02-01 | 127.138 |
| 2022-03-01 | 127.809 |
| 2022-04-01 | 127.043 |
| 2022-05-01 | 127.962 |
| 2022-06-01 | 126.99 |
| 2022-07-01 | 125.514 |
| 2022-08-01 | 126.614 |
| 2022-09-01 | 125.144 |
| 2022-10-01 | 122.977 |
| 2022-11-01 | 123.889 |
| 2022-12-01 | 130.585 |
| 2023-01-01 | 132.026 |
| 2023-02-01 | 134.341 |
| 2023-03-01 | 141.091 |
| 2023-04-01 | 140.619 |
| 2023-05-01 | 140.822 |
| 2023-06-01 | 140.15 |
| 2023-07-01 | 139.45 |
| 2023-08-01 | 138.661 |
| 2023-09-01 | 140.28 |
| 2023-10-01 | 141.202 |
| 2023-11-01 | 135.856 |
| 2023-12-01 | 129.354 |
| 2024-01-01 | 126.15 |
| 2024-02-01 | 127.204 |
| 2024-03-01 | 119.917 |
| 2024-04-01 | 117.416 |
| 2024-05-01 | 118.285 |
| 2024-06-01 | 118.32 |
| 2024-07-01 | 114.256 |
| 2024-08-01 | 109.427 |
| 2024-09-01 | 105.765 |
| 2024-10-01 | 107.281 |
| 2024-11-01 | 110.467 |
| 2024-12-01 | 114.574 |

|  | Births |
| --- | --- |
| 2015-07-01 | 316.085 |
| 2015-08-01 | 317.739 |
| 2015-09-01 | 321.675 |
| 2015-10-01 | 321.183 |
| 2015-11-01 | 322.067 |
| 2015-12-01 | 327.474 |
| 2016-01-01 | 330.185 |
| 2016-02-01 | 331.393 |
| 2016-03-01 | 330.166 |
| 2016-04-01 | 334.7 |
| 2016-05-01 | 333.666 |
| 2016-06-01 | 329.448 |
| 2016-07-01 | 328.303 |
| 2016-08-01 | 330.431 |
| 2016-09-01 | 326.976 |
| 2016-10-01 | 326.231 |
| 2016-11-01 | 325.391 |
| 2016-12-01 | 323.874 |
| 2017-01-01 | 326.841 |
| 2017-02-01 | 322.137 |
| 2017-03-01 | 321.67 |
| 2017-04-01 | 319.958 |
| 2017-05-01 | 318.932 |
| 2017-06-01 | 316.252 |
| 2017-07-01 | 313.333 |
| 2017-08-01 | 313.49 |
| 2017-09-01 | 309.471 |
| 2017-10-01 | 313.975 |
| 2017-11-01 | 312.558 |
| 2017-12-01 | 304.411 |
| 2018-01-01 | 299.716 |
| 2018-02-01 | 299.65 |
| 2018-03-01 | 297.794 |
| 2018-04-01 | 296.636 |
| 2018-05-01 | 294.573 |
| 2018-06-01 | 304.973 |
| 2018-07-01 | 309.032 |
| 2018-08-01 | 306.918 |
| 2018-09-01 | 307.892 |
| 2018-10-01 | 310.448 |
| 2018-11-01 | 310.304 |
| 2018-12-01 | 315.628 |
| 2019-01-01 | 314.608 |
| 2019-02-01 | 314.112 |
| 2019-03-01 | 321.043 |
| 2019-04-01 | 321.238 |
| 2019-05-01 | 321.384 |
| 2019-06-01 | 316.556 |
| 2019-07-01 | 314.492 |
| 2019-08-01 | 312.386 |
| 2019-09-01 | 313.408 |
| 2019-10-01 | 311.755 |
| 2019-11-01 | 313.749 |
| 2019-12-01 | 309.524 |
| 2020-01-01 | 309.159 |
| 2020-02-01 | 309.522 |
| 2020-03-01 | 302.443 |
| 2020-04-01 | 302.867 |
| 2020-05-01 | 304.89 |
| 2020-06-01 | 307.905 |
| 2020-07-01 | 310.027 |
| 2020-08-01 | 313.557 |
| 2020-09-01 | 314.961 |
| 2020-10-01 | 314.213 |
| 2020-11-01 | 313.755 |
| 2020-12-01 | 316.983 |
| 2021-01-01 | 318.315 |
| 2021-02-01 | 318.679 |
| 2021-03-01 | 321.256 |
| 2021-04-01 | 318.547 |
| 2021-05-01 | 320.872 |
| 2021-06-01 | 313.569 |
| 2021-07-01 | 306.742 |
| 2021-08-01 | 299.402 |
| 2021-09-01 | 297.762 |
| 2021-10-01 | 295.24 |
| 2021-11-01 | 298.358 |
| 2021-12-01 | 293.313 |
| 2022-01-01 | 293.195 |
| 2022-02-01 | 290.063 |
| 2022-03-01 | 288.578 |
| 2022-04-01 | 286.616 |
| 2022-05-01 | 285.075 |
| 2022-06-01 | 287.486 |
| 2022-07-01 | 292.188 |
| 2022-08-01 | 297.754 |
| 2022-09-01 | 300.045 |
| 2022-10-01 | 297.332 |
| 2022-11-01 | 295.879 |
| 2022-12-01 | 295.099 |
| 2023-01-01 | 293.726 |
| 2023-02-01 | 294.701 |
| 2023-03-01 | 295.43 |
| 2023-04-01 | 297.572 |
| 2023-05-01 | 294.944 |
| 2023-06-01 | 294.048 |
| 2023-07-01 | 295.695 |
| 2023-08-01 | 289.906 |
| 2023-09-01 | 283.687 |
| 2023-10-01 | 285.176 |
| 2023-11-01 | 278.993 |
| 2023-12-01 | 279.871 |
| 2024-01-01 | 279.308 |
| 2024-02-01 | 278.104 |
| 2024-03-01 | 259.887 |

Northern Territory:

Rolling average vasectomies and births per 100,000 male and female population respectively for 18-24 year olds from April 2016 to December 2024

Vasectomies per 100,000 male population

Births per 100,000 female population

| Month and year | Vasectomies |
| --- | --- |
| 2016-04-01 | 0 |
| 2016-05-01 | 0 |
| 2016-06-01 | 0 |
| 2016-07-01 | 0 |
| 2016-08-01 | 0 |
| 2016-09-01 | 0 |
| 2016-10-01 | 0 |
| 2016-11-01 | 0 |
| 2016-12-01 | 0 |
| 2017-01-01 | 0 |
| 2017-02-01 | 0 |
| 2017-03-01 | 0 |
| 2017-04-01 | 0 |
| 2017-05-01 | 0 |
| 2017-06-01 | 0.651296 |
| 2017-07-01 | 0.651296 |
| 2017-08-01 | 0.651296 |
| 2017-09-01 | 0.651296 |
| 2017-10-01 | 0.651296 |
| 2017-11-01 | 0.651296 |
| 2017-12-01 | 0.651296 |
| 2018-01-01 | 0.651296 |
| 2018-02-01 | 0.651296 |
| 2018-03-01 | 0.651296 |
| 2018-04-01 | 0.651296 |
| 2018-05-01 | 0.651296 |
| 2018-06-01 | 0 |
| 2018-07-01 | 0 |
| 2018-08-01 | 0 |
| 2018-09-01 | 0 |
| 2018-10-01 | 0.672043 |
| 2018-11-01 | 0.672043 |
| 2018-12-01 | 0.672043 |
| 2019-01-01 | 0.672043 |
| 2019-02-01 | 0.672043 |
| 2019-03-01 | 0.672043 |
| 2019-04-01 | 0.672043 |
| 2019-05-01 | 0.672043 |
| 2019-06-01 | 0.672043 |
| 2019-07-01 | 0.672043 |
| 2019-08-01 | 0.672043 |
| 2019-09-01 | 0.672043 |
| 2019-10-01 | 0 |
| 2019-11-01 | 0 |
| 2019-12-01 | 0 |
| 2020-01-01 | 0 |
| 2020-02-01 | 0 |
| 2020-03-01 | 0 |
| 2020-04-01 | 0 |
| 2020-05-01 | 0 |
| 2020-06-01 | 0 |
| 2020-07-01 | 0 |
| 2020-08-01 | 0 |
| 2020-09-01 | 0 |
| 2020-10-01 | 0 |
| 2020-11-01 | 0 |
| 2020-12-01 | 0 |
| 2021-01-01 | 0 |
| 2021-02-01 | 0 |
| 2021-03-01 | 0 |
| 2021-04-01 | 0 |
| 2021-05-01 | 0.684238 |
| 2021-06-01 | 0.684238 |
| 2021-07-01 | 0.684238 |
| 2021-08-01 | 0.684238 |
| 2021-09-01 | 0.684238 |
| 2021-10-01 | 0.684238 |
| 2021-11-01 | 0.684238 |
| 2021-12-01 | 0.684238 |
| 2022-01-01 | 0.684238 |
| 2022-02-01 | 0.684238 |
| 2022-03-01 | 0.684238 |
| 2022-04-01 | 0.684238 |
| 2022-05-01 | 0 |
| 2022-06-01 | 0 |
| 2022-07-01 | 0 |
| 2022-08-01 | 0.644147 |
| 2022-09-01 | 0.644147 |
| 2022-10-01 | 0.644147 |
| 2022-11-01 | 0.644147 |
| 2022-12-01 | 0.644147 |
| 2023-01-01 | 0.644147 |
| 2023-02-01 | 0.644147 |
| 2023-03-01 | 0.644147 |
| 2023-04-01 | 0.644147 |
| 2023-05-01 | 1.26391 |
| 2023-06-01 | 1.26391 |
| 2023-07-01 | 1.26391 |
| 2023-08-01 | 0.619763 |
| 2023-09-01 | 0.619763 |
| 2023-10-01 | 0.619763 |
| 2023-11-01 | 0.619763 |
| 2023-12-01 | 0.619763 |
| 2024-01-01 | 0.619763 |
| 2024-02-01 | 0.619763 |
| 2024-03-01 | 0.619763 |
| 2024-04-01 | 0.619763 |
| 2024-05-01 | 0 |
| 2024-06-01 | 0 |
| 2024-07-01 | 0 |
| 2024-08-01 | 0 |
| 2024-09-01 | 0 |
| 2024-10-01 | 0 |
| 2024-11-01 | 0 |
| 2024-12-01 | 0 |

|  | Births |
| --- | --- |
| 2015-07-01 | 682.739 |
| 2015-08-01 | 680.893 |
| 2015-09-01 | 677.882 |
| 2015-10-01 | 669.462 |
| 2015-11-01 | 653.965 |
| 2015-12-01 | 657.706 |
| 2016-01-01 | 633.657 |
| 2016-02-01 | 636.496 |
| 2016-03-01 | 646.173 |
| 2016-04-01 | 645.048 |
| 2016-05-01 | 634.323 |
| 2016-06-01 | 632.541 |
| 2016-07-01 | 622.071 |
| 2016-08-01 | 622.682 |
| 2016-09-01 | 618.068 |
| 2016-10-01 | 613.236 |
| 2016-11-01 | 615.094 |
| 2016-12-01 | 607.305 |
| 2017-01-01 | 609.921 |
| 2017-02-01 | 610.346 |
| 2017-03-01 | 590.639 |
| 2017-04-01 | 587.139 |
| 2017-05-01 | 592.539 |
| 2017-06-01 | 580.056 |
| 2017-07-01 | 579.121 |
| 2017-08-01 | 574.464 |
| 2017-09-01 | 571.239 |
| 2017-10-01 | 571.909 |
| 2017-11-01 | 559.141 |
| 2017-12-01 | 562.802 |
| 2018-01-01 | 564.825 |
| 2018-02-01 | 557.044 |
| 2018-03-01 | 563.573 |
| 2018-04-01 | 557.05 |
| 2018-05-01 | 549.773 |
| 2018-06-01 | 554.596 |
| 2018-07-01 | 565.038 |
| 2018-08-01 | 565.593 |
| 2018-09-01 | 565.331 |
| 2018-10-01 | 562.996 |
| 2018-11-01 | 571.952 |
| 2018-12-01 | 575.631 |
| 2019-01-01 | 570.748 |
| 2019-02-01 | 562.899 |
| 2019-03-01 | 551.993 |
| 2019-04-01 | 558.425 |
| 2019-05-01 | 561.093 |
| 2019-06-01 | 550.92 |
| 2019-07-01 | 540.841 |
| 2019-08-01 | 530.041 |
| 2019-09-01 | 532.901 |
| 2019-10-01 | 541.614 |
| 2019-11-01 | 528.374 |
| 2019-12-01 | 512.87 |
| 2020-01-01 | 517.872 |
| 2020-02-01 | 512.905 |
| 2020-03-01 | 514.81 |
| 2020-04-01 | 506.417 |
| 2020-05-01 | 515.537 |
| 2020-06-01 | 518.446 |
| 2020-07-01 | 520.68 |
| 2020-08-01 | 525.842 |
| 2020-09-01 | 514.98 |
| 2020-10-01 | 508.802 |
| 2020-11-01 | 525.492 |
| 2020-12-01 | 520.603 |
| 2021-01-01 | 520.471 |
| 2021-02-01 | 538.581 |
| 2021-03-01 | 537.564 |
| 2021-04-01 | 535.411 |
| 2021-05-01 | 521.043 |
| 2021-06-01 | 522.725 |
| 2021-07-01 | 505.109 |
| 2021-08-01 | 504.4 |
| 2021-09-01 | 512.906 |
| 2021-10-01 | 510.279 |
| 2021-11-01 | 503.012 |
| 2021-12-01 | 514.261 |
| 2022-01-01 | 507.498 |
| 2022-02-01 | 488.591 |
| 2022-03-01 | 487.28 |
| 2022-04-01 | 470.146 |
| 2022-05-01 | 462.851 |
| 2022-06-01 | 460.071 |
| 2022-07-01 | 468.911 |
| 2022-08-01 | 467.763 |
| 2022-09-01 | 458.222 |
| 2022-10-01 | 450.152 |
| 2022-11-01 | 446.581 |
| 2022-12-01 | 444.568 |
| 2023-01-01 | 431.587 |
| 2023-02-01 | 449.671 |
| 2023-03-01 | 456.379 |
| 2023-04-01 | 471.449 |
| 2023-05-01 | 469.097 |
| 2023-06-01 | 468.997 |
| 2023-07-01 | 471.082 |
| 2023-08-01 | 473.17 |
| 2023-09-01 | 468.459 |
| 2023-10-01 | 484.72 |
| 2023-11-01 | 488.1 |
| 2023-12-01 | 483.09 |
| 2024-01-01 | 480.161 |
| 2024-02-01 | 465.151 |
| 2024-03-01 | 427.33 |

Rolling average vasectomies and births per 100,000 male and female population respectively for 25-34 year olds from April 2016 to December 2024

Vasectomies per 100,000 male population

Births per 100,000 female population

| Month and year | Vasectomies |
| --- | --- |
| 2016-04-01 | 8.98759 |
| 2016-05-01 | 8.54164 |
| 2016-06-01 | 7.82984 |
| 2016-07-01 | 8.17318 |
| 2016-08-01 | 7.83937 |
| 2016-09-01 | 8.52129 |
| 2016-10-01 | 7.49766 |
| 2016-11-01 | 7.15776 |
| 2016-12-01 | 6.47621 |
| 2017-01-01 | 7.16283 |
| 2017-02-01 | 8.19335 |
| 2017-03-01 | 9.22272 |
| 2017-04-01 | 10.5873 |
| 2017-05-01 | 11.2705 |
| 2017-06-01 | 12.6351 |
| 2017-07-01 | 11.9588 |
| 2017-08-01 | 13.0034 |
| 2017-09-01 | 13.0154 |
| 2017-10-01 | 15.802 |
| 2017-11-01 | 16.1704 |
| 2017-12-01 | 16.5321 |
| 2018-01-01 | 15.8559 |
| 2018-02-01 | 14.8462 |
| 2018-03-01 | 13.4752 |
| 2018-04-01 | 12.817 |
| 2018-05-01 | 12.4998 |
| 2018-06-01 | 12.1947 |
| 2018-07-01 | 13.2711 |
| 2018-08-01 | 12.2715 |
| 2018-09-01 | 13.0147 |
| 2018-10-01 | 10.2281 |
| 2018-11-01 | 11.343 |
| 2018-12-01 | 11.373 |
| 2019-01-01 | 11.3803 |
| 2019-02-01 | 11.395 |
| 2019-03-01 | 11.7544 |
| 2019-04-01 | 11.767 |
| 2019-05-01 | 11.4201 |
| 2019-06-01 | 11.0796 |
| 2019-07-01 | 10.3796 |
| 2019-08-01 | 10.7553 |
| 2019-09-01 | 10.0605 |
| 2019-10-01 | 10.4224 |
| 2019-11-01 | 9.36211 |
| 2019-12-01 | 9.37308 |
| 2020-01-01 | 10.4631 |
| 2020-02-01 | 11.5562 |
| 2020-03-01 | 12.2839 |
| 2020-04-01 | 13.3754 |
| 2020-05-01 | 13.7401 |
| 2020-06-01 | 14.4696 |
| 2020-07-01 | 16.2937 |
| 2020-08-01 | 15.9431 |
| 2020-09-01 | 15.2258 |
| 2020-10-01 | 16.6859 |
| 2020-11-01 | 16.3313 |
| 2020-12-01 | 17.0675 |
| 2021-01-01 | 15.6181 |
| 2021-02-01 | 14.5361 |
| 2021-03-01 | 15.2735 |
| 2021-04-01 | 14.9215 |
| 2021-05-01 | 15.2912 |
| 2021-06-01 | 15.3013 |
| 2021-07-01 | 15.2701 |
| 2021-08-01 | 15.9748 |
| 2021-09-01 | 16.6885 |
| 2021-10-01 | 17.0085 |
| 2021-11-01 | 16.9863 |
| 2021-12-01 | 16.2353 |
| 2022-01-01 | 16.2353 |
| 2022-02-01 | 16.2142 |
| 2022-03-01 | 15.8072 |
| 2022-04-01 | 14.7001 |
| 2022-05-01 | 13.9575 |
| 2022-06-01 | 13.553 |
| 2022-07-01 | 11.7341 |
| 2022-08-01 | 10.9806 |
| 2022-09-01 | 11.6399 |
| 2022-10-01 | 11.2186 |
| 2022-11-01 | 11.8683 |
| 2022-12-01 | 11.8298 |
| 2023-01-01 | 12.1714 |
| 2023-02-01 | 12.1461 |
| 2023-03-01 | 12.0954 |
| 2023-04-01 | 12.0826 |
| 2023-05-01 | 12.0698 |
| 2023-06-01 | 11.3544 |
| 2023-07-01 | 12.3444 |
| 2023-08-01 | 12.6503 |
| 2023-09-01 | 12.2615 |
| 2023-10-01 | 10.5407 |
| 2023-11-01 | 10.1618 |
| 2023-12-01 | 10.8067 |
| 2024-01-01 | 12.1417 |
| 2024-02-01 | 11.7937 |
| 2024-03-01 | 13.1098 |
| 2024-04-01 | 12.7713 |
| 2024-05-01 | 15.4327 |
| 2024-06-01 | 16.4274 |
| 2024-07-01 | 16.4072 |
| 2024-08-01 | 16.724 |
| 2024-09-01 | 17.7065 |
| 2024-10-01 | 20.0274 |
| 2024-11-01 | 20.3433 |
| 2024-12-01 | 19.9921 |

|  | Births |
| --- | --- |
| 2015-07-01 | 833.147 |
| 2015-08-01 | 838.757 |
| 2015-09-01 | 842.352 |
| 2015-10-01 | 837.887 |
| 2015-11-01 | 839.693 |
| 2015-12-01 | 843.079 |
| 2016-01-01 | 846.662 |
| 2016-02-01 | 848.231 |
| 2016-03-01 | 849.676 |
| 2016-04-01 | 853.545 |
| 2016-05-01 | 854.198 |
| 2016-06-01 | 841.824 |
| 2016-07-01 | 833.651 |
| 2016-08-01 | 837.602 |
| 2016-09-01 | 833.791 |
| 2016-10-01 | 838.47 |
| 2016-11-01 | 834.684 |
| 2016-12-01 | 828.261 |
| 2017-01-01 | 822.658 |
| 2017-02-01 | 815.391 |
| 2017-03-01 | 817.586 |
| 2017-04-01 | 809.921 |
| 2017-05-01 | 798.393 |
| 2017-06-01 | 803.862 |
| 2017-07-01 | 798.088 |
| 2017-08-01 | 796.892 |
| 2017-09-01 | 791.122 |
| 2017-10-01 | 792.645 |
| 2017-11-01 | 798.046 |
| 2017-12-01 | 787.889 |
| 2018-01-01 | 782.693 |
| 2018-02-01 | 792.362 |
| 2018-03-01 | 781.519 |
| 2018-04-01 | 776.923 |
| 2018-05-01 | 785.797 |
| 2018-06-01 | 780.883 |
| 2018-07-01 | 774.26 |
| 2018-08-01 | 759.521 |
| 2018-09-01 | 755.723 |
| 2018-10-01 | 746.214 |
| 2018-11-01 | 743.076 |
| 2018-12-01 | 747.015 |
| 2019-01-01 | 739.804 |
| 2019-02-01 | 731.163 |
| 2019-03-01 | 736.015 |
| 2019-04-01 | 740.189 |
| 2019-05-01 | 735.478 |
| 2019-06-01 | 736.092 |
| 2019-07-01 | 738.249 |
| 2019-08-01 | 744.697 |
| 2019-09-01 | 748.282 |
| 2019-10-01 | 757.96 |
| 2019-11-01 | 753.736 |
| 2019-12-01 | 755.928 |
| 2020-01-01 | 764.741 |
| 2020-02-01 | 755.752 |
| 2020-03-01 | 759.63 |
| 2020-04-01 | 770.552 |
| 2020-05-01 | 778.615 |
| 2020-06-01 | 778.5 |
| 2020-07-01 | 788.093 |
| 2020-08-01 | 796.973 |
| 2020-09-01 | 798.738 |
| 2020-10-01 | 789.459 |
| 2020-11-01 | 790.183 |
| 2020-12-01 | 790.161 |
| 2021-01-01 | 790.902 |
| 2021-02-01 | 800.952 |
| 2021-03-01 | 793.862 |
| 2021-04-01 | 785.591 |
| 2021-05-01 | 776.004 |
| 2021-06-01 | 781.54 |
| 2021-07-01 | 765.423 |
| 2021-08-01 | 741.849 |
| 2021-09-01 | 738.739 |
| 2021-10-01 | 741.626 |
| 2021-11-01 | 748.05 |
| 2021-12-01 | 737.707 |
| 2022-01-01 | 734.106 |
| 2022-02-01 | 733.692 |
| 2022-03-01 | 723.534 |
| 2022-04-01 | 715.218 |
| 2022-05-01 | 708.728 |
| 2022-06-01 | 696.881 |
| 2022-07-01 | 696.002 |
| 2022-08-01 | 709.875 |
| 2022-09-01 | 702.024 |
| 2022-10-01 | 691.21 |
| 2022-11-01 | 673.89 |
| 2022-12-01 | 678.97 |
| 2023-01-01 | 675.498 |
| 2023-02-01 | 665.537 |
| 2023-03-01 | 668.997 |
| 2023-04-01 | 661.127 |
| 2023-05-01 | 670.67 |
| 2023-06-01 | 675.711 |
| 2023-07-01 | 674.344 |
| 2023-08-01 | 674.931 |
| 2023-09-01 | 671.858 |
| 2023-10-01 | 679.059 |
| 2023-11-01 | 688.653 |
| 2023-12-01 | 684.903 |
| 2024-01-01 | 678.074 |
| 2024-02-01 | 665.087 |
| 2024-03-01 | 623.03 |

Rolling average vasectomies and births per 100,000 male and female population respectively for 35-44 year olds from April 2016 to December 2024

Vasectomies per 100,000 male population

Births per 100,000 female population

| Month and year | Vasectomies |
| --- | --- |
| 2016-04-01 | 50.2328 |
| 2016-05-01 | 48.5189 |
| 2016-06-01 | 48.3983 |
| 2016-07-01 | 44.9107 |
| 2016-08-01 | 43.6005 |
| 2016-09-01 | 41.8555 |
| 2016-10-01 | 41.4128 |
| 2016-11-01 | 39.662 |
| 2016-12-01 | 37.0377 |
| 2017-01-01 | 38.7827 |
| 2017-02-01 | 38.3472 |
| 2017-03-01 | 37.9116 |
| 2017-04-01 | 36.5979 |
| 2017-05-01 | 37.0275 |
| 2017-06-01 | 38.3244 |
| 2017-07-01 | 43.1039 |
| 2017-08-01 | 43.9671 |
| 2017-09-01 | 44.8327 |
| 2017-10-01 | 48.7561 |
| 2017-11-01 | 51.8078 |
| 2017-12-01 | 53.5527 |
| 2018-01-01 | 53.5605 |
| 2018-02-01 | 54.4406 |
| 2018-03-01 | 56.1944 |
| 2018-04-01 | 59.685 |
| 2018-05-01 | 59.2544 |
| 2018-06-01 | 58.8275 |
| 2018-07-01 | 54.9293 |
| 2018-08-01 | 55.3951 |
| 2018-09-01 | 56.29 |
| 2018-10-01 | 55.0376 |
| 2018-11-01 | 53.7725 |
| 2018-12-01 | 52.939 |
| 2019-01-01 | 50.7685 |
| 2019-02-01 | 49.4774 |
| 2019-03-01 | 50.3906 |
| 2019-04-01 | 46.0584 |
| 2019-05-01 | 46.9693 |
| 2019-06-01 | 43.9392 |
| 2019-07-01 | 43.9604 |
| 2019-08-01 | 44.4489 |
| 2019-09-01 | 45.8124 |
| 2019-10-01 | 44.0876 |
| 2019-11-01 | 45.0138 |
| 2019-12-01 | 43.7172 |
| 2020-01-01 | 46.4002 |
| 2020-02-01 | 45.9847 |
| 2020-03-01 | 43.3788 |
| 2020-04-01 | 45.614 |
| 2020-05-01 | 44.3082 |
| 2020-06-01 | 46.0964 |
| 2020-07-01 | 46.544 |
| 2020-08-01 | 43.4563 |
| 2020-09-01 | 42.1396 |
| 2020-10-01 | 42.1227 |
| 2020-11-01 | 42.1001 |
| 2020-12-01 | 45.624 |
| 2021-01-01 | 43.3894 |
| 2021-02-01 | 45.5796 |
| 2021-03-01 | 49.0882 |
| 2021-04-01 | 48.18 |
| 2021-05-01 | 49.0497 |
| 2021-06-01 | 49.9118 |
| 2021-07-01 | 54.239 |
| 2021-08-01 | 56.8266 |
| 2021-09-01 | 57.6432 |
| 2021-10-01 | 57.1308 |
| 2021-11-01 | 55.7257 |
| 2021-12-01 | 53.4369 |
| 2022-01-01 | 51.2188 |
| 2022-02-01 | 48.9398 |
| 2022-03-01 | 46.5997 |
| 2022-04-01 | 45.1974 |
| 2022-05-01 | 46.7897 |
| 2022-06-01 | 46.6326 |
| 2022-07-01 | 41.7563 |
| 2022-08-01 | 39.9053 |
| 2022-09-01 | 36.7898 |
| 2022-10-01 | 38.3417 |
| 2022-11-01 | 37.7884 |
| 2022-12-01 | 37.6531 |
| 2023-01-01 | 39.7144 |
| 2023-02-01 | 39.6308 |
| 2023-03-01 | 41.9679 |
| 2023-04-01 | 43.5343 |
| 2023-05-01 | 45.0101 |
| 2023-06-01 | 44.0383 |
| 2023-07-01 | 44.7798 |
| 2023-08-01 | 45.8993 |
| 2023-09-01 | 49.5041 |
| 2023-10-01 | 48.541 |
| 2023-11-01 | 52.0873 |
| 2023-12-01 | 51.1623 |
| 2024-01-01 | 55.1204 |
| 2024-02-01 | 55.0482 |
| 2024-03-01 | 57.262 |
| 2024-04-01 | 55.1642 |
| 2024-05-01 | 53.7964 |
| 2024-06-01 | 53.3064 |
| 2024-07-01 | 59.15 |
| 2024-08-01 | 63.7727 |
| 2024-09-01 | 60.8527 |
| 2024-10-01 | 66.5917 |
| 2024-11-01 | 64.0096 |
| 2024-12-01 | 67.8311 |

|  | Births |
| --- | --- |
| 2015-07-01 | 239.181 |
| 2015-08-01 | 246.704 |
| 2015-09-01 | 247.324 |
| 2015-10-01 | 246.261 |
| 2015-11-01 | 246.146 |
| 2015-12-01 | 253.518 |
| 2016-01-01 | 256.175 |
| 2016-02-01 | 267.73 |
| 2016-03-01 | 270.375 |
| 2016-04-01 | 272.004 |
| 2016-05-01 | 267.469 |
| 2016-06-01 | 266.286 |
| 2016-07-01 | 278.51 |
| 2016-08-01 | 277.658 |
| 2016-09-01 | 284.309 |
| 2016-10-01 | 278.965 |
| 2016-11-01 | 280.563 |
| 2016-12-01 | 271.429 |
| 2017-01-01 | 268.392 |
| 2017-02-01 | 261.579 |
| 2017-03-01 | 257.602 |
| 2017-04-01 | 259.57 |
| 2017-05-01 | 260.12 |
| 2017-06-01 | 272.656 |
| 2017-07-01 | 258.909 |
| 2017-08-01 | 261.256 |
| 2017-09-01 | 253.524 |
| 2017-10-01 | 256.961 |
| 2017-11-01 | 261.677 |
| 2017-12-01 | 257.783 |
| 2018-01-01 | 260.747 |
| 2018-02-01 | 265.495 |
| 2018-03-01 | 264.814 |
| 2018-04-01 | 267.291 |
| 2018-05-01 | 265.671 |
| 2018-06-01 | 257.163 |
| 2018-07-01 | 259.208 |
| 2018-08-01 | 258.893 |
| 2018-09-01 | 253.701 |
| 2018-10-01 | 256.547 |
| 2018-11-01 | 253.9 |
| 2018-12-01 | 257.709 |
| 2019-01-01 | 254.678 |
| 2019-02-01 | 252.463 |
| 2019-03-01 | 252.17 |
| 2019-04-01 | 247.186 |
| 2019-05-01 | 244.479 |
| 2019-06-01 | 245.748 |
| 2019-07-01 | 249.651 |
| 2019-08-01 | 246.337 |
| 2019-09-01 | 259.651 |
| 2019-10-01 | 264.068 |
| 2019-11-01 | 265.815 |
| 2019-12-01 | 267.661 |
| 2020-01-01 | 272.528 |
| 2020-02-01 | 274.215 |
| 2020-03-01 | 275.158 |
| 2020-04-01 | 281.29 |
| 2020-05-01 | 283.514 |
| 2020-06-01 | 291.736 |
| 2020-07-01 | 290.956 |
| 2020-08-01 | 292.357 |
| 2020-09-01 | 282.837 |
| 2020-10-01 | 278.11 |
| 2020-11-01 | 270.806 |
| 2020-12-01 | 270.058 |
| 2021-01-01 | 277.831 |
| 2021-02-01 | 272.682 |
| 2021-03-01 | 278.385 |
| 2021-04-01 | 276.108 |
| 2021-05-01 | 269.24 |
| 2021-06-01 | 262.988 |
| 2021-07-01 | 262.331 |
| 2021-08-01 | 262.049 |
| 2021-09-01 | 263.659 |
| 2021-10-01 | 267.295 |
| 2021-11-01 | 278.163 |
| 2021-12-01 | 280.167 |
| 2022-01-01 | 274.373 |
| 2022-02-01 | 279.315 |
| 2022-03-01 | 277.453 |
| 2022-04-01 | 269.812 |
| 2022-05-01 | 270.282 |
| 2022-06-01 | 261.334 |
| 2022-07-01 | 255.837 |
| 2022-08-01 | 255.255 |
| 2022-09-01 | 255.211 |
| 2022-10-01 | 249.804 |
| 2022-11-01 | 245.146 |
| 2022-12-01 | 243.509 |
| 2023-01-01 | 242.245 |
| 2023-02-01 | 237.697 |
| 2023-03-01 | 238.121 |
| 2023-04-01 | 238.22 |
| 2023-05-01 | 245.679 |
| 2023-06-01 | 252.692 |
| 2023-07-01 | 254.402 |
| 2023-08-01 | 251.111 |
| 2023-09-01 | 247.909 |
| 2023-10-01 | 246.76 |
| 2023-11-01 | 247.549 |
| 2023-12-01 | 245.585 |
| 2024-01-01 | 241.101 |
| 2024-02-01 | 240.704 |
| 2024-03-01 | 223.777 |
